# Characteristics and completeness of reporting of systematic reviews of prevalence studies in adult populations: a meta-research study

**DOI:** 10.1101/2024.01.31.24302077

**Authors:** Diana Buitrago-Garcia, William Gildardo Robles-Rodriguez, Javier Eslava-Schmalbach, Georgia Salanti, Nicola Low

## Abstract

**Objective:** The Preferred Reporting Items for Systematic reviews and Meta-Analyses (PRISMA) statement, first published in 2009, has been widely endorsed and compliance is high in systematic reviews of intervention studies. Systematic reviews of prevalence studies are increasing in frequency, but their characteristics and reporting quality have not been examined in large studies. Our objectives were to describe the characteristics of systematic reviews of prevalence studies in adults, evaluate the completeness of reporting and explore study-level characteristics associated with the completeness of reporting.

**Study design and setting:** We did a meta-research study. We searched 5 databases from January 2010 to December 2020 to identify systematic reviews of prevalence studies in adult populations. We used the PRISMA 2009 checklist to assess completeness of reporting and recorded additional characteristics. We conducted a descriptive analysis of review characteristics and linear regression to assess the relationship between compliance with PRISMA and publication characteristics.

**Results:** We included 1172 systematic reviews of prevalence studies. The number of reviews increased from 25 in 2010 to 273 in 2020. The median PRISMA score for systematic reviews without meta-analysis was 17.5 out of a maximum of 23 and, for systematic reviews with meta-analysis, 22 out of a maximum of 25. Completeness of reporting, particularly for key items in the methods section was suboptimal. Systematic reviews that included a meta-analysis or reported using a reporting or conduct guideline were the factors most strongly associated with increased compliance with PRISMA 2009.

**Conclusion:** Reporting of systematic reviews of prevalence was adequate for many PRISMA items. Nonetheless, this study highlights aspects for which special attention is needed. Development of a specific tool to assess the risk of bias in prevalence studies and an extension to the PRISMA statement could improve the conduct and reporting of systematic reviews of prevalence studies.

**Plain language summary:** A systematic review is a type of research study, which is used to summarise the available information from different studies about a specific topic, such as the prevalence of a disease. Meta-analysis is a statistical method for combining data from individual studies, which can be used to obtain the prevalence of the disease of interest in the populations studied in a systematic review.

The PRISMA statement (Preferred Reporting Items in Systematic Reviews and Meta-Analyses) is a guideline for researchers. It includes a checklist of all information that authors of a systematic review should include in their report. Many scientific journals ask authors to use the PRISMA statement. How well authors use the guideline to report the systematic reviews of prevalence studies is not known.

In our paper, we aimed to describe the characteristics of systematic reviews of studies of We included 1,172 systematic reviews of prevalence studies. The number of these reviews grew, from 25 in 2010 to 273 in 2020. Systematic review authors reported the information required for many items in the PRISMA checklist. Other items were reported less well, such as registering a protocol for the systematic review, assessing the risk of biased results in studies included in the review, reporting the methods planned for analysis, discussing limitations and reporting sources of funding. Systematic reviews of prevalence that included a meta-analysis (a statistical method to combine prevalence data) or followed a guideline were better at complying to the PRISMA 2009.

Our study suggests reporting of systematic reviews of prevalence might improve if there were an extension of the PRISMA statement specifically for systematic reviews of prevalence studies and if there were a new tool to assess the risk of bias in prevalence studies.

**What is new?:**

- Systematic reviews of prevalence increased from 25 in 2010 to 2020, to 273 in 2020.
- Reporting of systematic reviews of prevalence has improved but is still suboptimal.
- Reporting was better in reviews with a meta-analysis and which followed a guideline.
- Journals should encourage adherence to PRISMA for systematic reviews of prevalence.
- A risk of bias tool and a PRISMA extension for systematic reviews of prevalence should be developed.

## 1 Introduction

Prevalence studies quantify the occurrence of a disease and can be used to contribute to estimation of the burden of disease and as a measure to evaluate healthcare interventions [1, 2]. Systematic reviews (SRs) of prevalence studies allow the synthesis of evidence about prevalence, which also informs burden of disease estimates and provides a resource for policymakers to help set priorities [1]. The volume of SRs of prevalence studies is increasing, but the methods used to conduct them have been reported to be variable and suboptimal [3, 4].

The usefulness of any systematic review depends on the completeness of reporting and the information provided in the included publications. The Preferred Reporting Items for Systematic Reviews and Meta-Analyses (PRISMA) statement was first published in 2009 to help with transparent and complete reporting of systematic reviews that assess the benefits or harms of interventions [5]. Since then, extensions to the PRISMA statement have covered other study designs, including diagnostic test accuracy [6], protocols [7] and network meta-analysis [8]. An update to the statement, PRISMA 2020, included items that are also applicable to systematic reviews of aetiology, prognosis, and prevalence studies, whilst still being designed primarily for reviews of studies of health interventions [9]. Some studies have shown that the PRISMA statement and extensions have enhanced the reporting of systematic reviews [10–12], although others show that improvement is still needed [11, 13]. The completeness of reporting of SRs of prevalence is, to our knowledge, unknown. The objectives of this study were to describe the characteristics of SRs of prevalence studies in adults, the completeness of reporting, and to explore study level characteristics associated with the quality and completeness of reporting.

## 2 Methods

We conducted a meta-research study of SRs of prevalence studies, which were identified through a systematic review of SRs. The protocol for the systematic review of SRs was registered in the PROSPERO register (CRD42020151625). Differences between the methods in the protocol and the study reported here are in Appendix A. We report our findings according to the Guidelines for Reporting Meta-Epidemiological Research(Appendix B) [14].

### 2.1 Search methods

We searched MEDLINE-Ovid, Embase-Ovid, CINAHL and LILACS from January 2010 to December 2020 without language restrictions. We also searched grey literature in opengrey.com [15]. We used terms for “prevalence” and “systematic reviews” as medical subject heading (MeSH) terms, Emtree life science thesaurus terms and free text keywords to identify potential SRs that met our inclusion criteria(Appendix C).

### 2.2 Eligibility criteria

We included SRs of studies conducted in adults (individuals aged ≥18 years) in any setting that assessed the prevalence of a disease, symptom, risk factor or behaviour as their primary aim. We excluded SRs of diagnostic test accuracy and of incidence studies, unless prevalence estimates were also presented separately. We also excluded overviews of SRs, studies that conducted a meta-analysis or pooled prevalence data without conducting a SR and conference abstracts, since it was not possible fully assess the completeness of reporting.

### 2.3 Study selection and data extraction

One author screened titles, abstracts and reviewed full-text reports for potential eligibility (D.B.G.) and a second author (W.R.) verified 20% of studies, using the online tool Rayyan [16] (96% agreement for title and abstract screening). One reviewer extracted data using a pre-piloted form (D.B.G.) and a second author verified the extraction in 20% of included studies (W.R.). We resolved disagreements through discussion. We extracted the following information: publication year, journal and Journal Impact Factor (Web of Science Journal Citation Reports 2022 or, if not available, the impact factor reported on the journal’s website) country of the first author (using the first affiliation, if more than one was listed), number of authors, medical speciality and targeted condition, population, primary objective, design of the included studies, geographic coverage, type of numerical data extracted from the included studies, number of studies included, tool reported to have been used to assess the risk of bias or quality in included studies, statistical methods, and approaches used to assess heterogeneity. In addition, if authors reported the use of guidelines or recommendations for SRs, such as the PRISMA checklist [5], the Reporting Guidelines for Meta-analyses of Observational Studies (MOOSE) [17], or for conduct, such as the Cochrane Handbook for Systematic Reviews of Interventions [18].

### 2.4 Assessment of the completeness of reporting

The completeness of reporting of each SR was assessed using the PRISMA checklist published in 2009 [5], which was appropriate for the publication dates of the included studies. The PRISMA 2009 checklist has 27 items. We decided to exclude two items related to reporting biases across studies, e.g., publication bias and other biases due to missing studies or missing results within studies (item 15 in the methods and item 22 in the results). The statistical methods used to assess these biases were developed for comparative studies, but their relevance and interpretation in evidence from prevalence studies is less clear and needs to be further investigated [19]. We assigned each of the 25 items one point if the item was adequately reported or no points if the item was not reported. For some items, we awarded half a point if the information was partially reported (Appendix D). The maximum score for SRs with a meta-analysis was 25 points, and 23 points without a meta-analysis (items 14 and 21 were not applicable).

### 2.5 Data analysis

We summarised the study characteristics (discipline, number of studies etc.) using proportions or medians with interquartile ranges (IQR). The completeness of reporting for each review was calculated as a) the achieved PRISMA reporting score, and b) the scaled reporting score, which was the achieved score divided by the maximum possible value; the scaled reporting score takes values between 0 and 100%. Completeness of reporting was summarised as, a) the median (IQR) PRISMA scores, and b) the proportion of SRs that completely reported, partially reported, or did not report, each PRISMA item. Suboptimal reporting for an item was defined as less than 70%, based on a previous study [10].

We conducted univariable and multivariable linear regression analyses to assess the relationship between the scaled reporting score and the year of publication, the Journal Impact Factor, the journal’s publishing model (open access or not), the number of co-authors, the number of studies included in the review, the use of a guideline to report or conduct the systematic review, the medical specialty and the type of review (SRs with or without a meta-analysis). All analyses were performed using R version 4.3.1 [20].

## 3 Results

We screened 9580 references and included 1172 systematic reviews, which fulfilled our inclusion criteria. The main reason for exclusion of potentially eligible reviews was because the primary aim of the review was not to assess prevalence (Figure 1, Appendix E).

**Figure 1.** PRISMA flowchart of the selection of the inclusion of systematic reviews of prevalence studies.

### 3.1 Characteristics of the reviews

The number of SRs of prevalence increased from 25 in 2010 to 273 in 2020 (Figure 2). There were 387 SRs without meta-analysis and 785 SRs with a meta-analysis. The median number of studies included in all SRs was 25 (IQR 14, 46). The SRs were published across 645 different journals, with PLOS ONE being the most frequent (n=46), followed by BMC Public Health (n=21), and BMC Infectious Diseases (n=20) (Table 1, Table S1). First authors were affiliated with institutions in 65 countries, amongst whom half were in five countries: the United Kingdom (n=155), the United States (n=120), Iran (n=107), Brazil (n=105), and China (n=98) (Figure S1, Table S2). Most SRs evaluated the prevalence of a medical condition or risk factor (n=1,036, 88%) and extracted worldwide prevalence data (765, 65%). About half of the SRs (n=565, 48.2%) were conducted to assess the prevalence of infectious diseases, psychiatric conditions, cardiology, and neurology (Figure S2, Table S3). Prevalence data were extracted from diverse populations, most commonly from general adult populations (n=436, 37.2%), adults with a specific condition (n=382, 32.6%), older populations (n=100, 8.5%), women (n=89, 7.6%) or workers (n=34, 2.9%) (Table 1, Table S4).

**Figure 2.** PRISMA score and number of publications (n) by year of publication and type of systematic review.

**Table 1.**
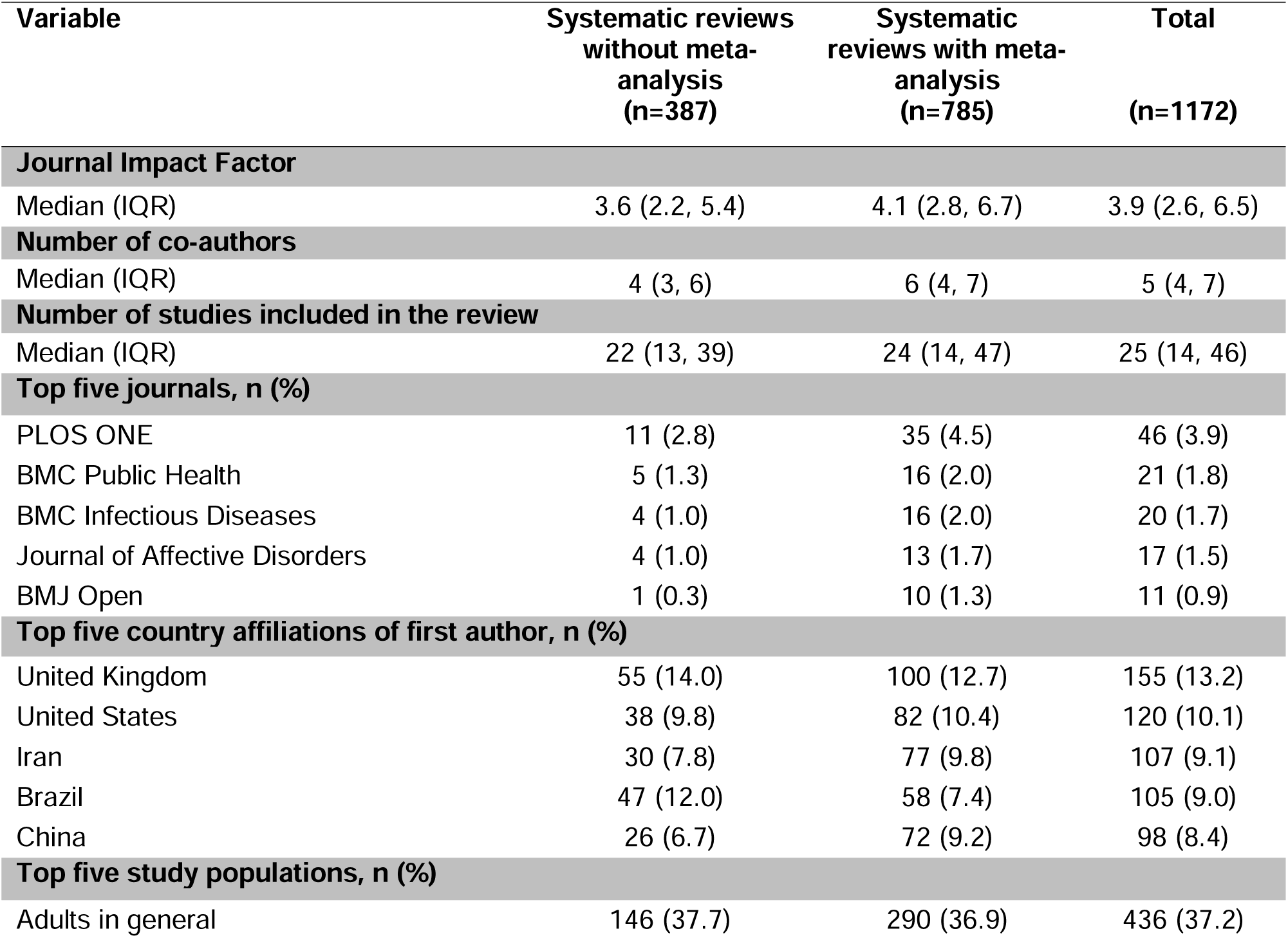
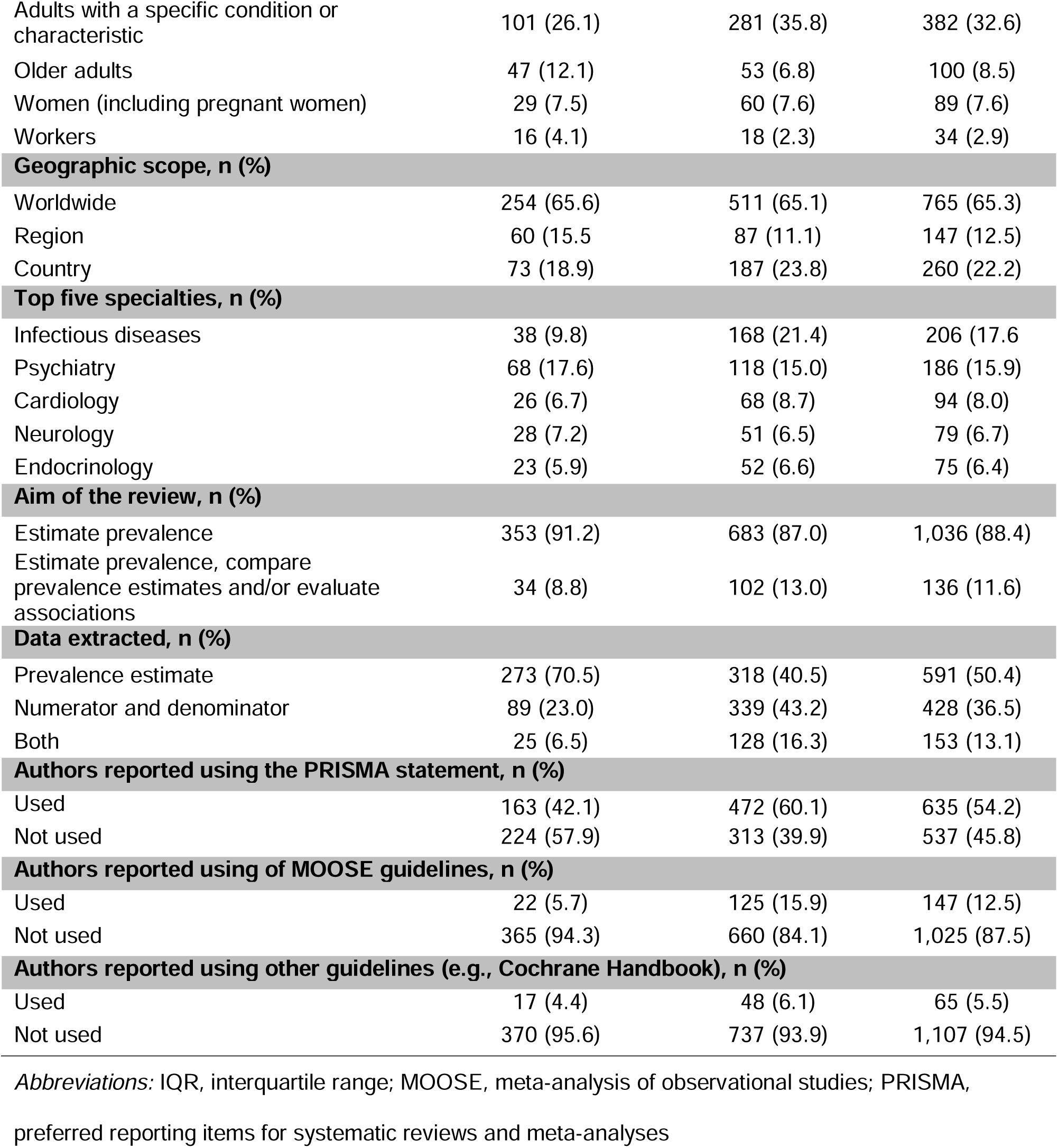
Characteristics of systematic reviews of prevalence studies in adults, published between 2010-2020

In total, 62% (727/1172) of SRs described using one or more reporting or conducting guidelines. The PRISMA checklist was cited most often, followed by the MOOSE guideline [17]. Most authors included studies that answered their review questions without restrictions on study design (n=623, 53%) (Table 1, Table S5-S6). Among SRs with meta-analysis, most (n=703, 90%) used a random-effects model (Table S6). Only 21% (n=163) of SRs reported a method used for statistical transformation of prevalence values, such as the Freeman-Tukey double arcsine, log, or logit functions. Heterogeneity between studies was assessed in most SRs, mainly using the I^2^ statistic (n=720; 92%).

### 3.2 Completeness of reporting

#### 3.2.1 PRISMA scores

The median PRISMA score for SRs without meta-analyisis was 17.5 (IQR 15.0,19.0) out of a possible 23, and for SRs with meta-analysis, it was 22.0 (IQR 20.5, 23.5) out of a possible 25 (Table S7). Figure 2 shows an increasing reporting score over the years. The median scaled reporting score for all included SRs was 84. 8% (IQR 76.0, 92.0).

#### 3.2.2 Reporting of selected PRISMA items

Over 80% of the included SRs complied with more than 70% of the PRISMA checklist items. Two of 387 SRs without meta-analysis (0.5%) and 66 of785 SRs with meta-analysis (8%) were entirely compliant with PRISMA 2009. Completeness of reporting was below 70% for reporting the existence of a protocol for the review, the search strategy, additional analyses and assessment of the risk of bias (Figure 3). Findings for these items are reported below.

**Figure 3.** Percentage of adequate reporting of PRISMA items in 2009 in 387 systematic reviews without meta-analysis (SR-M) and 785 systematic reviews with meta-analysis (SR+M). *^a^ (M), item in methods section; (R), item in results section*.

##### Protocol

Only 296/1172 (25%) of all SRs reported the existence of a protocol, which could be accessed, while 62/1172 (6%) mentioned a protocol without information on access details. For SRs without meta-analysis, the number with any protocol was 1/15 in 2010 and 9/47 in 2020; for SRs with meta-analysis, 0/10 reviews in 2010 and 91/226 in 2020 had a protocol (Table S8).

##### Search strategy

In 607/1172 (52%) SRs, authors adequately reported information about the search strategy for at least one database. In 498/1172 (42%) reviews, authors only reported the keywords used and 67/1172 (6%) did not provide any information.

##### Assessment of risk of bias

In the *methods section*, 798/1172 (68%) review authors reported the use of any tool to assess the risk of bias in included studies (Table S9). For 12 (1%), authors mentioned using a tool but did not report the items assessed or the tool. The most frequently reported tool was the Newcastle-Ottawa Scale (153/1172, 13.1%), which is for assessment of the quality of non-randomised studies [21]. In 213/1172 (18%) reviews, authors reported the use of a tool designed explicitly for assessment of quality or risk of bias in prevalence studies. The most frequent was the tool developed by the JBI (formerly Joanna Briggs Institute) [22]. The risk of bias in the included studies was adequately reported in the *results section* in 675/1172 (58%) reviews. Completeness of reporting of the study-level risk of bias assessment in the results was lower than in the methods section (68%). In 56/1172 reviews (5%) authors reported assessing the risk of bias but there was no description of this in the results. In 25/1172 reviews (2%), authors reported using the assessment of the quality of the studies to exclude studies from the review.

##### Additional analyses

526/785 (67%) SRs with meta-analysis reported in the *methods section* additional analyses such as sensitivity, subgroup analysis, or meta-regression. We observed an increase over the years, from 8/10 (80%) reviews in 2010 to 161/226 (71%) in 2020. Additional analysis results were presented adequately in the *results section* in 576/785 SRs with meta-analysis (73%).

##### Reporting of funding sources

764/1172 (65%) SRs reported their source of funding. The reporting of this item improved from 12/25 (48%) in 2010 to 202/273 (74%) in 2020.

### 3.3 Factors associated with the completeness of reporting

Inclusion of a meta-analysis in the SRs and citing the use of a reporting or methodological guideline were the factors most strongly associated with a higher scaled reporting score (Table 2). Publishing in an open access journal, the year of publication and the Journal Impact Factor were also positively associated with higher scaled reporting scores.

**Table 2.**
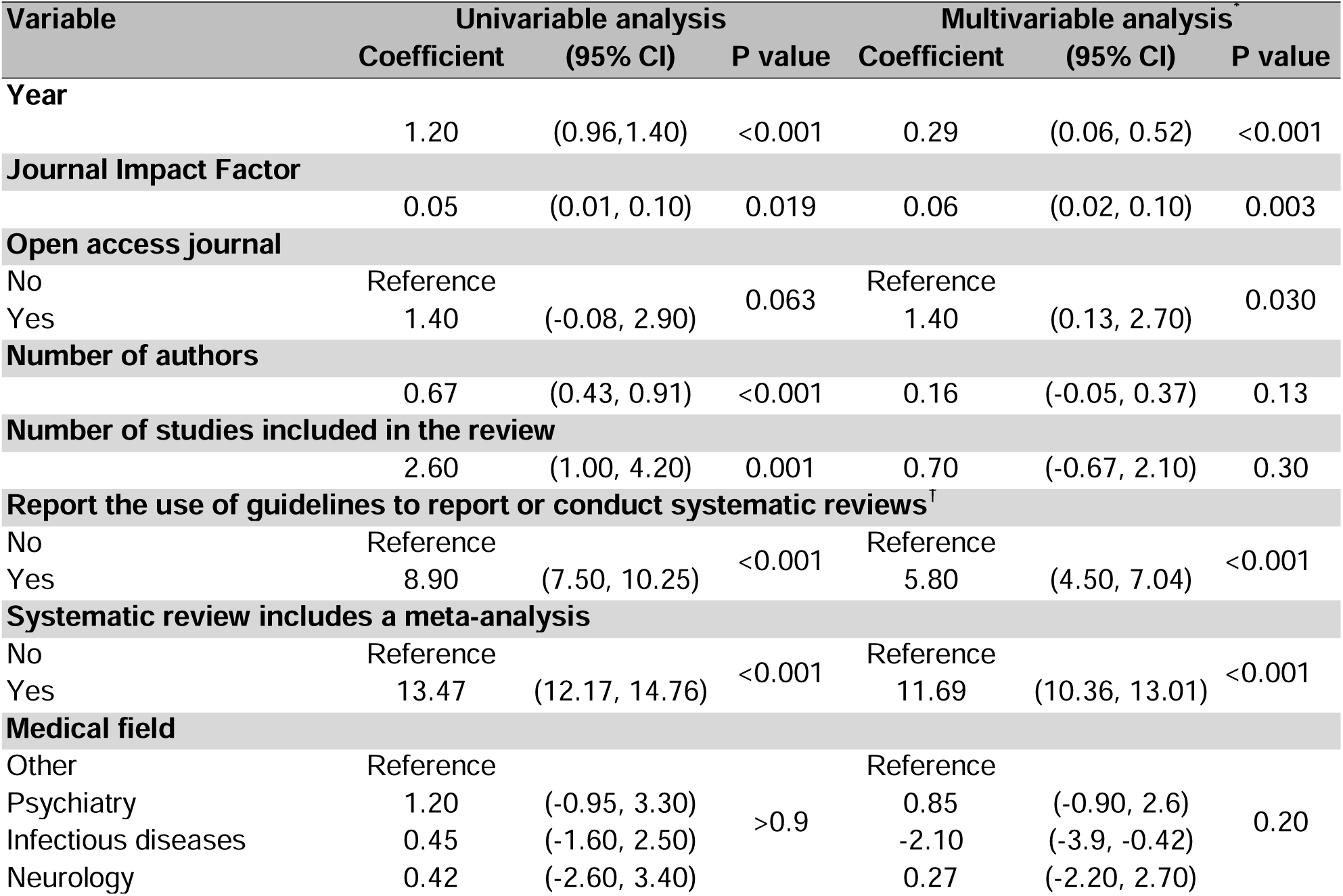
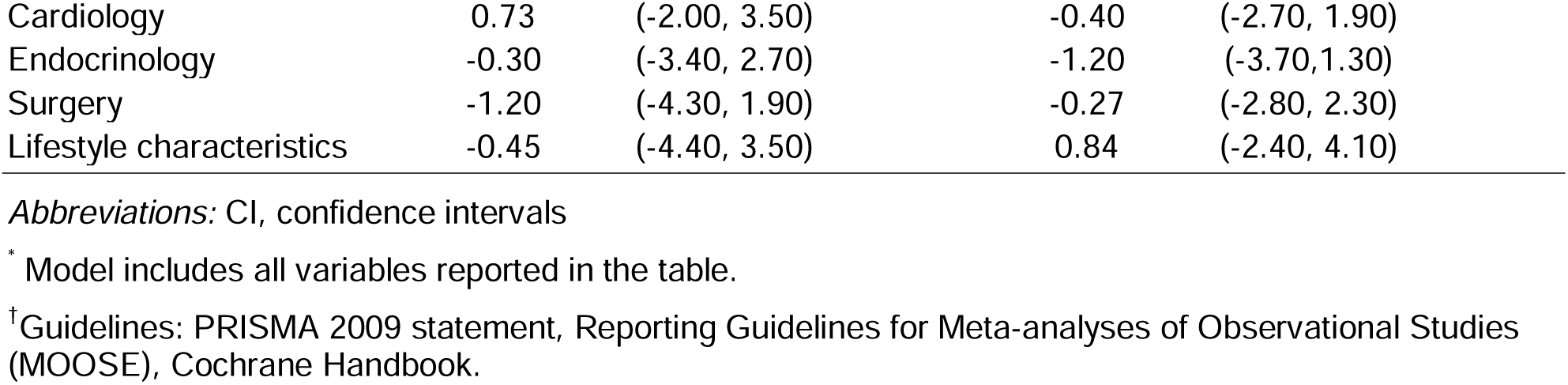
Univariable and multivariable linear regression between characteristics of the published systematic reviews of prevalence studies and scaled reporting score according to the PRISMA guidelines

## 4 Discussion

This meta-research study found an 11-fold increase in the number of SRs of prevalence in adult populations published from 25 in 2010 to 273 in 2020. The median PRISMA 2009 score for SRs without meta-analysis was 17.5 (IQR 15.0, 19.0), and for SRs with meta-analysis was 22.0, (IQR 20.5, 23.5). The items with the lowest compliance (<70%) were the availability of a protocol, search methods, assessment of the risk of bias in methods and results, additional analyses, and sources of funding. In multivariable analysis, SRs that included a meta-analysis, reported using a reporting or conduct guideline, and publications in more recent years, in an open access journal, or in journals with a higher Journal Impact Factor were on average more compliant with the PRISMA 2009 checklist.

### 4.1 Strengths and limitations

Strengths of this study include the detailed assessment of characteristics of SRs of prevalence, including 11 years of publications in 1172 SRs. In addition to recording whether PRISMA checklist items were reported, we extracted additional information for several items and conducted a multivariable regression analysis, which allowed more detailed interpretation of the findings than simple descriptive statistics. Our study also has limitations. First, we did not include SRs of prevalence published before the launch of the PRISMA statement in 2009 [5], which does not allow us to assess if there was improvement after the checklist was published. Second, we did not extend our search after 2020, so the end date of the search means that our findings correspond to the items and scope of the PRISMA 2009 statement [5]. The COVID-19 pandemic interrupted work on this study from 2021 and when we returned to it, the PRISMA 2020 checklist had been published [9]. Our study therefore provides an initial assessment of the completeness of reporting of SRs of prevalence and a future assessment will help to understand whether the extended scope of PRISMA 2020 is associated with further changes in the completeness of reporting. Third, we did not use PRISMA extensions, such as the PRISMA checklist for abstracts [23] or the extension for reporting literature searches in SRs [24], which might change the results of the items assessed with PRISMA 2009. Fourth, we limited the scope of topics to reviews conducted in adult populations, but we believe that reviews conducted in children would yield similar methodological findings. Fifth, we acknowledge that the PRISMA 2009 checklist was not designed to give a score. This method has been used previously [12] and, for our objectives, provided a pragmatic, if simplified, way to highlight aspects of reporting of SRs that could be improved.

### 4.2 Interpretation and comparison with other studies

Incomplete reporting of systematic reviews of prevalence should be seen in the context of published guidance for the conduct and reporting of SRs, most of which has been developed for randomised or non-randomised intervention studies. Whilst reported use of a guideline for reporting or conduct of SRs was associated with more complete reporting, the content of some items may indicate a lack of specific methodological guidance for prevalence studies. In particular, 30% of authors did not report the use of a tool for assessment of the risk of bias in individual studies and, amongst those that did, more than 30 different tools were used. In a systematic search, we identified 10 tools for assessing the risk of bias in prevalence studies [25], but only 284 (24%) of reviews in our study used one of these tools. Most of the tools listed were not designed for use with prevalence studies, such as the STROBE checklist for reporting of cross-sectional studies, which does not allow explicit assessment of risk of bias [26].

Completeness of reporting of SRs was associated with inclusion of a meta-analysis, publication in open access journals and publication in journals with a higher impact factor. These characteristics could be related to the level of experience and recognition of the methodological requirements of reporting of a systematic review team or with the expectations and requirements of journals.

We found two smaller studies, which assessed the characteristics of SRs of prevalence but did not use the PRISMA checklist to quantify completeness of reporting. Borges Migliavaca et al. [3] evaluated 235 SRs of prevalence published in 2017 and 2018 and found substantial differences in terms of conduct, reporting, risk of bias assessment and data synthesis. Whilst we decided not to assess the reporting of publication bias because of doubts about its relevance to prevalence studies, Borges Migliavaca et al. extracted this information. They found that 48/235 SRs examined publication bias either graphically or using a statistical test [3]. The authors also found that some reviews used the GRADE approach, despite the absence of GRADE guidance on assessing the quality of the body of evidence in a SR of prevalence. Hoffmann et al. [4] reported on 215 SRs of prevalence and incidence, identified from a random sample of publications up to 2018. The authors did not report on their findings for SRs of prevalence and incidence separately, but concluded that heterogeneity in characteristics, reporting, and methodology of these SRs might be due to the absence of specific guidance.

Reporting for some items in SRs of prevalence was consistent with other study designs. Page et al. [10] summarised meta-research studies of adherence to the PRISMA 2009 statement published up to mid-2017. They also found that items such as protocol registration and assessment of the risk of bias of individual studies were likely to be incomplete. Veroniki et al. [12] assessed the reporting in 1144 SRs with network meta-analysis and also found that the items least likely to be adequately reported were publication of a protocol (25%), and of a full search strategy (48%). Wasiak et al. [11] assessed 50 SRs in burn care management and concluded that methodology was the section most in need of improvement. They also found an improvement in the PRISMA score when the systematic review incorporated a meta-analysis.

## 5 Conclusions and recommendations

Reporting of SRs of prevalence was adequate for many PRISMA items. The completeness of reporting has also improved but there is room for improvement. There are items that authors who conduct any type of SR can improve without further guidance, such as the publication of a protocol. To improve the consistency and utility of SRs of prevalence more specific guidance about reporting of certain methodological features is required. Development of a specific tool to assess the risk of bias in prevalence studies and an extension to the PRISMA statement could improve the conduct and reporting SRs of prevalence studies.

## Declarations of interest

None

## Declarations of interest of generative AI in scientific writing

The authors declare that no AI was used in the scientific writing of the manuscript.

## Data Availability

Raw data and bibliographic details of the included studies are published on Open Science Framework https://osf.io/m5n6s/

## Acknowledgments

We would like to thank Yuli Baron for designing our graphical abstract.

## Author contributions

Study design: DBG, JES, NL, GS. Data collection: DBG, WR. Methodology DBG, JES, NL, GS Writing original draft: DBG

## Approval

All authors approved the final version of the manuscript.

## Funding

This work received support from the Swiss government excellence scholarship (grant number 2019.0774), the SSPH+ Global PhD Fellowship Programme in Public Health Sciences of the Swiss School of Public Health.

Georgia Salanti acknowledges the Swiss National Science Foundation for funding of her work through the NRP 78 initiative (Project 198418)

## Availability of data and materials

Raw data and bibliographic details of the included studies are published on Open Science Framework (https://osf.io/m5n6s/)

## Abbreviations

IQR: Interquartile ranges
MOOSE: Reporting Guidelines for Meta-analyses of Observational Studies
PRISMA: Preferred Reporting Items for Systematic Reviews and Meta-Analyses
SR: Systematic review

## Notes

### Competing Interest Statement

The authors have declared no competing interest.

### Summary of Updates

In this revision, we have revised the study title (from meta-epidemiological study to meta-research), conducted minor revisions in the text, and added a plain-language summary.

