## Appendices for "Characteristics and completeness of reporting of systematic reviews of prevalence studies in adult populations: a meta-research study"

***Appendix A. Differences between the published protocol and study***

**Protocol registration number (CRD42020151625)**

Our protocol was registered on 28 April 2020 and we did not update it.

The overall aim was to conduct a systematic review of systematic reviews of prevalence in adult populations, with an assessment of each step in the review process. For the methodological assessment of each included systematic reviews, we planned to use the AMSTAR-2 tool.

Since the registration of our protocol, two studies have shown the heterogeneity in methods for developing and conducting systematic reviews of prevalence, and observed that there is no standard methodology [1, 2]. We decided that a more useful contribution the evidence base would be to conduct a meta-research study, of the completeness of reporting of systematic reviews of prevalence with a structured assessment using the PRISMA statement.

The methods for identification and selection of potenitally eligible studies were largely the same as for the registered protocol.

***Summary of changes:***

- **Study design:** The current study is a meta-research study, rather than a systematic review of systematic reviews with methodological assessment of the included systematic reviews. We assessed the completeness reporting of the included systematic reviews of prevalence in adult populations using the PRISMA 2009 checklist [3].
- **Searches:** We updated our search up to December 2020 and we added an additional search in Open Grey[4].

- ***Types of study to be included***
    - *Inclusion Criteria:* No change.
    - *Exclusion Criteria:* No change.
- **Condition or domain being studied:** No change.
- **Participants/population:** No change.
- **Intervention(s), exposure(s):** No change.
- **Main outcomes:** No change.
- **Data extraction:** Due to time constraints during the COVID-19 pandemic, one author screened studies for inclusion, and a second author verified 20% of studies.

  In addition to the list of variables reported in the protocol, we extracted the Journal Impact Factor, the journal's publishing model, and the items listed PRISMA 2009 checklist [3].
- **Risk of bias assessment:** We did not assess the risk of bias in the included systematic reviews. Instead we assessed the completeness of reporting using the PRISMA 2009 checklist [3].
- **Strategy for data synthesis:** In addition to the descriptive analysis reported in the protocol, we conducted a univariable and multivariable linear regression analyses to assess the relationship between the completeness of reporting and the following variables:
    - Year of publication
    - Journal Impact Factor
    - Journal's publishing model (open access or not)
    - Number of co-authors
    - Number of studies included in the review
    - Use of a guideline to report or conduct the systematic review:
    - Medical specialty
    - Type of review (systematic review with or without a meta-analysis)
- **Analysis of subgroups or subsets:** No change.
- **Review team:** No change.

### *Appendix B. Items for reporting of meta-epidemiological research, adapted from the PRISMA Checklist*[1]

| Section and Topic | Item # | Checklist item | Location where item is reported |
|---|---|---|---|
| **TITLE** | | | |
| Title | 1 | Identify the report as a meta-epidemiologic study. | Page 1 |
| **ABSTRACT** | | | |
| Abstract | 2 | Provide a structured summary that includes the background of the topic, goal of the study, data sources, method of data selection, appraisal and synthesis methods, results, limitations, conclusions and implications of key findings. | Page 2 |
| **INTRODUCTION** | | | |
| Rationale | 3 | Describe the rationale for the meta-epidemiological study in the context of what is already known. | Page 5 |
| Objectives | 4 | Provide an explicit statement of the goal of the meta-epidemiological study and the hypothesis being empirically tested. | Page 5 |
| **METHODS** | | | |
| Protocol | 5 | Indicate if a protocol exists, if and where it can be accessed (eg, Web address). Registration of a protocol is not mandatory. | Page 6 |
| Eligibility criteria | 6 | Specify study characteristics used as criteria for eligibility with a rationale. | Page 6 |
| Information sources | 7 | Describe all information sources (eg, databases with dates of coverage, contact with experts to identify additional studies, Internet searches) and search date | Page 6 |
| Search | 8 | Present full electronic search strategy for at least one database, including any limits used, such that it could be repeated. Search is commonly not driven by a clinical question. | Page 6 Appendix C |
| Study selection | 9 | Describe the process for selecting studies for inclusion (ie, how many reviewers selected studies, reviewing in duplicate or by single individuals). | Page 6-7 |
| Data collection process | 10 | Describe method of data extraction from reports (eg, piloted forms, independently, in duplicate) and any processes used for manipulating data or obtaining and confirming data from investigators. | Page 6-8 |
| Data items | 11 | List and define all variables for which data were sought and any assumptions and imputations made. | Page 6 |
| Risk of bias in individual studies | 12 | If risk of bias assessment of individual studies was relevant to the analysis, describe the items used and how this information is to be used during data synthesis. | Not applicable |
| Summary measures | 13 | State the principal summary measures (eg, ratio of risk ratios, difference in means) and explain its meaning and direction to readers. | Page 7-8 |
| Synthesis of | 14 | Describe the statistical or descriptive methods of synthesis including measures of consistency if relevant. If applicable, describe | Page 7-8 |

| Section and Topic | Item # | Checklist item | Location where item is reported |
|---|---|---|---|
| results | | the development of statistical or simulation modelling based on theoretical background. Describe and justify assumptions and computational approximations. Describe methods of additional analyses (eg, sensitivity or subgroup analyses, meta-regression), if done, indicating which were prespecified. | |
| **RESULTS** | | | |
| Study selection | 15 | Give numbers of studies assessed for eligibility and included in the study, with reasons for exclusions at each stage, ideally with a flow diagram. Present a measure of inter-reviewer agreement (eg, kappa statistic). | Page 8 Figure 1 |
| Study characteristics | 16 | For each study, present characteristics for which data were extracted and provide the citations. Clinical characteristics may not always be relevant. | Page 8-12 Online repository |
| Risk of bias within studies | 17 | If risk of bias assessment of individual studies was used in the meta-epidemiological analysis, report risk of bias indicators of each study to allow replication of findings. | Not applicable |
| Results of individual studies | 18 | Present data elements used in the meta-epidemiological analysis from each study (results of clinical outcomes may not be relevant). | Page 8-12 |
| Synthesis of results | 19 | Present results of statistical analysis done, including measures of precision and measures of consistency. Present validity of assumptions and fit of statistical or simulation modelling, if applicable. | Page 8-12 |
| Additional analysis | 20 | Give results of additional analyses, if done (eg, sensitivity or subgroup analyses, meta-regression). | Page 13 |
| **DISCUSSION** | | | |
| Summary of evidence | 21 | Summarise the main findings and compare them with existing knowledge about the topic. The quality of evidence may not be relevant; however, investigators should describe their certainty in the results to readers. | Page 14 |
| Limitations | 22 | Discuss limitations at research methodology level (eg, likelihood of reporting or publication bias). | Page 14-15 |
| Conclusions | 23 | Provide general interpretation of the results and implications for future research. Provide any plausible impact on clinical practice. | Page 17 |
| **FUNDING** | | | |
| Funding | 27 | Describe sources of funding for the methodology research and role of funders. | Page 17 |

1. Murad MH, Wang Z. Guidelines for reporting meta-epidemiological methodology research. Evid Based Med. 2017;22(4):139-142.

doi: 10.1136/ebmed-2017-110713. Reference 14 in main text.

[4] OpenGrey. System for Information on Grey Literature in Europe 2020.

***Appendix  C. Search Strategy***

MEDLINE-OVID

Search Strategy:

--------------------------------------------------------------------------------

1 Prevalence/

2 prevalenc*.ti,ab.

3 prevalence stud*.ti,ab.

4 prevalence.mp.

5 or/1-4

6 (MEDLINE or systematic review).tw. or meta analysis.pt.

7 5 and 6

8 exp animals/ not humans.sh.

9 7 not 8 (

10 Adult/

11 adult*.ti,ab.

12 Young Adult/

13 Middle Aged/

14 Aged/

15 or/10-14

16 9 and 15

17 limit 16 to dt=20100101-20201231

EMBASE-ELSEVIER

--------------------------------------------------------------------------------

1 Prevalence/

2 prevalenc*.ti,ab.

3 prevalence stud*.ti,ab.

4 prevalence.mp.

5 or/1-4

6 (meta-analysis or systematic review).tw.

7 5 and 6

8 (animal$ not human$).sh,hw.

9 7 not 8

10 limit 9 to (adult <18 to 64 years> or aged <65+ years>)

11 limit 10 to yr="2010”

12 limit 11 to embase

CINAHL

S12 S9 AND S10 Limiters - Published Date: 20100101-20201231

S11 S9 AND S10

S10 adults or adult or aged or elderly

S9 S4 AND S8

S8 S5 OR S6 OR S7

S7 TI meta-analysis

S6 TI systematic review

S5 systematic review or meta-analysis

S4 S1 OR S2 OR S3

S3 prevalence stud*

S2 TI prevalence

S1 prevalence

LILACS

(tw:((tw:(tw:((tw:((tw:(prevalenc*)) OR (tw:(prevalenc* stud*)) OR (tw:(prevalencia)))) ) AND ((tw:((tw:(systematic review)) OR (tw:(meta-analysis )))) ) AND NOT ((tw:((tw:(children)) OR (tw:(child*)) OR (tw:(teen*)) OR (tw:(infant*)) OR (tw:(adolescent*))))) ))) ) AND ( db:("LILACS" OR "IBECS" OR "BDENF" OR "BRISA" OR "BBO" OR "CUMED" OR "BINACIS" OR "INDEXPSI" OR "LIPECS" OR "SES-SP")) AND (year_cluster:[2010 TO 2020])

OPEN GREY

prevalence AND (system* NEAR review)

***Appendix D.Criteria for assessing systematic reviews of prevalence according to the PRISMA 2009 checklist***

| Item no. | Item name | Response[a] | Judgment |
|---|---|---|---|
| 1 | Title | YES | Authors should identify their report as a systematic review or meta-analysis. |
| | | NO | The report is not identified as a as a systematic review or meta-analysis |
| 2 | Structured summary | YES | Provide a structured summary including the following as applicable: background; objectives; data sources; study eligibility criteria, participants, and interventions; study appraisal and synthesis methods; results; limitations; conclusions and implications of key findings; systematic review registration number. |
| | | NO | No abstract, or parts missing |
| 3 | Introduction-rationale | YES | Describe the rationale for the review in the context of what is already known |
| | | NO | No rationale described or, the context is minimum, and the importance of the review is not clearly described. |
| 4 | Introduction-objectives | YES | Provide an explicit statement of questions being addressed with reference to participants, interventions, comparisons, outcomes, and study design (PICOS)<br><br>*For our study specify the prevalence of the condition, disease, or risk factor*. |
| | | NO | No questions or objectives reported |
| **METHODS (11 items)** | | | |
| 5 | Protocol registration | YES | Indicate if a review protocol exists, if and where it can be accessed (e.g., web address), and, if available, provide registration information including registration number |
| | | PARTIAL | The authors report that a protocol exists but there is no information about how it can be accessed. |
| | | NO | No information about the protocol |
| 6 | Eligibility criteria | YES | Specify study characteristics (e.g., PICOS, length of follow-up) and report characteristics (e.g., years considered, language, publication status) used as criteria for eligibility, giving rationale.<br><br>**Notes:** *Since there is no explicit guidance* |

| | | | |
|---|---|---|---|
| | | | *about the studies to include for a systematic review of prevalence we assessed and extracted what the authors reported as eligibility criteria.* |
| | | NO | The inclusion criteria are not reported |
| 7 | Information sources | YES | Describe all information sources (e.g., databases with dates of coverage, contact with study authors to identify additional studies) in the search and date last searched |
| | | PARTIAL | Only some aspects are reported i.e., no date of search provided |
| | | NO | No information about the sources and databases searched for the review |
| 8 | Search | YES | Present full electronic search strategy for at least one database, including any limits used, such that it could be repeated |
| | | PARTIAL | Only keywords presented |
| | | NO | No information about the search strategy |
| 9 | Study selection | YES | State the process for selecting studies (i.e., screening, eligibility, included in the systematic review, and, if applicable, included in the meta-analysis) |
| | | NO | The selection process is not reported |
| 10 | Data collection process | YES | Describe the method of data extraction from reports (e.g., piloted forms, independently, in duplicate) and any processes for obtaining and confirming data from investigators |
| | | NO | The data collection process is not reported |
| 11 | Data items | YES | List and define all variables for which data were sought (e.g., PICOS, funding sources) and any assumptions and simplifications made. For our systematic review we extracted data about population and prevalence data (nominator, denominator, prevalence rate)<br>Data items are not completely reported specially information about prevalence |
| | | NO | The data items are not reported |
| 12 | Risk of bias in individual studies | YES | Describe methods used for assessing the risk of bias of individual studies (including specification of whether this was done at the study or outcome level) and how this information is to be used in any data synthesis |
| | | PARTIAL | The authors don't describe the tool used to assess risk of bias |

| | | | |
|---|---|---|---|
| | | NO | Information about risk of bias assessment not reported |
| 13 | Summary measures | YES | State the principal summary measures (e.g., risk ratio, the difference in means)- for our review Prevalence rates, pooled prevalence etc |
| | | NO | No information reported |
| 14 | Synthesis of results | YES | Describe the methods of handling data and combining results of studies, if done, including measures of consistency (e.g., I2) for each meta-analysis- explicit methods for meta-analyse prevalence<br><br>(a) Specify the effect measure<br>(b) The statistical method (such as inverse variance), and whether a fixed-effects or random-effects approach, or some other method (such<br>as Bayesian) or specific methods for transforming prevalence. If possible, authors should explain the reasons for those choices. |
| | | NO | No information reported |
| | | NOT APPLICABLE | The authors did not conduct a meta-analysis |
| 15 | Risk of bias across studies[b] | NOT APPLICABLE | Specify any assessment of risk of bias that may affect the cumulative evidence (e.g., publication bias, selective reporting within studies) |
| 16 | Additional analyses- | YES | Describe methods of additional analyses (e.g., sensitivity or subgroup analyses, meta-regression), if done, indicating which were pre-specified |
| | | NO | No information reported |
| **RESULTS (6 items)** | | | |
| 17 | Study selection | YES | Give numbers of studies screened, assessed for eligibility, and included in the review, with reasons for exclusions at each stage, ideally with a flow diagram |
| | | NO | No information not reported |
| 18 | Study characteristics | YES | For each study, present characteristics for which data were extracted (e.g., study size, PICOS, follow-up period) and provide the citations |
| | | PARTIAL | The authors partially report the characteristics of studies |
| | | NO | No information reported |

| | | | |
|---|---|---|---|
| 19 | Risk of bias within studies | YES | Present data on the risk of bias of each study and, if available, any outcome level assessment |
| | | PARTIAL | The authors partially report information about risk of bias |
| | | NO | No information reported |
| 20 | Results of individual studies | YES | For all outcomes present a simple summary data for each intervention group and intervention group and effect estimates and confidence intervals, ideally with a forest plot. (Prevalence proportion) |
| | | NO | No information reported |
| 21 | Synthesis of results | YES | Present results of each meta-analysis done, including confidence intervals and measures of consistency |
| | | NO | No information reported |
| | | NOT APPLICABLE | No meta-analysis conducted |
| 22 | Risk of bias across studies[b] | NOT APP | Specify any assessment of risk of bias that may affect the cumulative evidence (e.g., publication bias, selective reporting within studies) |
| 23 | Additional analysis | YES | Describe methods of additional analyses (e.g., sensitivity or subgroup analyses, meta-regression) |
| | | NO | No information reported but additional analyses planned in the methods section |
| **DISCUSSION (4 items)** | | | |
| 24 | Summary of evidence | YES | Summarize the main findings including the strength of evidence for each main outcome; consider their relevance to key groups (e.g., healthcare providers, users, and policymakers) |
| | | NO | No information reported |
| 25 | Limitations | YES | Discuss limitations at study and outcome level (e.g., risk of bias) and at review-level (e.g., incomplete retrieval of identified research, reporting bias) |
| | | NO | No information reported |
| 26 | Conclusions | YES | Provide a general interpretation of the results in the context of other evidence and implications for future research |
| | | NO | No information reported |
| 27 | Funding | YES | Describe sources of funding for the systematic review and other support |
| | | NO | No information reported |

a. Yes, 1 point; Partial, ½ point; No, 0 points. Maximum score, 23 if systematic review alone; 25 if systematic review with meta-analysis;

b. Item not scored for any study because the relevance to prevalence studies is not known.

### *Appendix E. List of excluded studies*

| AUTHOR | TITLE | REASONS FOR EXCLUSION |
|---|---|---|
| Adegbola, Richard A. | Carriage of Streptococcus pneumoniae and other respiratory bacterial pathogens in low and lower-middle income countries: a systematic review and meta-analysis | Merged adults and children |
| Adlan, A | Autonomic function and rheumatoid arthritis: a systematic review | Prevalence was not the primary outcome |
| Agbabiaka, T | Concurrent Use of Prescription Drugs and Herbal Medicinal Products in Older Adults: A Systematic Review | Prevalence was not the primary outcome |
| Aggarwal, Shilpa | Youth self-harm in low- and middle-income countries: Systematic review of the risk and protective factors | Wrong population |
| Agostini, Bernardo Antonio | How Common is Dry Mouth? Systematic Review and Meta-Regression Analysis of Prevalence Estimates | Not a SR of prevalence |
| Akioyamen, Leo E. | Estimating the prevalence of heterozygous familial hypercholesterolaemia: a systematic review and meta-analysis | Merged adults and children |
| Alexander C. Ford, Avantika Marwaha, Nicholas J. Talley, Allen Lim, Paul Moayyedi | Effect of validation status and type of dyspepsia questionnaire on the prevalence of dyspepsia: Systematic review and meta-analysis | Not a SR of prevalence |
| Anderson, Lesley | Prevalence of human papillomavirus in women attending cervical screening in the UK and Ireland: new data from northern Ireland and a systematic review and meta-analysis | Wrong study design |
| Araujo, Juliana de Oliveira | Prevalência de violência sexual em refugiados: uma revisão sistemática | Did not assess a disease, symptom, or lifestyle factor |
| Araújo, Maria Elizete de Almeida | Prevalência de utilização de serviços de saúde no Brasil: revisão sistemática e metanálise | Did not assess a disease, symptom, or lifestyle factor |
| Assis, Amanda Vervloet Dutra Agostinho | Prevalência de HPV na cavidade oral de indivíduos HIV+ e HIV-. Revisão sistemática e metanálise | Full text not available |
| Barker, M. M. | Prevalence and Incidence of Anxiety and Depression Among Children, Adolescents, and Young Adults With Life-Limiting Conditions | Merged adults and children |
| Bazyar, Jafar | The prevalence of sexual violence during pregnancy in Iran and the world: a systematic review and meta-analysis | Did not assess a disease, symptom, or lifestyle factor |

| | | |
|---|---|---|
| Beesley, Vanessa L. | A systematic literature review of the prevalence of and risk factors for supportive care needs among women with gynaecological cancer and their caregivers | Did not assess a disease, symptom, or lifestyle factor |
| Belyhun, Yeshambel | Hepatitis viruses in Ethiopia: a systematic review and meta-analysis | Prevalence was not the primary outcome |
| Bentohami, A. | Complications following volar locking plate fixation for distal radial fractures: a systematic review | Mixed incidence and prevalence data |
| Bertram, Melanie Y. | The disability adjusted life years due to stroke in South Africa in 2008 | Prevalence was not the primary outcome |
| Borges, M. K. | Frailty as a predictor of cognitive disorders: A systematic review and meta-analysis | Prevalence was not the primary outcome |
| Bruening, Meg | The Struggle Is Real: A Systematic Review of Food Insecurity on Postsecondary Education Campuses | Did not assess a disease, symptom, or lifestyle factor |
| Bussotti, Maurizio | Anxiety and depression in patients with pulmonary hypertension: impact and management challenges | Wrong study design |
| Cabanillas-Balsera, Daniel | Prevalence of Active Long-term Problems in Patients With Anorectal Malformations: A Systematic Review | Merged adults and children |
| Cao, Xiao-Lan | Prevalence of suicidal ideation and suicide attempts in the general population of China: A meta-analysis | Wrong study design |
| Carrara, Elena | Determinants of inappropriate empirical antibiotic treatment: systematic review and meta-analysis | Prevalence was not the primary outcome |
| Carvalho, Tatiana Lins | Depression and anxiety in individuals with amyotrophic lateral sclerosis: a systematic review | Prevalence was not the primary outcome |
| Chai, E. | Prevalence of type ii and type iii workplace violence against physicians: A systematic review and meta-analysis | Did not assess a disease, symptom, or lifestyle factor |
| Charlson, F. J. | Predicting the impact of the 2011 conflict in libya on population mental health: PTSD and depression prevalence and mental health service requirements | Not a SR of prevalence |
| Chen, Chen Hsiu | Current status of accurate prognostic awareness in advanced/terminally ill cancer patients: Systematic review and meta-regression analysis | Prevalence was not the primary outcome |
| Cooper, Katy L. | Prevalence of visits to five types of complementary and alternative medicine practitioners by the general population: a systematic review | Did not assess a disease, symptom, or lifestyle factor |

| | | |
|---|---|---|
| Cowgill, Karen D. | Obstetric fistula in low-resource countries: an under-valued and under-studied problem--systematic review of its incidence, prevalence, and association with stillbirth | Mixed incidence and prevalence data |
| Eades, Claire E. | Prevalence of impaired glucose regulation in Europe: a meta-analysis | Wrong study design |
| Elliott, Robert E. | The prevalence of the ponticulus posticus (arcuate foramen) and its importance in the Goel-Harms procedure: meta-analysis and review of the literature | Did not assess a disease, symptom, or lifestyle factor |
| El-Sayed, A. M. | Ethnic inequalities in obesity among children and adults in the UK: a systematic review of the literature | Merged adults and children |
| Emaneini, Mohammad | Nasal carriage rate of methicillin resistant Staphylococcus aureus among Iranian healthcare workers: a systematic review and meta-analysis | Did not assess a disease, symptom, or lifestyle factor |
| Fabrizi, Fabrizio | Hepatitis C virus increases the risk of kidney disease among HIV-positive patients: Systematic review and meta-analysis | Mixed incidence and prevalence data |
| Farhangi, M. A. | Higher dietary acid load potentially increases serum triglyceride and obesity prevalence in adults: An updated systematic review and meta-analysis | Prevalence was not the primary outcome |
| Fernandez-Garrido, Julio | Clinical features of prefrail older individuals and emerging peripheral biomarkers: a systematic review | Prevalence was not the primary outcome |
| Freeman | | Mixed incidence and prevalence data |
| Friend, Amanda J. | Mental health of long-term survivors of childhood and young adult cancer: A systematic review | Merged adults and children |
| Gell, Lucy | Alcohol consumption among the over 50s: international comparisons | Wrong study design |
| Geng, Qin | Comparison of comorbid depression between irritable bowel syndrome and inflammatory bowel disease: A meta-analysis of comparative studies | Prevalence was not the primary outcome |
| Gomes, Vanessa | Prevalence of medicine use among Brazilian adults: a systematic review | Did not assess a disease, symptom, or lifestyle factor |
| Gómez-Urquiza, Jose L. | Factores de riesgo y niveles de burnout en enfermeras de atención primaria: una revisión sistemática | Prevalence was not the primary outcome |
| Gonçalves, Mariana | Prevalence of Violence against Immigrant Women: A Systematic Review of the Literature | Did not assess a disease, symptom, or lifestyle factor |

| | | |
|---|---|---|
| Goutaki, Myrofora | Clinical manifestations in primary ciliary dyskinesia: systematic review and meta-analysis | Prevalence was not the primary outcome |
| Gregory, Julie | An examination of the prevalence of acute pain for hospitalised adult patients: a systematic review | Prevalence was not the primary outcome |
| Gu, B. | Comparison of the prevalence and changing resistance to nalidixic acid and ciprofloxacin of Shigella between Europe-America and Asia-Africa from 1998 to 2009 | Prevalence was not the primary outcome |
| Guan, Y. | The negative effect of urologic chronic pelvic pain syndrome on female sexual function: a systematic review and meta-analysis | Prevalence was not the primary outcome |
| Gupta, Rishi K. | Prevalence of tuberculosis in post-mortem studies of HIV-infected adults and children in resource-limited settings: a systematic review and meta-analysis | Merged adults and children |
| Harris, P. E. | Prevalence of complementary and alternative medicine (CAM) use by the general population: a systematic review and update | Did not assess a disease, symptom, or lifestyle factor |
| Harris, Philip E. | Prevalence of visits to massage therapists by the general population: a systematic review | Did not assess a disease, symptom, or lifestyle factor |
| Haysom, Leigh | Prevalence and Risk Factors for Methicillin-Resistant Staphylococcus aureus (MRSA) Infections in Custodial Populations: A Systematic Review | Mixed incidence and prevalence data |
| Hennequin-Hoenderdos, N. L. | The prevalence of oral and peri-oral piercings in young adults: a systematic review | Did not assess a disease, symptom, or lifestyle factor |
| Hernández-Romero, Hebe | Prevalence of Victimization and Perpetration of Sexual Aggression in Undergraduate Students: A Systematic Review 2008-2018 | Did not assess a disease, symptom, or lifestyle factor |
| Heydarpour, Pouria | Multiple Sclerosis Epidemiology in Middle East and North Africa: A Systematic Review and Meta-Analysis | Mixed incidence and prevalence data |
| Hidaka, Hiroshi | Clinical and bacteriological influence of diabetes mellitus on deep neck infection: Systematic review and meta-analysis | Prevalence was not the primary outcome |
| Hill-Taylor, B. | Application of the STOPP/START criteria: a systematic review of the prevalence of potentially inappropriate prescribing in older adults, and evidence of clinical, humanistic and economic impact | Prevalence was not the primary outcome |

| | | |
|---|---|---|
| Horneber, Markus | How many cancer patients use complementary and alternative medicine: a systematic review and metaanalysis | Prevalence was not the primary outcome |
| Hu, Y. J. | Available evidence of antibiotic resistance from extended-spectrum s-lactamase-producing enterobacteriaceae in paediatric patients in 20 countries: A systematic review and meta-analysis | Merged adults and children |
| Huang, Chang-Quan | Cognitive function and risk for depression in old age: a meta-analysis of published literature | Prevalence was not the primary outcome |
| Iraji, N. | Constipation in Iran: Sepahan systematic review no. 5 | Prevalence was not the primary outcome |
| Izquierdo, Y | Cambios radiográficos del penacho de la falange distal de las manos en pacientes con esclerosis sistémica. Revisión sistemática | Did not assess a disease, symptom, or lifestyle factor |
| Jahrami, H. | Eating disorders risk among medical students: a global systematic review and meta-analysis | Prevalence was not the primary outcome |
| Jha, Sunita R. | Frailty in advanced heart failure: a systematic review | Prevalence was not the primary outcome |
| John, J | The Burden of Typhoid and Paratyphoid in India: Systematic Review and Meta-analysis | Prevalence was not the primary outcome |
| Kaczor, Marcin P. | IL28B polymorphism (rs12979860) associated with clearance of HCV infection in Poland: systematic review of its prevalence in chronic hepatitis C patients and general population frequency | Prevalence was not the primary outcome |
| Kaiser, R. | Epidemiology, etiology, and types of severe adult brachial plexus injuries requiring surgical repair: systematic review and meta-analysis | Prevalence was not the primary outcome |
| Katsanos, A. H. | Restless legs syndrome and cerebrovascular/cardiovascular events: Systematic review and meta-analysis | Prevalence was not the primary outcome |
| Katsanos, Aristeidis H. | Complex atheromatous plaques in the descending aorta and the risk of stroke: a systematic review and meta-analysis | Prevalence was not the primary outcome |
| Khalifeh, Hind | Recent physical and sexual violence against adults with severe mental illness: a systematic review and meta-analysis | Did not assess a disease, symptom, or lifestyle factor |
| Knapik, Joseph J. | A systematic review and meta-analysis on the prevalence of dietary supplement use by military personnel | Did not assess a disease, symptom, or lifestyle factor |
| Knight, Tristan | Prevalence of tic disorders: a systematic review and meta-analysis | Merged adults and children |

| | | |
|---|---|---|
| Kok, Laura M. | The occurrence of musculoskeletal complaints among professional musicians: a systematic review | Not a SR of prevalence |
| Kolkhir, P. | Chronic spontaneous urticaria and internal parasites--a systematic review | Review on animal studies |
| Kraychete, Durval Campos | Clinical evidence on visceral pain. Systematic review | Prevalence was not the primary outcome |
| Lan, Xiao-peng | Immunotherapy of DC-CIK cells enhances the efficacy of chemotherapy for solid cancer: a meta-analysis of randomized controlled trials in Chinese patients | Prevalence was not the primary outcome |
| Landeiro, Filipa | Delayed Hospital Discharges of Older Patients: A Systematic Review on Prevalence and Costs | Did not assess a disease, symptom, or lifestyle factor |
| Lauzier, Francois | Clinical outcomes, predictors, and prevalence of anterior pituitary disorders following traumatic brain injury: a systematic review | Prevalence was not the primary outcome |
| Li, H. | Diabetes prevalence and determinants in adults in China mainland from 2000 to 2010: A systematic review | Prevalence was not the primary outcome |
| Li, Xi | Prevalence and trends of the abdominal aortic aneurysms epidemic in general population--a meta-analysis | Wrong study design |
| Liu, M. L. | [The prevalence of blindness caused by primary angle closure glaucoma in middle-aged Chinese population: a systematic review and meta-analysis] | Full text not available |
| MacLeod, Jana B. A. | Alcohol-related injury visits: do we know the true prevalence in U.S. trauma centres? | Wrong study design |
| Mann, Elizabeth A. | Comparison of mortality associated with sepsis in the burn, trauma, and general intensive care unit patient: a systematic review of the literature | Prevalence was not the primary outcome |
| Martinez-Gonzalez, Miguel A. | Low consumption of fruit and vegetables and risk of chronic disease: a review of the epidemiological evidence and temporal trends among Spanish graduates | Prevalence was not the primary outcome |
| May, Stephen | Centralization and directional preference: a systematic review | Did not assess a disease, symptom, or lifestyle factor |
| Mazzucco, Sara | Prevalence of patent foramen ovale in cryptogenic transient ischaemic attack and non-disabling stroke at older ages: a population-based study, systematic review, and meta-analysis | Wrong study design |
| Meer, I. | Prevalence of vitamin D deficiency among Turkish, Moroccan, Indian and sub- | Wrong study design |

| | | |
|---|---|---|
| | Sahara African populations in Europe and their countries of origin: an overview | |
| Melville, C. A. | Definitions, measurement and prevalence of sedentary behaviour in adults with intellectual disabilities - A systematic review | Prevalence was not the primary outcome |
| Merry, Sarah | E-cigarette use in New Zealand-a systematic review and narrative synthesis | Did not assess a disease, symptom, or lifestyle factor |
| Moran, Andrew E. | The global burden of ischemic heart disease in 1990 and 2010: the Global Burden of Disease 2010 study | Mixed incidence and prevalence data |
| Morhason-Bello, Imran O. | Reported oral and anal sex among adolescents and adults reporting heterosexual sex in sub-Saharan Africa: a systematic review | Merged adults and children |
| Morris, Brian J. | Circumcision and lifetime risk of urinary tract infection: a systematic review and meta-analysis | Merged adults and children |
| Murphy, Kevin | Substance use in young persons in Ireland, a systematic review | Merged adults and children |
| Nestler, Kai | Strength Training for Women as a Vehicle for Health Promotion at Work | Prevalence was not the primary outcome |
| Nicola Veronese | Association between urinary incontinence and frailty: a systematic review and meta-analysis | Prevalence was not the primary outcome |
| Nilsson, C. | Definitions, measurements and prevalence of fear of childbirth: A systematic review | Did not assess a disease, symptom, or lifestyle factor |
| Nogueira, Fabiana | Prevalência do uso e efeitos de recursos ergogênicos por praticantes de musculação nas academias brasileiras: uma revisão sistematizada | Did not assess a disease, symptom, or lifestyle factor |
| Nowe, E. | Cancer-related fatigue in adolescents and young adults: A systematic review of the literature | Merged adults and children |
| Pagotto, Valeria | [Self-assessment of health by older Brazilians: systematic review of the literature] | Did not assess a disease, symptom, or lifestyle factor |
| Peris, Franc | Prevención de la muerte súbita por miocardiopatía arritmogénica del ventrículo derecho en deportistas | Not a SR of prevalence |
| Porto De Toledo, Isabela | Prevalence of otologic signs and symptoms in adult patients with temporomandibular disorders: a systematic review and meta-analysis | Did not assess a disease, symptom, or lifestyle factor |
| Reitzel, R. A. | Epidemiology of Infectious and Noninfectious Catheter Complications in Patients Receiving Home Parenteral Nutrition: A Systematic Review and Meta-Analysis | Prevalence was not the primary outcome |
| Reus, L. | The effect of growth hormone treatment or physical training on motor performance in | Prevalence was not the primary outcome |

| | | |
|---|---|---|
| | Prader-Willi syndrome: A systematic review | |
| Rouhi, Azin | Prevalence and risk factors for liver fibrosis detected by transient elastography or shear wave elastography in inflammatory arthritis: a systematic review | Not a SR of prevalence |
| Santana, Inayara Oliveira de | Prevalência da violência contra o idoso no Brasil: revisão analítica | Did not assess a disease, symptom, or lifestyle factor |
| Schaan, C. W. | Prevalence of excessive screen time and TV viewing among Brazilian adolescents: a systematic review and meta-analysis | Did not assess a disease, symptom, or lifestyle factor |
| Schuster, Isabell | Prevalence of Sexual Aggression Victimization and Perpetration in Chile: A Systematic Review | Did not assess a disease, symptom, or lifestyle factor |
| Serrano, Julia | Oral lesions in Sjögren's syndrome: A systematic review | Not a SR of prevalence |
| Shoormasti, R. S. | Are the most common food allergens in an Iranian atopic population compatible with worldwide reports? A systemic review and meta-analysis with molecular classification of frequent allergens | Did not assess a disease, symptom, or lifestyle factor |
| Silva, Josy Maria de Pinho da | Concepts, prevalence and characteristics of severe maternal morbidity and near miss in Brazil: a systematic review | Prevalence was not the primary outcome |
| Silva, Solange Moreira da | Manifestações bucais na infecção pelo Vírus da Imunodeficiência Humana: uma revisão sistemática da literatura | Not a SR of prevalence |
| Sinha, Dhirendra N. | Global burden of all-cause and cause-specific mortality due to smokeless tobacco use: systematic review and meta-analysis | Prevalence was not the primary outcome |
| Kristyn Zajac, Kennedy, Caitlin E., Fonner, Virginia A., Armstrong, Kevin S., O'Reilly, Kevin R. and Sweat, Michael D. | A Systematic Review of the Effects of Behavioral Counseling on Sexual Risk Behaviors and HIV/STI Prevalence in Low- and Middle-Income Countries | Prevalence was not the primary outcome |
| Smith, Kristy L. | Relative Age Effects Across and Within Female Sport Contexts: A Systematic Review and Meta-Analysis | Did not assess a disease, symptom, or lifestyle factor |
| Solaski, Myrill | Contribution of socio-economic status on the prevalence of cerebral palsy: a systematic search and review | Prevalence was not the primary outcome |
| Spencer, A. H. | The prevalence and clinical characteristics of punding in Parkinson's disease | Wrong study design |
| Stock, Christian | Population-based prevalence estimates of history of colonoscopy or | Did not assess a disease, symptom, or lifestyle factor |

| | | |
|---|---|---|
| | sigmoidoscopy: review and analysis of recent trends | |
| Storms, Hannelore | Prevalence of inappropriate medication use in residential long-term care facilities for the elderly: A systematic review | Did not assess a disease, symptom, or lifestyle factor |
| Subota, Ann | The association between dementia and epilepsy: A systematic review and meta-analysis | Prevalence was not the primary outcome |
| Tavares, Ana Sofia R | Psychosocial factors and performance enhancing substances in gym users: A systematic review | Did not assess a disease, symptom, or lifestyle factor |
| Thai, Michele | Prevalence of statin-drug interactions in older people: a systematic review | Did not assess a disease, symptom, or lifestyle factor |
| Torres-Sánchez, I | Cognitive impairment in COPD: a systematic review | Prevalence was not the primary outcome |
| Trevisonno, Jordan | Physical urticaria: Review on classification, triggers and management with special focus on prevalence including a meta-analysis | Prevalence was not the primary outcome |
| Tsai, Chung-Fen | Comparing Risk Factor Profiles between Intracerebral Hemorrhage and Ischemic Stroke in Chinese and White Populations: Systematic Review and Meta-Analysis | Prevalence was not the primary outcome |
| Twomey, C. D. | Cross-sectional associations of depressive symptom severity and functioning with health service use by older people in low-and-middle income countries | Prevalence was not the primary outcome |
| Van Lancker, Aurelie | Prevalence of symptoms in older cancer patients receiving palliative care: a systematic review and meta-analysis. | Did not assess a disease, symptom, or lifestyle factor |
| van Rooij, F | Cognitive Impairment in Transient Ischemic Attack Patients: A Systematic Review | Prevalence was not the primary outcome |
| Vuong, Huy Gia | The changing characteristics and molecular profiles of papillary thyroid carcinoma over time: a systematic review | Prevalence was not the primary outcome |
| Walker, Zoe J. | Psychiatric disorders among people with cancer in low- and lower-middle-income countries: study protocol for a systematic review and meta-analysis | Protocol for a review |
| Waller, K. M. J. | Residual risk of infection with blood-borne viruses in potential organ donors at increased risk of infection: systematic review and meta-analysis | Prevalence was not the primary outcome |
| Wang, Meng-Yao | Contraceptive practices among unmarried women in China, 1982-2017: systematic review and meta-analysis | Did not assess a disease, symptom, or lifestyle factor |
| Warmling, Deise | Prevalência de violência por parceiro íntimo em idosos e | Did not assess a disease, symptom, or lifestyle factor |

| | | |
|---|---|---|
| | fatores associados: revisão sistemática | |
| Willgoss, Thomas G. | Review of risk factors and preventative strategies for fall-related injuries in people with intellectual disabilities | Not a SR of prevalence |
| Wilson, Lisa M. | Impact of tobacco control interventions on smoking initiation, cessation, and prevalence: a systematic review | Prevalence was not the primary outcome |
| Yammine, K | The prevalence of the sesamoid bones of the hand: a systematic review and meta-analysis | Did not assess a disease, symptom, or lifestyle factor |
| Yammine, K | The sesamoids of the feet in humans: a systematic review and meta-analysis | Did not assess a disease, symptom, or lifestyle factor |
| Yammine, K. | The anatomy and prevalence of the juncturae tendinum in the hands. A systematic review and meta-analysis | Did not assess a disease, symptom, or lifestyle factor |
| Yammine, Kaissar | Clinical prevalence of palmaris longus agenesis: a systematic review and meta-analysis | Did not assess a disease, symptom, or lifestyle factor |
| Yan, Elsie | A systematic review of prevalence and risk factors for elder abuse in Asia | Did not assess a disease, symptom, or lifestyle factor |
| Yang, Lin-Sheng | Prevalence of suicide attempts among college students in China: a meta-analysis | Wrong study design |
| Yee, A. | Clinical factors associated with sexual dysfunction among men in methadone maintenance treatment and buprenorphine maintenance treatment: a meta-analysis study | Prevalence was not the primary outcome |
| Yon, Yongjie | Elder abuse prevalence in community settings: a systematic review and meta-analysis | Did not assess a disease, symptom, or lifestyle factor |
| Yon, Yongjie | The Prevalence of Self-Reported Elder Abuse Among Older Women in Community Settings: A Systematic Review and Meta-Analysis | Did not assess a disease, symptom, or lifestyle factor |
| Zhao, Jian | Maternal education and breastfeeding practices in China: A systematic review and meta-analysis | Did not assess a disease, symptom, or lifestyle factor |
| Zheng, Zhen | Association between Asthma and Autism Spectrum Disorder: A Meta-Analysis | Wrong population |
| Zhu, Bifan | Disease burden of COPD in China: a systematic review | Prevalence was not the primary outcome |
| Sumanac | Is there a relationship between H. pylori prevalence and overweight? A systematic review of observational studies | Congress abstract |
| Alexander C. Ford, | Effect of validation status and type of dyspepsia questionnaire on the prevalence of dyspepsia: Systematic review and meta-analysis | Prevalence was not the primary outcome |

| | | |
|---|---|---|
| Al Nasser, Yasser | Subclinical cardiovascular changes in pediatric solid organ transplant recipients: A systematic review and meta-analysis | Wrong population |
| Asmelash, Daniel | The Burden of Undiagnosed Diabetes Mellitus in Adult African Population: A Systematic Review and Meta-Analysis | Wrong study design |
| Anandan, C. | Is the prevalence of asthma declining? Systematic review of epidemiological studies | Merged adults and children |
| Arican, I. | Prevalence of attention deficit hyperactivity disorder symptoms in patients with schizophrenia | Merged adults and children |
| Chaponda, Mas | Systematic review of the prevalence of psychiatric illness and sleep disturbance as co-morbidities of HIV infection in the UK | Congress abstract |
| Crasto, Gray, Srinivasan, Brady, Sutton, McNally, Harris, Davies | Systematic review and meta-analysis examining the prevalence of diabetic nephropathy in South Asians and white Europeans with type 2 diabetes | Congress abstract |
| Huang, J. | Prevalence of colorectal neoplasia in an average-risk Chinese population: a systematic review and meta-analysis | Congress abstract |
| Worku, Misganaw Gebrie | Prevalence and Associated Factor of Brown Adipose Tissue: Systematic Review and Meta-Analysis | Did not assess a disease, symptom, or lifestyle factor |
| A. D. S. Zanetti; A. F. Malheiros; T. A. de Matos; F. G. Longhi; L. M. Moreira; S. L. Silva; S. K. I. Castrillon; S. M. B. Ferreira; E. Ignotti; O. A. Espinosa | Prevalence of Blastocystis sp. infection in several hosts in Brazil: a systematic review and meta-analysis | Review on animal studies |
| F. Yehualashet; E. Tegegne; M. Tessema; M. Endeshaw | Human immunodeficiency virus positive status disclosure to a sexual partner and its determinant factors in Ethiopia: a systematic review and meta-analysis | Did not assess a disease, symptom, or lifestyle factor |
| Y. Xia; Q. Wu; H. Wang; S. Zhang; Y. Jiang; T. Gong; X. Xu; Q. Chang; K. Niu; Y. Zhao | Global, regional and national burden of gout, 1990–2017: a systematic analysis of the Global Burden of Disease Study | Wrong study design |
| N. Tanko; R. O. Bolaji; A. T. Olayinka; B. O. Olayinka | A systematic review on the prevalence of extended spectrum beta lactamase producing gram-negative bacteria in Nigeria | Did not assess a disease, symptom, or lifestyle factor |
| G. Sulis; P. Adam; V. Nafade; G. Gore; B. Daniels; A. Daftary; J. Das; S. Gandra; M. Pai | Antibiotic prescription practices in primary care in low- and middle-income countries: A systematic review and meta-analysis | Did not assess a disease, symptom, or lifestyle factor |

| | | |
|---|---|---|
| A. Stulz; K. Lamore; L. Montalescot; N. Favez; C. Flahault | Sexual health in colon cancer patients: A systematic review | Prevalence was not the primary outcome |
| Z. Saleem; M. A. Hassali; B. Godman; A. Versporten; F. K. Hashmi; H. Saeed; F. Saleem; M. Salman; I. U. Rehman; T. M. Khan | Point prevalence surveys of antimicrobial use: a systematic review and the implications | Did not assess a disease, symptom, or lifestyle factor |
| R. S. Poudel; S. Shrestha | Underestimation of the Prevalence of Medication Errors in Nursing Homes.. | Wrong study design |
| Peitzmeier | Intimate Partner Violence in Transgender Populations: Systematic Review and Meta-analysis of Prevalence and Correlates | Did not assess a disease, symptom, or lifestyle factor |
| Palamuthusingam, D. | Postoperative non-fatal outcomes in people receiving chronic dialysis: A systematic review and meta-analysis of 42 studies and 78,805 patients | Congress abstract |
| R. Y. K. Oliphant; E. M. Smith; V. Grahame | What is the Prevalence of Self-harming and Suicidal Behaviour in Under 18s with ASD, With or Without an Intellectual Disability? | Wrong population |
| W. A. Mason; | A systematic review of research on adolescent solitary alcohol and marijuana use in the United States | Wrong population |
| K. F. Muchie; | Epidemiology of preterm birth in Ethiopia: systematic review and meta-analysis | Wrong population |
| R. M. Mogire | Prevalence of vitamin D deficiency in Africa: a systematic review and meta-analysis | Merged adults and children |
| J. N. R. Martins; D. Marques; E. J. N. Leal Silva; J. Carames; A. Mata; M. A. Versiani | Influence of Demographic Factors on the Prevalence of a Second Root Canal in Mandibular Anterior Teeth - A Systematic Review and Meta-Analysis of Cross-Sectional Studies Using Cone Beam Computed Tomography | Did not assess a disease, symptom, or lifestyle factor |
| J. M. Lurie; A. Weidman; S. Huynh; D. Delgado; I. Easthausen; G. Kaur | Painful gynecologic and obstetric complications of female genital mutilation/cutting: A systematic review and meta-analysis | Prevalence was not the primary outcome |
| A. Licari; M. Votto; L. Scudeller; A. De Silvestri; C. Rebuffi; A. Cianferoni; G. L. Marseglia | Epidemiology of Nonesophageal Eosinophilic Gastrointestinal Diseases in Symptomatic Patients: A Systematic Review and Meta-Analysis | Merged adults and children |
| Samantha Josephine Judina Mallett , Ronald Fraser | Condom associated erection problems (CAEP) in heterosexual young men (under 40): A systematic review and qualitative synthesis | Congress abstract |

| | | |
|---|---|---|
| F. Khoshhal; H. Hashemi; E. Hooshmand; M. Saatchi; A. Yekta; M. Aghamirsalim; H. Ostadimoghaddam; M. Khabazkhoob | The prevalence of refractive errors in the Middle East: a systematic review and meta-analysis | Merged adults and children |
| A. Howren; D. Bowie; H. K. Choi; S. K. Rai; M. A. De Vera | Epidemiology of depression and anxiety in gout: A systematic review and metaanalysis | Mixed incidence and prevalence data |
| Q. L. Gong; D. Li; N. C. Diao; Y. Liu; B. Y. Li; T. Tian; G. Y. Ge; B. Zhao; Y. H. Song; D. L. Li; X. Leng; R. Du | Mink Aleutian disease seroprevalence in China during 1981-2017: A systematic review and meta-analysis | Review on animal studies |
| N. A. Deebel; G. Galdon; N. P. Zarandi; K. Stogner-Underwood; S. Howards; J. Lovato; S. Kogan; A. Atala; Y. Lue; H. Sadri-Ardekani | Age-related presence of spermatogonia in patients with Klinefelter syndrome: a systematic review and meta-analysis | Merged adults and children |
| C. M. Daniel; L. Davila; U. E. Makris; H. Mayo; L. Caplan; L. Davis; E. B. Solow | Ethnic Disparities in Atherosclerotic Cardiovascular Disease Incidence and Prevalence Among Rheumatoid Arthritis Patients in the United States: a Systematic Review | Prevalence was not the primary outcome |
| C. Crump | Preterm birth and mortality in adulthood: a systematic review | Prevalence was not the primary outcome |
| J. M. Cénat; S.-E. McIntee; C. Blais-Rochette | Symptoms of posttraumatic stress disorder, depression, anxiety and other mental health problems following the 2010 earthquake in Haiti: A systematic review and meta-analysis | Merged adults and children |
| A. Astarita; M. Covella; F. Vallelonga; M. Cesareo; S. Totaro; L. Ventre; F. Apraà; F. Veglio; A. Milan; F. Aprà | Hypertensive emergencies and urgencies in emergency departments: a systematic review and meta-analysis | Congress abstract |
| D. D. Barth; A. Moloi; B. M. Mayosi; M. E. Engel | Prevalence of group A Streptococcal infection in Africa to inform GAS vaccines for rheumatic heart disease: A systematic review and meta-analysis | Merged adults and children |
| Kolkhir P, Metz M, Altrichter S, Maurer M. | omorbidity of chronic spontaneous urticaria and autoimmune thyroid diseases: A systematic review. | Merged adults and children |
| Mansfield, Michael | Cervical spine radiculopathy epidemiology: A systematic review | Prevalence was not the primary outcome |
| Pintar KD, Christidis T, Thomas MK, | A Systematic Review and Meta-Analysis of the Campylobacter spp. | Review on animal studies |

| | | |
|---|---|---|
| Anderson M, Nesbitt A, Keithlin J, Marshall B, Pollari F. A | Prevalence and Concentration in Household Pets and Petting Zoo Animals for Use in Exposure Assessments | |
| Y. F. A. Alshehri; J. S. Park; E. Kruger; M. Tennant | Association between body mass index and dental caries in the Kingdom of Saudi Arabia: Systematic review | Prevalence was not the primary outcome |
| W. G. Alemu; T. A. Zeleke; W. W. Takele; S. S. Mekonnen | Prevalence and risk factors for khat use among youth students in Ethiopia: systematic review and meta-analysis, 2018 | Merged adults and children |
| G. P. Aguiar; M. D. Saraiva; E. J. B. Khazaal; D. C. de Andrade; W. Jacob-Filho; C. K. Suemoto | Persistent pain and cognitive decline in older adults: a systematic review and meta-analysis from longitudinal studies | Prevalence was not the primary outcome |
| J. J. Feddema and Claassen, E | Prevalence of viral respiratory infections amongst asthmatics: Results of a meta-regression analysis | Wrong study design |
| Saeid Safiri, Kolahi, Ali-Asghar, Cross, Marita, Carson-Chahhoud, Kristin, Hoy, Damian, Almasi-Hashiani, Amir, Sepidarkish, Mahdi, Ashrafi-Asgarabad, Ahad, Moradi-Lakeh, Maziar, Mansournia, Mohammad Ali, Kaufman, Jay S., Collins, Gary, Woolf, Anthony D., March, Lyn and Smith, Emma | Prevalence, Incidence, and Years Lived With Disability Due to Gout and Its Attributable Risk Factors for 195 Countries and Territories 1990–2017: A Systematic Analysis of the Global Burden of Disease Study 2017 | Wrong study design |
| Zhang Xinyue, Lizhen, Wang, Yanlin, Zheng, Lu, Deng, Xiaoying, Huang, Zhang, Xinyue, Wang, Lizhen, Zheng, Yanlin, Deng, Lu and Huang, Xiaoying | Prevalence of dry eye disease in the elderly: A protocol of systematic review and meta-analysis | Protocol for a review |
| Clarence S. Yah, Ndlovu, Sithembiso, Kutywayo, Alison, Naidoo, Nicolette, Mahuma, Tshepo and Mullick, Saiqa | The prevalence of pregnancy among adolescent girls and young women across the Southern African development community economic hub: A systematic review and meta-analysis | Merged adults and children |
| J. J. Young, Hartvigsen, J., Jensen, R. K., Roos, E. M., Ammendolia, C. and Juhl, C. B. | Prevalence of multimorbid degenerative lumbar spinal stenosis with knee and/or hip osteoarthritis: protocol for a systematic review and meta-analysis | Protocol for a review |
| Aranda-Flores CE. | Infección por el virus del papiloma humano en varones | |

| | | |
|---|---|---|
| | [Infection with human papillomavirus in men | |
| S. Zhang, Lin, X., Liu, J., Pan, Y., Zeng, X., Chen, F. and Wu, J. | Prevalence of childhood trauma measured by the short form of the Childhood Trauma Questionnaire in people with substance use disorder: A meta-analysis | Wrong study design |
| Zhang, Simei<br>Lin, Xiujin<br>Yang, Tingyu<br>Zhang, Shengjie<br>Pan, Yuli<br>Lu, Jianping<br>Liu, Jianbo | Prevalence of childhood trauma among adults with affective disorder using the Childhood Trauma Questionnaire: A meta-analysis | Wrong study design |
| A. B. N. Tonouhewa; R. Amagbegnon; S. P. Atchade; A. Hamidovic; A. Mercier; M. Dambrun; F. Migot-Nabias; Y. S. Savi de Tove; H. Sahibi; M. Laboudi; S. Sahidou; M. L. Darde; D. Kinde-Gazard; S. Farougou | [Seroprevalence of Toxoplasmosis among Pregnant Women in Benin: Meta-Analysis and Meta-Regression] | Wrong study design |
| L. R. Rith-Najarian; M. M. Boustani; B. F. Chorpita | A systematic review of prevention programs targeting depression, anxiety, and stress in university students | Prevalence was not the primary outcome |
| H. Bao; K. Liu; Z. Wu; X. Wang; C. Chai; T. He; W. Wang; F. Wang; Y. Peng; B. Chen; J. Jiang | Tuberculosis outbreaks among students in mainland China: a systematic review and meta-analysis | Merged adults and children |
| M. Kiiti Borges; N. Oiring de Castro Cezar; A. Silva Santos Siqueira; M. Yassuda; M. Cesari; I. Aprahamian | The Relationship between Physical Frailty and Mild Cognitive Impairment in the Elderly: A Systematic Review | Prevalence was not the primary outcome |
| J. Huang; T. W. Y. Pang; C. Leung; H. Ding; J. Wang; Y. Jin; Z. J. Zheng; M. C. S. Wong | Prevalence of colorectal neoplasia in an average-risk Chinese population: a systematic review and meta-analysis | Congress abstract |
| U. Basharat; M. M. Aiche; M. M. Kim; M. Sohal; E. H. Chang | A Systematic Review on the Association between Rhinovirus and Sinusitis | Congress abstract |
| Candyce Hamel, Ghannad, Mona, McInnes, Matthew D. F., Marshall, John, Earnshaw, Jonothan, Ward, Roxanne, Skidmore, Becky and Garritty, Chantelle | Potential benefits and harms of offering ultrasound surveillance to men aged 65 years and older with a subaneurysmal (2.5-2.9 cm) infrarenal aorta | Wrong study design |

| | | |
|---|---|---|
| Q. Xia, Wang, T., Xian, J., Song, J., Qiao, Y., Mu, Z., Liu, H. and Sun, Z. | Relation of Chlamydia trachomatis infections to ectopic pregnancy: A meta-analysis and systematic review | Prevalence was not the primary outcome |
| L. Mowszowski, Lampit, A., Walton, C. C. and Naismith, S. L. | Strategy-Based Cognitive Training for Improving Executive Functions in Older Adults: a Systematic Review | Prevalence was not the primary outcome |
| Neslihan Keser Özcan, Günaydın, Sevil and Çitil, Elif Tuğçe | Domestic Violence Against Women In Turkey: A Systematic Review And Meta Analysis | Did not assess a disease, symptom, or lifestyle factor |
| M. A. P. Vilela, Sbruzzi, G. and Pellanda, L. C. | Prevalence of ophthalmological abnormalities in children and adolescents with CHD: Systematic review and meta-analysis of observational studies | Wrong population |
| Menati W, Valizadeh R, Menati R, Niazi M, Nazarzadeh M, Bidel Z. | Determination of opium abuse prevalence in Iranian young people: a systematic review and meta-analysis | Merged adults and children |
| Tracy Jackson, Thomas, Sarah, Stabile, Victoria, Han, Xue, Shotwell, Matthew and McQueen, Kelly | Prevalence of chronic pain in low-income and middle-income countries: a systematic review and meta-analysis | Congress abstract |
| D. Seitz, Sherman, C. and Kirkham, J. | Prevalence of psychotropic medication use among older adults with dementia: Meta-analysis | Congress abstract |
| Kristyn Zajac, Kennedy, Caitlin E., Fonner, Virginia A., Armstrong, Kevin S., O'Reilly, Kevin R. and Sweat, Michael D. | A Systematic Review of the Effects of Behavioral Counseling on Sexual Risk Behaviors and HIV/STI Prevalence in Low- and Middle-Income Countries | Merged adults and children |
| Milad Nazarzadeh, Bidel, Zeinab, Ayubi, Erfan, Bahrami, Abolfazl, Jafari, Fatemeh, Mohammadpoorasl, Asghar, Delpisheh, Ali and Taremian, Farhad | Smoking status in Iranian male adolescents: a cross-sectional study and a meta-analysis | Wrong population |
| S. Oram, Trevillion, K., Feder, G. and Howard, L. M. | Prevalence of experiences of domestic violence among psychiatric patients: systematic review | Did not assess a disease, symptom, or lifestyle factor |
| Emilie Sonne-Holm, Wong, Christian and Sonne-Holm, Stig | Multiple cartilaginous exostoses and development of chondrosarcomas--a systematic review | Prevalence was not the primary outcome |
| Sylvie D. Lambert, Harrison, James D., Smith, Ellen, Bonevski, Billie, Carey, Mariko, Lawsin, Catalina, Paul, Chris and Girgis, Afaf | The unmet needs of partners and caregivers of adults diagnosed with cancer: a systematic review | Did not assess a disease, symptom, or lifestyle factor |

| | | |
|---|---|---|
| Emily F. Rothman, Exner, Deinera and Baughman, Allyson L. | The prevalence of sexual assault against people who identify as gay, lesbian, or bisexual in the United States: a systematic review | Did not assess a disease, symptom, or lifestyle factor |
| Vlak MH, Algra A, Brandenburg R, Rinkel GJ. | Prevalence of unruptured intracranial aneurysms: An updated systematic review with emphasis on sex, age, country, indication for study and time-trend | Congress abstract |
| A. Van Lancker, Velghe, A., Verbrugghe, M., Bekkering, G. E. and Beeckman, D. | Prevalence of symptoms in the elderly with incurable cancer: a systematic review | Protocol for a review |
| Y. Zhang, L. He and N. Chen | Prevalences of migraine and other types of primary headache in China: A systematic review and meta-analysis | Congress abstract |
| Stewart J. Jackson, Steer, Andrew C. and Campbell, Harry | Systematic Review: Estimation of global burden of non-suppurative sequelae of upper respiratory tract infection: rheumatic fever and post-streptococcal glomerulonephritis | Wrong population |
| Gray Srinivasan Brady Sutton McNally Harris Davies Crasto | Systematic review and meta-analysis examining the prevalence of diabetic nephropathy in South Asians and white Europeans with type 2 diabetes | Congress abstract |
| D. G. Hoy, March, L., Brooks, P., Woolf, A., Blyth, F., Smith, E., Vos, T. and Buchbinder, R. | The prevalence of low back pain throughout the world | Congress abstract |
| A. Murphy, Pogosova, N., Anderson, L., Goldman, L., Mensah, G. and Moran, A. | Epidemiology of ischemic heart disease in Eastern Europe from 1985 to 2008: A systematic review for the Global Burden of Disease study | Congress abstract |
| Posadzki P, Alotaibi A, Ernst E. | Prevalence of use of complementary and alternative medicine (CAM) by physicians in the UK: a systematic review of surveys | Did not assess a disease, symptom, or lifestyle factor |
| Operario D, Underhill K, Chuong C, Cluver L. | HIV infection and sexual risk behaviour among youth who have experienced orphanhood: systematic review and meta-analysis | Merged adults and children |
| Wang X, Wang W. | Prevalence of Bicuspid Aortic Valve in Chinese Patients with Aortic Valve Disease: A Systematic Review. | Full text not available |
| Fereshteh Dastouri, Hosseini, Ahmad Mirmohammad, Haworth, Elizabeth, Khandaker, Gulam, Rashid, Harunor and Booy, Robert | Complications of serogroup B meningococcal disease in survivors: a review | Full text not available |

| | | |
|---|---|---|
| Dastouri F, Hosseini AM, Haworth E, Khandaker G, Rashid H, Booy R. | Complications of serogroup B meningococcal disease in survivors: a review. Infect Disord Drug Targets | Full text not available |
| Hasan DM, Emeash AH, Mustafa SB, Abdelazim GEA, El-din AA | Hypertension in Egypt: a systematic review. | Full text not available |
| He Y, Sun LY, Gong R, Liu Q, Long YK, Liu F, et al. | The prevalence of EML4-ALK variants in patients with non-small-cell lung cancer: A systematic review and meta-analysis. | Full text not available |
| Vilela MAP, Sbruzzi G, Pellanda LC. | Prevalence of ophthalmological abnormalities in children and adolescents with CHD: Systematic review and meta-analysis of observational studies. | Full text not available |
| Yang X, Li X, Han J, Xu L, Zou L, Wang J, et al. | Prevalence of methicillin-resistant staphylococcus aureus in healthy population in China: A meta-analysis. Chinese Journal of Evidence Based Medicine. | Full text not available |
| S. J. J. Mallett; R. Frase | Condom associated erection problems (CAEP) in heterosexual young men (under 40): A systematic review and qualitative synthesi | Full text not available |
| Rafiq SM, Banik GR, Khan S, Rashid H, Khandaker G. | Current Burden of Hepatitis C Virus Infection Among Injecting Drug Users: A Mini Systematic Review of Prevalence Studies | Full text not available |
| Savi L, Ribaldone DG, Fagoonee S, Pellicano R. | Is Helicobacter pylori the infectious trigger for headache?: A review. | Full text not available |
