## Supplementary material for "Characteristics and completeness of reporting of systematic reviews of prevalence studies in adult populations: a meta-research study": Supp_tables

### Supplementary tables and figures

**Figure S1.** Geographic heat map of the first author reported affiliation in systematic reviews of prevalence in adults 2010-2020 ........ 20
**Figure S2.** Specialty or subject of systematic reviews of prevalence in adult populations published 2010-2020 ........ 23

**Table S1**.Journals that published systematic reviews of prevalence in adults 2010-2020

| Journal | Systematic reviews without meta-analysis n=387* | Systematic reviews with meta-analysis n=785* | Total, n=1172* |
|---|---|---|---|
| PloS one | 11 (2.8%) | 35 (4.5%) | 46 (3.9%) |
| BMC Public Health | 5 (1.3%) | 18 (2.3%) | 23 (2.0%) |
| BMC Infectious Diseases | 4 (1.0%) | 16 (2.0%) | 20 (1.7%) |
| Journal of Affective Disorders | 4 (1.0%) | 13 (1.7%) | 17 (1.5%) |
| BMJ Open | 1 (0.3%) | 10 (1.3%) | 11 (0.9%) |
| Age and Ageing | 2 (0.5%) | 7 (0.9%) | 9 (0.8%) |
| Tropical Medicine & International Health | 3 (0.8%) | 6 (0.8%) | 9 (0.8%) |
| Alimentary Pharmacology & Therapeutics | 1 (0.3%) | 7 (0.9%) | 8 (0.7%) |
| Clinical Gastroenterology and Hepatology | 0 (0%) | 8 (1.0%) | 8 (0.7%) |
| Medicine | 1 (0.3%) | 7 (0.9%) | 8 (0.7%) |
| British Journal of Ophthalmology | 3 (0.8%) | 4 (0.5%) | 7 (0.6%) |
| Journal of the American Medical Directors Association | 2 (0.5%) | 5 (0.6%) | 7 (0.6%) |
| Rheumatology | 2 (0.5%) | 5 (0.6%) | 7 (0.6%) |
| International Journal of Environmental Research and Public Health | 3 (0.8%) | 3 (0.4%) | 6 (0.5%) |
| Journal of Hypertension | 2 (0.5%) | 4 (0.5%) | 6 (0.5%) |
| Maturitas | 3 (0.8%) | 3 (0.4%) | 6 (0.5%) |
| Nutrients | 0 (0%) | 6 (0.8%) | 6 (0.5%) |
| PLoS medicine | 1 (0.3%) | 5 (0.6%) | 6 (0.5%) |
| Revista Ciência & Saúde Coletiva | 3 (0.8%) | 3 (0.4%) | 6 (0.5%) |
| Sexually Transmitted Infections | 1 (0.3%) | 5 (0.6%) | 6 (0.5%) |
| The Lancet Infectious Diseases | 2 (0.5%) | 4 (0.5%) | 6 (0.5%) |
| Acta Psychiatrica Scandinavica | 2 (0.5%) | 3 (0.4%) | 5 (0.4%) |
| Cadernos de Saúde Pública | 3 (0.8%) | 2 (0.3%) | 5 (0.4%) |
| Climacteric | 4 (1.0%) | 1 (0.1%) | 5 (0.4%) |
| Critical Care Medicine | 0 (0%) | 5 (0.6%) | 5 (0.4%) |
| Diabetes Research and Clinical Practice | 3 (0.8%) | 2 (0.3%) | 5 (0.4%) |
| International Journal of STD & AIDS | 1 (0.3%) | 4 (0.5%) | 5 (0.4%) |
| Revista de Saúde Pública | 4 (1.0%) | 1 (0.1%) | 5 (0.4%) |
| Sleep and Breathing | 1 (0.3%) | 4 (0.5%) | 5 (0.4%) |
| The Journal of Allergy and Clinical Immunology | 2 (0.5%) | 3 (0.4%) | 5 (0.4%) |
| The Lancet | 3 (0.8%) | 2 (0.3%) | 5 (0.4%) |
| Addiction | 0 (0%) | 4 (0.5%) | 4 (0.3%) |
| American Journal of Gastroenterology | 0 (0%) | 4 (0.5%) | 4 (0.3%) |
| Asian Pacific Journal of Cancer Prevention | 2 (0.5%) | 2 (0.3%) | 4 (0.3%) |
| BioMed Research International | 2 (0.5%) | 2 (0.3%) | 4 (0.3%) |
| BMC Musculoskeletal Disorders | 2 (0.5%) | 2 (0.3%) | 4 (0.3%) |

| **Journal** | **Systematic reviews without meta-analysis n=387*** | **Systematic reviews with meta-analysis n=785*** | **Total**, n=1172* |
|---|---|---|---|
| British Journal of Dermatology | 0 (0%) | 4 (0.5%) | 4 (0.3%) |
| British Journal of Psychiatry | 0 (0%) | 4 (0.5%) | 4 (0.3%) |
| Bulletin of the World Health Organization | 2 (0.5%) | 2 (0.3%) | 4 (0.3%) |
| General Hospital Psychiatry | 2 (0.5%) | 2 (0.3%) | 4 (0.3%) |
| International Journal of Cardiology | 1 (0.3%) | 3 (0.4%) | 4 (0.3%) |
| International Psychogeriatrics | 4 (1.0%) | 0 (0%) | 4 (0.3%) |
| Journal of Global Health | 0 (0%) | 4 (0.5%) | 4 (0.3%) |
| Journal of Stroke and Cerebrovascular Diseases | 0 (0%) | 4 (0.5%) | 4 (0.3%) |
| Journal of the American Geriatrics Society | 4 (1.0%) | 0 (0%) | 4 (0.3%) |
| Ophthalmology | 0 (0%) | 4 (0.5%) | 4 (0.3%) |
| Oral Diseases | 0 (0%) | 4 (0.5%) | 4 (0.3%) |
| Palliative Medicine | 2 (0.5%) | 2 (0.3%) | 4 (0.3%) |
| PLoS Neglected Tropical Diseases | 0 (0%) | 4 (0.5%) | 4 (0.3%) |
| Psychiatry Research | 1 (0.3%) | 3 (0.4%) | 4 (0.3%) |
| Public Health | 2 (0.5%) | 2 (0.3%) | 4 (0.3%) |
| Revista Da Escola de Enfermagem | 1 (0.3%) | 3 (0.4%) | 4 (0.3%) |
| Revista Panamericana de Salud Pública | 0 (0%) | 4 (0.5%) | 4 (0.3%) |
| Rheumatology International | 2 (0.5%) | 2 (0.3%) | 4 (0.3%) |
| Scientific Reports | 0 (0%) | 4 (0.5%) | 4 (0.3%) |
| Seminars in Arthritis & Rheumatism | 0 (0%) | 4 (0.5%) | 4 (0.3%) |
| Sexual Health | 0 (0%) | 4 (0.5%) | 4 (0.3%) |
| Sports Medicine | 3 (0.8%) | 1 (0.1%) | 4 (0.3%) |
| The Lancet Global Health | 0 (0%) | 4 (0.5%) | 4 (0.3%) |
| AIDS and Behavior | 0 (0%) | 3 (0.4%) | 3 (0.3%) |
| American Journal of Public Health | 1 (0.3%) | 2 (0.3%) | 3 (0.3%) |
| Annals of Hematology | 1 (0.3%) | 2 (0.3%) | 3 (0.3%) |
| Annals of Oncology | 2 (0.5%) | 1 (0.1%) | 3 (0.3%) |
| Archives of Women's Mental Health | 0 (0%) | 3 (0.4%) | 3 (0.3%) |
| Asian Journal of Psychiatry | 2 (0.5%) | 1 (0.1%) | 3 (0.3%) |
| Australian and New Zealand Journal of Psychiatry | 1 (0.3%) | 2 (0.3%) | 3 (0.3%) |
| BMC Cancer | 1 (0.3%) | 2 (0.3%) | 3 (0.3%) |
| BMC Geriatrics | 1 (0.3%) | 2 (0.3%) | 3 (0.3%) |
| BMC Psychiatry | 1 (0.3%) | 2 (0.3%) | 3 (0.3%) |
| BMC Pulmonary Medicine | 1 (0.3%) | 2 (0.3%) | 3 (0.3%) |
| BMC Research Notes | 2 (0.5%) | 1 (0.1%) | 3 (0.3%) |
| British Journal of Surgery | 1 (0.3%) | 2 (0.3%) | 3 (0.3%) |
| Critical Care | 0 (0%) | 3 (0.4%) | 3 (0.3%) |
| Diabetic Medicine | 2 (0.5%) | 1 (0.1%) | 3 (0.3%) |
| Drug and Alcohol Dependence | 1 (0.3%) | 2 (0.3%) | 3 (0.3%) |
| European Journal of Gastroenterology & Hepatology | 0 (0%) | 3 (0.4%) | 3 (0.3%) |

| **Journal** | **Systematic reviews without meta-analysis n=387*** | **Systematic reviews with meta-analysis n=785*** | **Total**, n=1172* |
|---|---|---|---|
| European Spine Journal | 1 (0.3%) | 2 (0.3%) | 3 (0.3%) |
| Hepatitis Monthly | 0 (0%) | 3 (0.4%) | 3 (0.3%) |
| International Journal of Chronic Obstructive Pulmonary Disease | 1 (0.3%) | 2 (0.3%) | 3 (0.3%) |
| International Journal of Geriatric Psychiatry | 2 (0.5%) | 1 (0.1%) | 3 (0.3%) |
| International Journal of Infectious Diseases | 1 (0.3%) | 2 (0.3%) | 3 (0.3%) |
| International Journal of Preventive Medicine | 2 (0.5%) | 1 (0.1%) | 3 (0.3%) |
| International Journal of Rheumatic Diseases | 2 (0.5%) | 1 (0.1%) | 3 (0.3%) |
| JAMA Network Open | 0 (0%) | 3 (0.4%) | 3 (0.3%) |
| Journal of Clinical Endocrinology and Metabolism | 0 (0%) | 3 (0.4%) | 3 (0.3%) |
| Journal of Clinical Psychiatry | 1 (0.3%) | 2 (0.3%) | 3 (0.3%) |
| Journal of Dentistry | 1 (0.3%) | 2 (0.3%) | 3 (0.3%) |
| Journal of Infectious Diseases | 1 (0.3%) | 2 (0.3%) | 3 (0.3%) |
| Journal of Pain & Symptom Management | 1 (0.3%) | 2 (0.3%) | 3 (0.3%) |
| Journal of Pain and Symptom Management | 1 (0.3%) | 2 (0.3%) | 3 (0.3%) |
| Journal of the American Academy of Dermatology | 1 (0.3%) | 2 (0.3%) | 3 (0.3%) |
| Nephrology Dialysis Transplantation | 1 (0.3%) | 2 (0.3%) | 3 (0.3%) |
| Pain | 1 (0.3%) | 2 (0.3%) | 3 (0.3%) |
| Psychological Medicine | 2 (0.5%) | 1 (0.1%) | 3 (0.3%) |
| Revista Brasileira de Psiquiatria | 0 (0%) | 3 (0.4%) | 3 (0.3%) |
| Revista Clínica Española | 0 (0%) | 3 (0.4%) | 3 (0.3%) |
| Revista da Associação Médica Brasileira | 3 (0.8%) | 0 (0%) | 3 (0.3%) |
| Revista de Salud Pública | 3 (0.8%) | 0 (0%) | 3 (0.3%) |
| Sexually Transmitted Diseases | 2 (0.5%) | 1 (0.1%) | 3 (0.3%) |
| Acta Diabetologica | 0 (0%) | 2 (0.3%) | 2 (0.2%) |
| Acta Obstetricia et Gynecologica Scandinavica | 2 (0.5%) | 0 (0%) | 2 (0.2%) |
| Acta Tropica | 0 (0%) | 2 (0.3%) | 2 (0.2%) |
| Addictive Behaviors | 2 (0.5%) | 0 (0%) | 2 (0.2%) |
| Advances in Therapy | 1 (0.3%) | 1 (0.1%) | 2 (0.2%) |
| Ageing Research Reviews | 1 (0.3%) | 1 (0.1%) | 2 (0.2%) |
| Aging and Mental Health | 1 (0.3%) | 1 (0.1%) | 2 (0.2%) |
| Alzheimer's & Dementia | 1 (0.3%) | 1 (0.1%) | 2 (0.2%) |
| American Heart Journal | 2 (0.5%) | 0 (0%) | 2 (0.2%) |
| American Journal of Geriatric Psychiatry | 1 (0.3%) | 1 (0.1%) | 2 (0.2%) |
| American Journal of Infection Control | 1 (0.3%) | 1 (0.1%) | 2 (0.2%) |
| Annals of Epidemiology | 1 (0.3%) | 1 (0.1%) | 2 (0.2%) |

| Journal | Systematic reviews without meta-analysis n=387* | Systematic reviews with meta-analysis n=785* | **Total**, n=1172* |
|---|---|---|---|
| Annals of General Psychiatry | 0 (0%) | 2 (0.3%) | 2 (0.2%) |
| Arthritis & Rheumatism | 0 (0%) | 2 (0.3%) | 2 (0.2%) |
| Arthritis Care & Research | 0 (0%) | 2 (0.3%) | 2 (0.2%) |
| Asia-Pacific Journal of Public Health | 2 (0.5%) | 0 (0%) | 2 (0.2%) |
| Autoimmunity Reviews | 1 (0.3%) | 1 (0.1%) | 2 (0.2%) |
| BMC Nephrology | 1 (0.3%) | 1 (0.1%) | 2 (0.2%) |
| BMC Neurology | 1 (0.3%) | 1 (0.1%) | 2 (0.2%) |
| BMC Nursing | 1 (0.3%) | 1 (0.1%) | 2 (0.2%) |
| BMC Pregnancy & Childbirth | 0 (0%) | 2 (0.3%) | 2 (0.2%) |
| Brain Injury | 2 (0.5%) | 0 (0%) | 2 (0.2%) |
| Brazilian Journal of Infectious Diseases | 1 (0.3%) | 1 (0.1%) | 2 (0.2%) |
| British Journal of Sports Medicine | 1 (0.3%) | 1 (0.1%) | 2 (0.2%) |
| Cadernos Brasileiros de Terapia Ocupacional | 1 (0.3%) | 1 (0.1%) | 2 (0.2%) |
| Circulation | 0 (0%) | 2 (0.3%) | 2 (0.2%) |
| Clinical & Experimental Allergy | 1 (0.3%) | 1 (0.1%) | 2 (0.2%) |
| Clinical Infectious Diseases | 0 (0%) | 2 (0.3%) | 2 (0.2%) |
| Clinical Interventions in Aging | 2 (0.5%) | 0 (0%) | 2 (0.2%) |
| Clinical Microbiology and Infection | 0 (0%) | 2 (0.3%) | 2 (0.2%) |
| Clinical Nutrition | 2 (0.5%) | 0 (0%) | 2 (0.2%) |
| Clinical Oral Investigations | 0 (0%) | 2 (0.3%) | 2 (0.2%) |
| Clinical Respiratory Journal | 1 (0.3%) | 1 (0.1%) | 2 (0.2%) |
| Clinics | 2 (0.5%) | 0 (0%) | 2 (0.2%) |
| Comprehensive Psychiatry | 2 (0.5%) | 0 (0%) | 2 (0.2%) |
| Dementia | 2 (0.5%) | 0 (0%) | 2 (0.2%) |
| Dementia and Geriatric Cognitive Disorders | 1 (0.3%) | 1 (0.1%) | 2 (0.2%) |
| Developmental Medicine and Child Neurology | 0 (0%) | 2 (0.3%) | 2 (0.2%) |
| Diabetes & Metabolic Syndrome | 0 (0%) | 2 (0.3%) | 2 (0.2%) |
| Digestive Diseases and Sciences | 0 (0%) | 2 (0.3%) | 2 (0.2%) |
| Disability and Rehabilitation | 2 (0.5%) | 0 (0%) | 2 (0.2%) |
| Drugs & Aging | 2 (0.5%) | 0 (0%) | 2 (0.2%) |
| Eastern Mediterranean Health Journal | 0 (0%) | 2 (0.3%) | 2 (0.2%) |
| Endocrine | 0 (0%) | 2 (0.3%) | 2 (0.2%) |
| Epilepsia | 1 (0.3%) | 1 (0.1%) | 2 (0.2%) |
| European Archives of Oto-Rhino-Laryngology | 2 (0.5%) | 0 (0%) | 2 (0.2%) |
| European Geriatric Medicine | 0 (0%) | 2 (0.3%) | 2 (0.2%) |
| European Heart Journal | 0 (0%) | 2 (0.3%) | 2 (0.2%) |
| European Journal of Epidemiology | 0 (0%) | 2 (0.3%) | 2 (0.2%) |
| European Urology | 1 (0.3%) | 1 (0.1%) | 2 (0.2%) |
| Eye | 1 (0.3%) | 1 (0.1%) | 2 (0.2%) |
| Frontiers in Psychiatry | 1 (0.3%) | 1 (0.1%) | 2 (0.2%) |

| **Journal** | **Systematic reviews without meta-analysis n=387*** | **Systematic reviews with meta-analysis n=785*** | **Total**, n=1172* |
|---|---|---|---|
| Gastroenterology | 0 (0%) | 2 (0.3%) | 2 (0.2%) |
| Globalization and Health | 0 (0%) | 2 (0.3%) | 2 (0.2%) |
| Gut | 0 (0%) | 2 (0.3%) | 2 (0.2%) |
| Gynecological Endocrinology | 0 (0%) | 2 (0.3%) | 2 (0.2%) |
| Human Reproduction Update | 0 (0%) | 2 (0.3%) | 2 (0.2%) |
| Infectious Diseases in Obstetrics and Gynecology | 1 (0.3%) | 1 (0.1%) | 2 (0.2%) |
| Internal and Emergency Medicine | 0 (0%) | 2 (0.3%) | 2 (0.2%) |
| Internal Medicine Journal | 2 (0.5%) | 0 (0%) | 2 (0.2%) |
| Iranian journal of Kidney Diseases | 0 (0%) | 2 (0.3%) | 2 (0.2%) |
| Iranian Journal of Psychiatry and Behavioral Sciences | 0 (0%) | 2 (0.3%) | 2 (0.2%) |
| Iranian Journal of Public Health | 0 (0%) | 2 (0.3%) | 2 (0.2%) |
| JACC Heart Failure | 0 (0%) | 2 (0.3%) | 2 (0.2%) |
| JBI Library of Systematic Reviews | 2 (0.5%) | 0 (0%) | 2 (0.2%) |
| Journal of Adolescent Health | 1 (0.3%) | 1 (0.1%) | 2 (0.2%) |
| Journal of Asthma | 0 (0%) | 2 (0.3%) | 2 (0.2%) |
| Journal of Burn Care and Research | 0 (0%) | 2 (0.3%) | 2 (0.2%) |
| Journal of Endocrinological Investigation | 0 (0%) | 2 (0.3%) | 2 (0.2%) |
| Journal of Gastrointestinal and Liver Diseases | 1 (0.3%) | 1 (0.1%) | 2 (0.2%) |
| Journal of Head Trauma Rehabilitation | 2 (0.5%) | 0 (0%) | 2 (0.2%) |
| Journal of Hepatology | 0 (0%) | 2 (0.3%) | 2 (0.2%) |
| Journal of Minimally Invasive Gynecology | 1 (0.3%) | 1 (0.1%) | 2 (0.2%) |
| Journal of Obstetrics and Gynaecology Canada | 1 (0.3%) | 1 (0.1%) | 2 (0.2%) |
| Journal of Oral Pathology & Medicine | 0 (0%) | 2 (0.3%) | 2 (0.2%) |
| Journal of Orofacial Pain | 2 (0.5%) | 0 (0%) | 2 (0.2%) |
| Journal of Orthopaedic Trauma | 0 (0%) | 2 (0.3%) | 2 (0.2%) |
| Journal of Personality Disorders | 0 (0%) | 2 (0.3%) | 2 (0.2%) |
| Journal of Psychosomatic Research | 1 (0.3%) | 1 (0.1%) | 2 (0.2%) |
| Journal of Public Health | 1 (0.3%) | 1 (0.1%) | 2 (0.2%) |
| Journal of Rheumatology | 1 (0.3%) | 1 (0.1%) | 2 (0.2%) |
| Journal of Substance Use | 0 (0%) | 2 (0.3%) | 2 (0.2%) |
| Journal of the American Heart Association | 0 (0%) | 2 (0.3%) | 2 (0.2%) |
| Journal of the International AIDS Society | 0 (0%) | 2 (0.3%) | 2 (0.2%) |
| Journal of the Neurological Sciences | 1 (0.3%) | 1 (0.1%) | 2 (0.2%) |
| Journal of Thrombosis & Haemostasis | 1 (0.3%) | 1 (0.1%) | 2 (0.2%) |
| Mayo Clinic Proceedings | 1 (0.3%) | 1 (0.1%) | 2 (0.2%) |
| Memórias do Instituto Oswaldo Cruz | 2 (0.5%) | 0 (0%) | 2 (0.2%) |
| Menopause | 1 (0.3%) | 1 (0.1%) | 2 (0.2%) |

| **Journal** | **Systematic reviews without meta-analysis n=387*** | **Systematic reviews with meta-analysis n=785*** | **Total**, n=1172* |
|---|---|---|---|
| Metabolic Syndrome & Related Disorders | 0 (0%) | 2 (0.3%) | 2 (0.2%) |
| Microbial Pathogenesis | 0 (0%) | 2 (0.3%) | 2 (0.2%) |
| Neurosurgical Review | 0 (0%) | 2 (0.3%) | 2 (0.2%) |
| Neurourology and Urodynamics | 1 (0.3%) | 1 (0.1%) | 2 (0.2%) |
| Nigerian Journal of Clinical Practice | 1 (0.3%) | 1 (0.1%) | 2 (0.2%) |
| Nutrición Hospitalaria | 2 (0.5%) | 0 (0%) | 2 (0.2%) |
| Oncotarget | 0 (0%) | 2 (0.3%) | 2 (0.2%) |
| Ophthalmic Research | 0 (0%) | 2 (0.3%) | 2 (0.2%) |
| Osteoporosis International | 0 (0%) | 2 (0.3%) | 2 (0.2%) |
| Pathogens and Global Health | 1 (0.3%) | 1 (0.1%) | 2 (0.2%) |
| Personality and Mental Health | 1 (0.3%) | 1 (0.1%) | 2 (0.2%) |
| Primary Care Diabetes | 0 (0%) | 2 (0.3%) | 2 (0.2%) |
| Public Health Nutrition | 2 (0.5%) | 0 (0%) | 2 (0.2%) |
| Revista Brasileira de Cirurgia Plástica | 1 (0.3%) | 1 (0.1%) | 2 (0.2%) |
| Revista Brasileira de Terapia Intensiva | 2 (0.5%) | 0 (0%) | 2 (0.2%) |
| Revista Cubana de Hematologia, Inmunologia y Hemoterapia | 0 (0%) | 2 (0.3%) | 2 (0.2%) |
| Revista da Sociedade Brasileira de Medicina Tropical | 0 (0%) | 2 (0.3%) | 2 (0.2%) |
| Sao Paulo Medical Journal | 0 (0%) | 2 (0.3%) | 2 (0.2%) |
| Saudi Journal of Kidney Diseases and Transplantation | 1 (0.3%) | 1 (0.1%) | 2 (0.2%) |
| Sleep Medicine | 0 (0%) | 2 (0.3%) | 2 (0.2%) |
| Sleep Medicine Reviews | 1 (0.3%) | 1 (0.1%) | 2 (0.2%) |
| Substance Abuse Treatment, Prevention and Policy | 0 (0%) | 2 (0.3%) | 2 (0.2%) |
| The Lancet Gastroenterology and Hepatology | 0 (0%) | 2 (0.3%) | 2 (0.2%) |
| The Lancet Psychiatry | 1 (0.3%) | 1 (0.1%) | 2 (0.2%) |
| Turk Kardiyoloji Dernegi Arsivi | 0 (0%) | 2 (0.3%) | 2 (0.2%) |
| World Journal of Gastroenterology | 2 (0.5%) | 0 (0%) | 2 (0.2%) |
| Zoonoses and Public Health | 0 (0%) | 2 (0.3%) | 2 (0.2%) |
| Journal of the European Academy of Dermatology and Venereology | 1 (0.3%) | 0 (0%) | 1 (<0.1%) |
| Abdominal Radiology | 0 (0%) | 1 (0.1%) | 1 (<0.1%) |
| Academic Radiology | 0 (0%) | 1 (0.1%) | 1 (<0.1%) |
| Acta Cardiologica Sinica | 0 (0%) | 1 (0.1%) | 1 (<0.1%) |
| Acta Fisiátrica | 1 (0.3%) | 0 (0%) | 1 (<0.1%) |
| Acta Medica Iranica | 0 (0%) | 1 (0.1%) | 1 (<0.1%) |
| Acta Odontologica Scandinavica | 0 (0%) | 1 (0.1%) | 1 (<0.1%) |
| Addictive Disorders and Their Treatment | 1 (0.3%) | 0 (0%) | 1 (<0.1%) |
| Advances in Nutrition | 1 (0.3%) | 0 (0%) | 1 (<0.1%) |
| African Health Sciences | 0 (0%) | 1 (0.1%) | 1 (<0.1%) |

| Journal | Systematic reviews without meta-analysis n=387* | Systematic reviews with meta-analysis n=785* | Total, n=1172* |
|---|---|---|---|
| African Journal of Emergency Medicine | 0 (0%) | 1 (0.1%) | 1 (<0.1%) |
| African Journal of Reproductive Health | 1 (0.3%) | 0 (0%) | 1 (<0.1%) |
| Aging and Disease | 1 (0.3%) | 0 (0%) | 1 (<0.1%) |
| AIDS Care | 0 (0%) | 1 (0.1%) | 1 (<0.1%) |
| AIDS Patient Care and Stds | 1 (0.3%) | 0 (0%) | 1 (<0.1%) |
| AIDS Reviews | 0 (0%) | 1 (0.1%) | 1 (<0.1%) |
| Air Medical Journal | 1 (0.3%) | 0 (0%) | 1 (<0.1%) |
| Alcohol and Alcoholism | 0 (0%) | 1 (0.1%) | 1 (<0.1%) |
| Alzheimer Disease and Associated Disorders | 1 (0.3%) | 0 (0%) | 1 (<0.1%) |
| American Journal of Clinical Pathology | 0 (0%) | 1 (0.1%) | 1 (<0.1%) |
| American Journal of Hypertension | 0 (0%) | 1 (0.1%) | 1 (<0.1%) |
| American Journal of Men's Health | 0 (0%) | 1 (0.1%) | 1 (<0.1%) |
| American Journal of Nephrology | 0 (0%) | 1 (0.1%) | 1 (<0.1%) |
| American Journal of Neuroradiology | 1 (0.3%) | 0 (0%) | 1 (<0.1%) |
| American Journal of Respiratory and Critical care medicine | 1 (0.3%) | 0 (0%) | 1 (<0.1%) |
| American Journal of Sports Medicine | 1 (0.3%) | 0 (0%) | 1 (<0.1%) |
| American Journal of Transplantation | 0 (0%) | 1 (0.1%) | 1 (<0.1%) |
| American Journal of Tropical Medicine and Hygiene | 0 (0%) | 1 (0.1%) | 1 (<0.1%) |
| Anais Brasileiros de Dermatologia | 1 (0.3%) | 0 (0%) | 1 (<0.1%) |
| Annali dell'Istituto Superiore di Sanita | 0 (0%) | 1 (0.1%) | 1 (<0.1%) |
| Annals of Allergy, Asthma, & Immunology | 0 (0%) | 1 (0.1%) | 1 (<0.1%) |
| Annals of Clinical Microbiology and Antimicrobials | 0 (0%) | 1 (0.1%) | 1 (<0.1%) |
| Annals of Medicine | 0 (0%) | 1 (0.1%) | 1 (<0.1%) |
| Annals of Physical and Rehabilitation Medicine | 1 (0.3%) | 0 (0%) | 1 (<0.1%) |
| Annals of Saudi medicine | 0 (0%) | 1 (0.1%) | 1 (<0.1%) |
| Antimicrobial Resistance & Infection Control | 0 (0%) | 1 (0.1%) | 1 (<0.1%) |
| Antiviral Therapy | 1 (0.3%) | 0 (0%) | 1 (<0.1%) |
| Archives of cardiovascular diseases | 0 (0%) | 1 (0.1%) | 1 (<0.1%) |
| Archives of endocrinology and metabolism | 1 (0.3%) | 0 (0%) | 1 (<0.1%) |
| Archives of Environmental & Occupational Health | 1 (0.3%) | 0 (0%) | 1 (<0.1%) |
| Archives of Gerontology and Geriatrics | 1 (0.3%) | 0 (0%) | 1 (<0.1%) |
| Archives of Gynecology and Obstetrics | 1 (0.3%) | 0 (0%) | 1 (<0.1%) |
| Archives of Iranian medicine | 1 (0.3%) | 0 (0%) | 1 (<0.1%) |
| Archives of Orthopaedic and Trauma Surgery | 1 (0.3%) | 0 (0%) | 1 (<0.1%) |
| Archives of Osteoporosis | 0 (0%) | 1 (0.1%) | 1 (<0.1%) |

| **Journal** | **Systematic reviews without meta-analysis n=387*** | **Systematic reviews with meta-analysis n=785*** | **Total**, n=1172* |
|---|---|---|---|
| Archives of Public Health | 0 (0%) | 1 (0.1%) | 1 (<0.1%) |
| Archives of Rehabilitation | 1 (0.3%) | 0 (0%) | 1 (<0.1%) |
| Archives of Rheumatology | 0 (0%) | 1 (0.1%) | 1 (<0.1%) |
| Archives of Sexual Behavior | 0 (0%) | 1 (0.1%) | 1 (<0.1%) |
| Archives of Virology | 0 (0%) | 1 (0.1%) | 1 (<0.1%) |
| Arthritis Research & Therapy | 0 (0%) | 1 (0.1%) | 1 (<0.1%) |
| Arthroscopy | 1 (0.3%) | 0 (0%) | 1 (<0.1%) |
| ARYA Atherosclerosis | 0 (0%) | 1 (0.1%) | 1 (<0.1%) |
| Asia-Pacific Psychiatry | 1 (0.3%) | 0 (0%) | 1 (<0.1%) |
| Asian Biomedicine | 0 (0%) | 1 (0.1%) | 1 (<0.1%) |
| Asian journal of surgery | 0 (0%) | 1 (0.1%) | 1 (<0.1%) |
| Atherosclerosis | 0 (0%) | 1 (0.1%) | 1 (<0.1%) |
| Australian Critical Care | 0 (0%) | 1 (0.1%) | 1 (<0.1%) |
| Biomarkers in Medicine | 0 (0%) | 1 (0.1%) | 1 (<0.1%) |
| Blood Pressure | 1 (0.3%) | 0 (0%) | 1 (<0.1%) |
| BMC Cardiovascular Disorders | 0 (0%) | 1 (0.1%) | 1 (<0.1%) |
| BMC Dermatology | 0 (0%) | 1 (0.1%) | 1 (<0.1%) |
| BMC Gastroenterology | 0 (0%) | 1 (0.1%) | 1 (<0.1%) |
| BMC Hematology | 0 (0%) | 1 (0.1%) | 1 (<0.1%) |
| BMC Medicine | 0 (0%) | 1 (0.1%) | 1 (<0.1%) |
| BMC Urology | 0 (0%) | 1 (0.1%) | 1 (<0.1%) |
| BMC Women's Health | 0 (0%) | 1 (0.1%) | 1 (<0.1%) |
| BMJ Open Sport & Exercise Medicine | 0 (0%) | 1 (0.1%) | 1 (<0.1%) |
| BMJ Supportive & Palliative Care | 0 (0%) | 1 (0.1%) | 1 (<0.1%) |
| Body Image | 0 (0%) | 1 (0.1%) | 1 (<0.1%) |
| Brain and Behavior | 1 (0.3%) | 0 (0%) | 1 (<0.1%) |
| Brain, Behavior, and Immunity | 0 (0%) | 1 (0.1%) | 1 (<0.1%) |
| Brazilian Journal of Nephrology | 0 (0%) | 1 (0.1%) | 1 (<0.1%) |
| British Journal of Anaesthesia | 0 (0%) | 1 (0.1%) | 1 (<0.1%) |
| British Journal of Clinical Pharmacology | 1 (0.3%) | 0 (0%) | 1 (<0.1%) |
| British Journal of Learning Disabilities | 1 (0.3%) | 0 (0%) | 1 (<0.1%) |
| Burns | 1 (0.3%) | 0 (0%) | 1 (<0.1%) |
| Calcified Tissue International | 0 (0%) | 1 (0.1%) | 1 (<0.1%) |
| Canadian Journal of Cardiology | 0 (0%) | 1 (0.1%) | 1 (<0.1%) |
| Canadian Journal of Psychiatry | 0 (0%) | 1 (0.1%) | 1 (<0.1%) |
| Canadian Respiratory Journal | 0 (0%) | 1 (0.1%) | 1 (<0.1%) |
| Cancer Treatment Reviews | 0 (0%) | 1 (0.1%) | 1 (<0.1%) |
| Cardiology Research and Practice | 0 (0%) | 1 (0.1%) | 1 (<0.1%) |
| Cardiovascular Diabetology | 1 (0.3%) | 0 (0%) | 1 (<0.1%) |
| Caspian Journal of Internal Medicine | 1 (0.3%) | 0 (0%) | 1 (<0.1%) |
| Chest | 0 (0%) | 1 (0.1%) | 1 (<0.1%) |
| Chinese Journal of Epidemiology | 0 (0%) | 1 (0.1%) | 1 (<0.1%) |

| **Journal** | **Systematic reviews without meta-analysis n=387*** | **Systematic reviews with meta-analysis n=785*** | **Total**, n=1172* |
|---|---|---|---|
| Chronic Obstructive Pulmonary Diseases | 0 (0%) | 1 (0.1%) | 1 (<0.1%) |
| Clinical & Experimental Rheumatology | 0 (0%) | 1 (0.1%) | 1 (<0.1%) |
| Clinical and Experimental Nephrology | 0 (0%) | 1 (0.1%) | 1 (<0.1%) |
| Clinical and Experimental Ophthalmology | 0 (0%) | 1 (0.1%) | 1 (<0.1%) |
| Clinical Endocrinology | 1 (0.3%) | 0 (0%) | 1 (<0.1%) |
| Clinical Journal of The American Society of Nephrology | 0 (0%) | 1 (0.1%) | 1 (<0.1%) |
| Clinical Medicine Insights: Women's Health | 0 (0%) | 1 (0.1%) | 1 (<0.1%) |
| Clinical Neurology and Neurosurgery | 1 (0.3%) | 0 (0%) | 1 (<0.1%) |
| Clinical Obesity | 0 (0%) | 1 (0.1%) | 1 (<0.1%) |
| Clinical Rheumatology | 1 (0.3%) | 0 (0%) | 1 (<0.1%) |
| CoDAs | 1 (0.3%) | 0 (0%) | 1 (<0.1%) |
| Critical Reviews in Food Science and Nutrition | 0 (0%) | 1 (0.1%) | 1 (<0.1%) |
| Critical Reviews in Oncology/Hematology | 0 (0%) | 1 (0.1%) | 1 (<0.1%) |
| Cureus | 1 (0.3%) | 0 (0%) | 1 (<0.1%) |
| Current Addiction Reports | 1 (0.3%) | 0 (0%) | 1 (<0.1%) |
| Current Cardiology Reviews | 0 (0%) | 1 (0.1%) | 1 (<0.1%) |
| Current Clinical Pharmacology | 0 (0%) | 1 (0.1%) | 1 (<0.1%) |
| Current Eye Research | 0 (0%) | 1 (0.1%) | 1 (<0.1%) |
| Current Hypertension Reports | 0 (0%) | 1 (0.1%) | 1 (<0.1%) |
| Current Hypertension Reviews | 1 (0.3%) | 0 (0%) | 1 (<0.1%) |
| Current Infectious Disease Reports | 0 (0%) | 1 (0.1%) | 1 (<0.1%) |
| Current Medical Research and Opinion | 1 (0.3%) | 0 (0%) | 1 (<0.1%) |
| Current Obesity Reports | 1 (0.3%) | 0 (0%) | 1 (<0.1%) |
| Current Orthopaedic Practice | 0 (0%) | 1 (0.1%) | 1 (<0.1%) |
| Current Problems in Cancer | 0 (0%) | 1 (0.1%) | 1 (<0.1%) |
| Dementia & Neuropsychologia | 1 (0.3%) | 0 (0%) | 1 (<0.1%) |
| Dental Traumatology | 0 (0%) | 1 (0.1%) | 1 (<0.1%) |
| Dermatology | 0 (0%) | 1 (0.1%) | 1 (<0.1%) |
| Dermatology Online Journal | 1 (0.3%) | 0 (0%) | 1 (<0.1%) |
| Deutsches Arzteblatt International | 0 (0%) | 1 (0.1%) | 1 (<0.1%) |
| Diabetes and Vascular Disease Research | 0 (0%) | 1 (0.1%) | 1 (<0.1%) |
| Diabetes, Metabolic Syndrome and Obesity: Targets and Therapy | 1 (0.3%) | 0 (0%) | 1 (<0.1%) |
| Diabetes/Metabolism Research and Reviews | 0 (0%) | 1 (0.1%) | 1 (<0.1%) |
| Diabetology & Metabolic Syndrome | 0 (0%) | 1 (0.1%) | 1 (<0.1%) |
| Diagnostic Microbiology and Infectious Disease | 0 (0%) | 1 (0.1%) | 1 (<0.1%) |

| **Journal** | **Systematic reviews without meta-analysis n=387*** | **Systematic reviews with meta-analysis n=785*** | **Total**, n=1172* |
|---|---|---|---|
| Diagnostics | 1 (0.3%) | 0 (0%) | 1 (<0.1%) |
| Digestive and Liver Disease | 0 (0%) | 1 (0.1%) | 1 (<0.1%) |
| Diseases of the Colon & Rectum | 1 (0.3%) | 0 (0%) | 1 (<0.1%) |
| Drug and Alcohol Review | 0 (0%) | 1 (0.1%) | 1 (<0.1%) |
| Drug Safety | 1 (0.3%) | 0 (0%) | 1 (<0.1%) |
| Dysphagia | 0 (0%) | 1 (0.1%) | 1 (<0.1%) |
| Eating and Weight Disorders | 0 (0%) | 1 (0.1%) | 1 (<0.1%) |
| Encephale | 1 (0.3%) | 0 (0%) | 1 (<0.1%) |
| Endocrine Practice | 0 (0%) | 1 (0.1%) | 1 (<0.1%) |
| Endoscopy | 0 (0%) | 1 (0.1%) | 1 (<0.1%) |
| Environmental Science and Pollution Research | 0 (0%) | 1 (0.1%) | 1 (<0.1%) |
| Epidemiology and Psychiatric Science | 0 (0%) | 1 (0.1%) | 1 (<0.1%) |
| Ethiopian Journal of Health Sciences | 0 (0%) | 1 (0.1%) | 1 (<0.1%) |
| European Eating Disorders Review | 0 (0%) | 1 (0.1%) | 1 (<0.1%) |
| European Journal of Cancer Prevention | 1 (0.3%) | 0 (0%) | 1 (<0.1%) |
| European Journal of Cardiovascular Nursing | 1 (0.3%) | 0 (0%) | 1 (<0.1%) |
| European Journal of Heart Failure | 1 (0.3%) | 0 (0%) | 1 (<0.1%) |
| European Journal of Hospital Pharmacy | 1 (0.3%) | 0 (0%) | 1 (<0.1%) |
| European Journal of Neurology | 0 (0%) | 1 (0.1%) | 1 (<0.1%) |
| European Journal of Nutrition | 1 (0.3%) | 0 (0%) | 1 (<0.1%) |
| European Journal of Pain | 0 (0%) | 1 (0.1%) | 1 (<0.1%) |
| European Journal of Preventive Cardiology | 1 (0.3%) | 0 (0%) | 1 (<0.1%) |
| European Journal of Radiology | 1 (0.3%) | 0 (0%) | 1 (<0.1%) |
| European Journal of Sport Science | 0 (0%) | 1 (0.1%) | 1 (<0.1%) |
| European Journal of Surgical Oncology | 1 (0.3%) | 0 (0%) | 1 (<0.1%) |
| European Psychiatry | 0 (0%) | 1 (0.1%) | 1 (<0.1%) |
| European Respiratory Review | 0 (0%) | 1 (0.1%) | 1 (<0.1%) |
| European Thyroid Journal | 0 (0%) | 1 (0.1%) | 1 (<0.1%) |
| Expert Review of Anti-Infective Therapy | 0 (0%) | 1 (0.1%) | 1 (<0.1%) |
| Expert Review of Neurotherapeutics | 1 (0.3%) | 0 (0%) | 1 (<0.1%) |
| F1000Research | 0 (0%) | 1 (0.1%) | 1 (<0.1%) |
| Familial Cancer | 0 (0%) | 1 (0.1%) | 1 (<0.1%) |
| Femina | 1 (0.3%) | 0 (0%) | 1 (<0.1%) |
| Fertility Research and Practice | 0 (0%) | 1 (0.1%) | 1 (<0.1%) |
| Fisioterapia em Movimento | 0 (0%) | 1 (0.1%) | 1 (<0.1%) |
| Food and Nutrition Bulletin | 1 (0.3%) | 0 (0%) | 1 (<0.1%) |
| Foot and Ankle Quarterly | 0 (0%) | 1 (0.1%) | 1 (<0.1%) |
| Frontiers in Physiology | 0 (0%) | 1 (0.1%) | 1 (<0.1%) |
| Gastrointestinal Endoscopy | 0 (0%) | 1 (0.1%) | 1 (<0.1%) |
| Gene Reports | 0 (0%) | 1 (0.1%) | 1 (<0.1%) |

| **Journal** | **Systematic reviews without meta-analysis n=387*** | **Systematic reviews with meta-analysis n=785*** | **Total**, n=1172* |
|---|---|---|---|
| Geriatrics & gerontology international | 0 (0%) | 1 (0.1%) | 1 (<0.1%) |
| Gerodontology | 1 (0.3%) | 0 (0%) | 1 (<0.1%) |
| Global Health Action | 1 (0.3%) | 0 (0%) | 1 (<0.1%) |
| Global Public Health | 0 (0%) | 1 (0.1%) | 1 (<0.1%) |
| Gut and Liver | 0 (0%) | 1 (0.1%) | 1 (<0.1%) |
| Harm Reduction Journal | 0 (0%) | 1 (0.1%) | 1 (<0.1%) |
| Head and neck | 0 (0%) | 1 (0.1%) | 1 (<0.1%) |
| Health Care for Women International | 0 (0%) | 1 (0.1%) | 1 (<0.1%) |
| Health Psychology | 0 (0%) | 1 (0.1%) | 1 (<0.1%) |
| Hearing Research | 1 (0.3%) | 0 (0%) | 1 (<0.1%) |
| Heart Lung and Circulation | 0 (0%) | 1 (0.1%) | 1 (<0.1%) |
| Helicobacter | 0 (0%) | 1 (0.1%) | 1 (<0.1%) |
| Hepatology | 0 (0%) | 1 (0.1%) | 1 (<0.1%) |
| Hepatology Communications | 0 (0%) | 1 (0.1%) | 1 (<0.1%) |
| Herz | 0 (0%) | 1 (0.1%) | 1 (<0.1%) |
| High Altitude Medicine and Biology | 0 (0%) | 1 (0.1%) | 1 (<0.1%) |
| Hipertension y Riesgo Vascular | 0 (0%) | 1 (0.1%) | 1 (<0.1%) |
| HIV/AIDS Research and Palliative Care | 0 (0%) | 1 (0.1%) | 1 (<0.1%) |
| Human Reproduction | 0 (0%) | 1 (0.1%) | 1 (<0.1%) |
| Human Resources for Health | 0 (0%) | 1 (0.1%) | 1 (<0.1%) |
| Human Vaccines and Immunotherapeutics | 0 (0%) | 1 (0.1%) | 1 (<0.1%) |
| Hypertension | 0 (0%) | 1 (0.1%) | 1 (<0.1%) |
| Indian Journal of Dental Research | 0 (0%) | 1 (0.1%) | 1 (<0.1%) |
| Indian Journal of Ophthalmology | 0 (0%) | 1 (0.1%) | 1 (<0.1%) |
| Indian Journal of Sexually Transmitted Diseases and AIDS | 1 (0.3%) | 0 (0%) | 1 (<0.1%) |
| Infectio | 1 (0.3%) | 0 (0%) | 1 (<0.1%) |
| Infectious Agents and Cancer | 0 (0%) | 1 (0.1%) | 1 (<0.1%) |
| Infectious Diseases of Poverty | 0 (0%) | 1 (0.1%) | 1 (<0.1%) |
| Infectious Diseases: Research and Treatment | 0 (0%) | 1 (0.1%) | 1 (<0.1%) |
| Influenza and Other Respiratory Viruses | 0 (0%) | 1 (0.1%) | 1 (<0.1%) |
| Injury | 1 (0.3%) | 0 (0%) | 1 (<0.1%) |
| Intellectual and Developmental Disabilities | 1 (0.3%) | 0 (0%) | 1 (<0.1%) |
| Intensive Care Medicine | 0 (0%) | 1 (0.1%) | 1 (<0.1%) |
| International Archives of Allergy and Immunology | 0 (0%) | 1 (0.1%) | 1 (<0.1%) |
| International Brazilian Journal of Urology | 0 (0%) | 1 (0.1%) | 1 (<0.1%) |
| International Cardiovascular Research Journal | 0 (0%) | 1 (0.1%) | 1 (<0.1%) |
| International Endodontic Journal | 0 (0%) | 1 (0.1%) | 1 (<0.1%) |

| **Journal** | **Systematic reviews without meta-analysis n=387*** | **Systematic reviews with meta-analysis n=785*** | **Total**, n=1172* |
|---|---|---|---|
| International Journal of Behavioral Medicine | 0 (0%) | 1 (0.1%) | 1 (<0.1%) |
| International Journal of Clinical Practice | 1 (0.3%) | 0 (0%) | 1 (<0.1%) |
| International Journal of Dermatology | 0 (0%) | 1 (0.1%) | 1 (<0.1%) |
| International Journal of General Medicine | 1 (0.3%) | 0 (0%) | 1 (<0.1%) |
| International Journal of Health Sciences | 1 (0.3%) | 0 (0%) | 1 (<0.1%) |
| International Journal of Immunopathology and Pharmacology | 0 (0%) | 1 (0.1%) | 1 (<0.1%) |
| International Journal of Injury Control and Safety Promotion | 0 (0%) | 1 (0.1%) | 1 (<0.1%) |
| International Journal of Molecular Sciences | 0 (0%) | 1 (0.1%) | 1 (<0.1%) |
| International Journal of MS Care | 0 (0%) | 1 (0.1%) | 1 (<0.1%) |
| International Journal of Nursing Studies | 1 (0.3%) | 0 (0%) | 1 (<0.1%) |
| International Journal of Offender Therapy and Comparative Criminology | 1 (0.3%) | 0 (0%) | 1 (<0.1%) |
| International Journal of Oncology | 1 (0.3%) | 0 (0%) | 1 (<0.1%) |
| International Journal of Oral & Maxillofacial Implants | 0 (0%) | 1 (0.1%) | 1 (<0.1%) |
| International Journal of Oral and Maxillofacial Surgery | 0 (0%) | 1 (0.1%) | 1 (<0.1%) |
| International Journal of Public Health | 0 (0%) | 1 (0.1%) | 1 (<0.1%) |
| International Journal of Sexual Health | 0 (0%) | 1 (0.1%) | 1 (<0.1%) |
| International Journal of Stroke | 0 (0%) | 1 (0.1%) | 1 (<0.1%) |
| International Journal Of Surgery | 0 (0%) | 1 (0.1%) | 1 (<0.1%) |
| International Journal of Tuberculosis and Lung Disease | 1 (0.3%) | 0 (0%) | 1 (<0.1%) |
| International Maritime Health | 1 (0.3%) | 0 (0%) | 1 (<0.1%) |
| International Orthopaedics | 0 (0%) | 1 (0.1%) | 1 (<0.1%) |
| International Review of Psychiatry | 0 (0%) | 1 (0.1%) | 1 (<0.1%) |
| International Urology and Nephrology | 0 (0%) | 1 (0.1%) | 1 (<0.1%) |
| Iranian Journal of Gastroenterology and Hepatology | 0 (0%) | 1 (0.1%) | 1 (<0.1%) |
| Iranian Journal of Medical Sciences | 0 (0%) | 1 (0.1%) | 1 (<0.1%) |
| Irish Journal of Medical Science | 0 (0%) | 1 (0.1%) | 1 (<0.1%) |
| Italian Journal of Pediatrics | 0 (0%) | 1 (0.1%) | 1 (<0.1%) |
| JAMA Dermatology | 0 (0%) | 1 (0.1%) | 1 (<0.1%) |
| JBI Database of Systematic Reviews & Implementation Reports | 0 (0%) | 1 (0.1%) | 1 (<0.1%) |
| JGH Open | 0 (0%) | 1 (0.1%) | 1 (<0.1%) |
| Joint, Bone, Spine | 1 (0.3%) | 0 (0%) | 1 (<0.1%) |
| Jornal Brasileiro de Patologia e Medicina Laboratoria | 1 (0.3%) | 0 (0%) | 1 (<0.1%) |
| Journal de Mycologie Medicale | 0 (0%) | 1 (0.1%) | 1 (<0.1%) |

| **Journal** | **Systematic reviews without meta-analysis n=387*** | **Systematic reviews with meta-analysis n=785*** | **Total**, n=1172* |
|---|---|---|---|
| Journal of Addiction Medicine | 0 (0%) | 1 (0.1%) | 1 (<0.1%) |
| Journal of Advanced Nursing | 1 (0.3%) | 0 (0%) | 1 (<0.1%) |
| Journal of Alzheimer's Disease | 0 (0%) | 1 (0.1%) | 1 (<0.1%) |
| Journal of Antimicrobial Chemotherapy | 0 (0%) | 1 (0.1%) | 1 (<0.1%) |
| Journal of Applied Research in Intellectual Disabilities | 0 (0%) | 1 (0.1%) | 1 (<0.1%) |
| Journal of Atrial Fibrillation | 0 (0%) | 1 (0.1%) | 1 (<0.1%) |
| Journal of Cardiac Failure | 0 (0%) | 1 (0.1%) | 1 (<0.1%) |
| Journal of cardiovascular medicine | 0 (0%) | 1 (0.1%) | 1 (<0.1%) |
| Journal of Cardiovascular Nursing | 0 (0%) | 1 (0.1%) | 1 (<0.1%) |
| Journal of Clinical Medicine | 0 (0%) | 1 (0.1%) | 1 (<0.1%) |
| Journal of Clinical Nursing | 0 (0%) | 1 (0.1%) | 1 (<0.1%) |
| Journal of Clinical Sleep Medicine | 0 (0%) | 1 (0.1%) | 1 (<0.1%) |
| Journal of Cranio-Maxillofacial Surgery | 0 (0%) | 1 (0.1%) | 1 (<0.1%) |
| Journal of Crohn's and Colitis | 0 (0%) | 1 (0.1%) | 1 (<0.1%) |
| Journal of Current Ophthalmology | 0 (0%) | 1 (0.1%) | 1 (<0.1%) |
| Journal of Diabetes and Metabolic Disorders | 0 (0%) | 1 (0.1%) | 1 (<0.1%) |
| Journal of Diabetes Investigation | 0 (0%) | 1 (0.1%) | 1 (<0.1%) |
| Journal of Diabetes Research | 0 (0%) | 1 (0.1%) | 1 (<0.1%) |
| Journal of Diabetic Nursing | 0 (0%) | 1 (0.1%) | 1 (<0.1%) |
| Journal of Dual Diagnosis | 1 (0.3%) | 0 (0%) | 1 (<0.1%) |
| Journal of Endodontics | 0 (0%) | 1 (0.1%) | 1 (<0.1%) |
| Journal of Epidemiology | 0 (0%) | 1 (0.1%) | 1 (<0.1%) |
| Journal of Evidence-Based Medicine | 0 (0%) | 1 (0.1%) | 1 (<0.1%) |
| Journal of Family Medicine and Primary Care | 0 (0%) | 1 (0.1%) | 1 (<0.1%) |
| Journal of Foot and Ankle Research | 0 (0%) | 1 (0.1%) | 1 (<0.1%) |
| Journal of Foot and Ankle Surgery | 0 (0%) | 1 (0.1%) | 1 (<0.1%) |
| Journal of Gastroenterology | 0 (0%) | 1 (0.1%) | 1 (<0.1%) |
| Journal of Gastroenterology and Hepatology | 0 (0%) | 1 (0.1%) | 1 (<0.1%) |
| Journal of Glaucoma | 0 (0%) | 1 (0.1%) | 1 (<0.1%) |
| Journal of global antimicrobial resistance | 0 (0%) | 1 (0.1%) | 1 (<0.1%) |
| Journal of Human Hypertension | 1 (0.3%) | 0 (0%) | 1 (<0.1%) |
| Journal of Infection | 0 (0%) | 1 (0.1%) | 1 (<0.1%) |
| Journal of Inherited Metabolic Disease | 1 (0.3%) | 0 (0%) | 1 (<0.1%) |
| Journal of Injury and Violence Research | 0 (0%) | 1 (0.1%) | 1 (<0.1%) |
| Journal of Internal Medicine | 0 (0%) | 1 (0.1%) | 1 (<0.1%) |
| Journal of Investigative Dermatology | 1 (0.3%) | 0 (0%) | 1 (<0.1%) |
| Journal of Laryngology & Otology | 0 (0%) | 1 (0.1%) | 1 (<0.1%) |
| Journal of Manipulative and Physiological Therapeutics | 1 (0.3%) | 0 (0%) | 1 (<0.1%) |
| Journal of Medical Microbiology | 0 (0%) | 1 (0.1%) | 1 (<0.1%) |

| **Journal** | **Systematic reviews without meta-analysis n=387*** | **Systematic reviews with meta-analysis n=785*** | **Total**, n=1172* |
|---|---|---|---|
| Journal of Mental Health | 0 (0%) | 1 (0.1%) | 1 (<0.1%) |
| Journal of NeuroInterventional Surgery | 0 (0%) | 1 (0.1%) | 1 (<0.1%) |
| Journal of Neurology | 0 (0%) | 1 (0.1%) | 1 (<0.1%) |
| Journal of Neurotrauma | 0 (0%) | 1 (0.1%) | 1 (<0.1%) |
| Journal of Obesity | 0 (0%) | 1 (0.1%) | 1 (<0.1%) |
| Journal of Occupational Health | 1 (0.3%) | 0 (0%) | 1 (<0.1%) |
| Journal of Occupational Medicine and Toxicology | 0 (0%) | 1 (0.1%) | 1 (<0.1%) |
| Journal of Ophthalmology | 0 (0%) | 1 (0.1%) | 1 (<0.1%) |
| Journal of Oral Rehabilitation | 0 (0%) | 1 (0.1%) | 1 (<0.1%) |
| Journal of Orthopaedics | 0 (0%) | 1 (0.1%) | 1 (<0.1%) |
| Journal of Parenteral and Enteral Nutrition | 1 (0.3%) | 0 (0%) | 1 (<0.1%) |
| Journal of Patient Safety | 0 (0%) | 1 (0.1%) | 1 (<0.1%) |
| Journal of Plastic Surgery and Hand Surgery | 1 (0.3%) | 0 (0%) | 1 (<0.1%) |
| Journal of Postgraduate Medicine | 0 (0%) | 1 (0.1%) | 1 (<0.1%) |
| Journal of Public Health Policy | 1 (0.3%) | 0 (0%) | 1 (<0.1%) |
| Journal of Racial and Ethnic Health Disparities | 1 (0.3%) | 0 (0%) | 1 (<0.1%) |
| Journal of Research in Medical Sciences | 0 (0%) | 1 (0.1%) | 1 (<0.1%) |
| Journal of Science and Medicine in Sport | 1 (0.3%) | 0 (0%) | 1 (<0.1%) |
| Journal of Sexual Medicine | 0 (0%) | 1 (0.1%) | 1 (<0.1%) |
| Journal of Shoulder and Elbow Surgery | 1 (0.3%) | 0 (0%) | 1 (<0.1%) |
| Journal of the American College of Nutrition | 0 (0%) | 1 (0.1%) | 1 (<0.1%) |
| Journal of the American Dental Association | 1 (0.3%) | 0 (0%) | 1 (<0.1%) |
| Journal of the Medical Association of Thailand | 1 (0.3%) | 0 (0%) | 1 (<0.1%) |
| Journal of the Pancreas | 0 (0%) | 1 (0.1%) | 1 (<0.1%) |
| Journal of Translational Medicine | 0 (0%) | 1 (0.1%) | 1 (<0.1%) |
| Journal of Viral Hepatitis | 0 (0%) | 1 (0.1%) | 1 (<0.1%) |
| Journal of Voice | 0 (0%) | 1 (0.1%) | 1 (<0.1%) |
| Journal of Wound Care | 0 (0%) | 1 (0.1%) | 1 (<0.1%) |
| Journal of Zanjan University of Medical Sciences and Health Services | 0 (0%) | 1 (0.1%) | 1 (<0.1%) |
| Kathmandu University Medical Journal | 1 (0.3%) | 0 (0%) | 1 (<0.1%) |
| Kidney International | 0 (0%) | 1 (0.1%) | 1 (<0.1%) |
| Knee Surgery, Sports Traumatology, Arthroscopy | 0 (0%) | 1 (0.1%) | 1 (<0.1%) |
| Laryngoscope | 0 (0%) | 1 (0.1%) | 1 (<0.1%) |
| Lupus | 0 (0%) | 1 (0.1%) | 1 (<0.1%) |
| Malawi Medical Journal | 1 (0.3%) | 0 (0%) | 1 (<0.1%) |

| Journal | Systematic reviews without meta-analysis n=387* | Systematic reviews with meta-analysis n=785* | Total, n=1172* |
|---|---|---|---|
| Mastology | 1 (0.3%) | 0 (0%) | 1 (<0.1%) |
| Maternal and Child Health Journal | 1 (0.3%) | 0 (0%) | 1 (<0.1%) |
| Medical Education | 1 (0.3%) | 0 (0%) | 1 (<0.1%) |
| Medicina Clínica | 1 (0.3%) | 0 (0%) | 1 (<0.1%) |
| Microbial Drug Resistance | 0 (0%) | 1 (0.1%) | 1 (<0.1%) |
| Middle East Fertility Society Journal | 0 (0%) | 1 (0.1%) | 1 (<0.1%) |
| Military Medicine | 0 (0%) | 1 (0.1%) | 1 (<0.1%) |
| Minerva Urologica e Nefrologica | 0 (0%) | 1 (0.1%) | 1 (<0.1%) |
| Molecular Genetics and Metabolism | 1 (0.3%) | 0 (0%) | 1 (<0.1%) |
| Movement Disorders | 0 (0%) | 1 (0.1%) | 1 (<0.1%) |
| Mycoses | 0 (0%) | 1 (0.1%) | 1 (<0.1%) |
| National Medical Journal of India | 0 (0%) | 1 (0.1%) | 1 (<0.1%) |
| Nephrology | 0 (0%) | 1 (0.1%) | 1 (<0.1%) |
| Neuro-Oncology Practice | 1 (0.3%) | 0 (0%) | 1 (<0.1%) |
| Neuroepidemiology | 0 (0%) | 1 (0.1%) | 1 (<0.1%) |
| NeuroImage Clinical | 0 (0%) | 1 (0.1%) | 1 (<0.1%) |
| Neurological Sciences | 0 (0%) | 1 (0.1%) | 1 (<0.1%) |
| Neurology | 0 (0%) | 1 (0.1%) | 1 (<0.1%) |
| Neuropathology and Applied Neurobiology | 0 (0%) | 1 (0.1%) | 1 (<0.1%) |
| Neuropsychiatric Disease and Treatment | 1 (0.3%) | 0 (0%) | 1 (<0.1%) |
| Neuroscience & Biobehavioral Reviews | 0 (0%) | 1 (0.1%) | 1 (<0.1%) |
| Nicotine & Tobacco Research | 0 (0%) | 1 (0.1%) | 1 (<0.1%) |
| Nure Investigación | 1 (0.3%) | 0 (0%) | 1 (<0.1%) |
| Nutrition Metabolism and Cardiovascular Diseases | 0 (0%) | 1 (0.1%) | 1 (<0.1%) |
| Nutrition Reviews | 1 (0.3%) | 0 (0%) | 1 (<0.1%) |
| Obesity | 0 (0%) | 1 (0.1%) | 1 (<0.1%) |
| Occupational and Environmental Medicine | 0 (0%) | 1 (0.1%) | 1 (<0.1%) |
| Occupational Medicine | 1 (0.3%) | 0 (0%) | 1 (<0.1%) |
| Oncology Nursing Forum | 1 (0.3%) | 0 (0%) | 1 (<0.1%) |
| Open Access Macedonian Journal of Medical Sciences | 1 (0.3%) | 0 (0%) | 1 (<0.1%) |
| Ophthalmology Retina | 1 (0.3%) | 0 (0%) | 1 (<0.1%) |
| Optometry & Visual Performance | 1 (0.3%) | 0 (0%) | 1 (<0.1%) |
| Optometry and Vision Science | 0 (0%) | 1 (0.1%) | 1 (<0.1%) |
| Orphanet Journal of Rare Diseases | 0 (0%) | 1 (0.1%) | 1 (<0.1%) |
| Orthopaedic Journal of Sports Medicine | 0 (0%) | 1 (0.1%) | 1 (<0.1%) |
| Otolaryngology-Head and Neck Surgery | 0 (0%) | 1 (0.1%) | 1 (<0.1%) |
| Otology & Neurotology | 1 (0.3%) | 0 (0%) | 1 (<0.1%) |
| Pain Management Nursing | 1 (0.3%) | 0 (0%) | 1 (<0.1%) |
| Pain Medicine | 1 (0.3%) | 0 (0%) | 1 (<0.1%) |
| Pain Research and Management | 0 (0%) | 1 (0.1%) | 1 (<0.1%) |

| **Journal** | **Systematic reviews without meta-analysis n=387*** | **Systematic reviews with meta-analysis n=785*** | **Total**, n=1172* |
|---|---|---|---|
| Patient Preference and Adherence | 1 (0.3%) | 0 (0%) | 1 (<0.1%) |
| Pediatric Emergency Care | 0 (0%) | 1 (0.1%) | 1 (<0.1%) |
| Pesquisa Brasileira em Odontopediatria e Clínica Integrada | 0 (0%) | 1 (0.1%) | 1 (<0.1%) |
| Physical Therapy Research | 0 (0%) | 1 (0.1%) | 1 (<0.1%) |
| Physician and Sportsmedicine | 1 (0.3%) | 0 (0%) | 1 (<0.1%) |
| Plastic and Reconstructive Surgery | 0 (0%) | 1 (0.1%) | 1 (<0.1%) |
| Platelets | 1 (0.3%) | 0 (0%) | 1 (<0.1%) |
| PM and R | 1 (0.3%) | 0 (0%) | 1 (<0.1%) |
| Population Health Metrics | 0 (0%) | 1 (0.1%) | 1 (<0.1%) |
| Postgraduate Medicine | 0 (0%) | 1 (0.1%) | 1 (<0.1%) |
| Primary Care Respiratory Journal | 1 (0.3%) | 0 (0%) | 1 (<0.1%) |
| Progress in Neuro Psychopharmacology and Biological Psychiatry | 1 (0.3%) | 0 (0%) | 1 (<0.1%) |
| Psychiatria Danubina | 1 (0.3%) | 0 (0%) | 1 (<0.1%) |
| Psycho-Oncology | 0 (0%) | 1 (0.1%) | 1 (<0.1%) |
| Psychology, Health & Medicine | 0 (0%) | 1 (0.1%) | 1 (<0.1%) |
| Psychosomatic Medicine | 0 (0%) | 1 (0.1%) | 1 (<0.1%) |
| Psychosomatics | 1 (0.3%) | 0 (0%) | 1 (<0.1%) |
| Regional Anesthesia and Pain Medicine | 0 (0%) | 1 (0.1%) | 1 (<0.1%) |
| Research in Developmental Disabilities | 1 (0.3%) | 0 (0%) | 1 (<0.1%) |
| Respiratory Care | 0 (0%) | 1 (0.1%) | 1 (<0.1%) |
| Respiratory Medicine | 0 (0%) | 1 (0.1%) | 1 (<0.1%) |
| Rev. Baiana Saúde Pública | 1 (0.3%) | 0 (0%) | 1 (<0.1%) |
| Reviews in Clinical Gerontology | 1 (0.3%) | 0 (0%) | 1 (<0.1%) |
| Reviews in Endocrine and Metabolic Disorders | 0 (0%) | 1 (0.1%) | 1 (<0.1%) |
| Reviews in Medical Microbiology | 0 (0%) | 1 (0.1%) | 1 (<0.1%) |
| Revista Brasileira de Epidemiologia | 1 (0.3%) | 0 (0%) | 1 (<0.1%) |
| Revista Brasileira De Reumatologia | 0 (0%) | 1 (0.1%) | 1 (<0.1%) |
| Revista Colombiana de Obstetricia y Ginecología | 1 (0.3%) | 0 (0%) | 1 (<0.1%) |
| Revista Colombiana de Psiquiatría | 1 (0.3%) | 0 (0%) | 1 (<0.1%) |
| Revista da Sociedade Brasileira de Clínica Medica | 1 (0.3%) | 0 (0%) | 1 (<0.1%) |
| Revista de APS | 1 (0.3%) | 0 (0%) | 1 (<0.1%) |
| Revista de la Asociación Española de Especialistas en Medicina del Trabajo | 1 (0.3%) | 0 (0%) | 1 (<0.1%) |
| Revista de la Facultad de Medicina | 1 (0.3%) | 0 (0%) | 1 (<0.1%) |
| Revista de Neurología | 1 (0.3%) | 0 (0%) | 1 (<0.1%) |
| Revista Eletrônica Saúde Mental Álcool e Drogas | 1 (0.3%) | 0 (0%) | 1 (<0.1%) |
| Revista Española de Drogodependencias | 1 (0.3%) | 0 (0%) | 1 (<0.1%) |

| Journal | Systematic reviews without meta-analysis n=387* | Systematic reviews with meta-analysis n=785* | Total, n=1172* |
|---|---|---|---|
| Revista Española de Geriatría y Gerontología | 0 (0%) | 1 (0.1%) | 1 (<0.1%) |
| Revista Española de Salud Pública | 1 (0.3%) | 0 (0%) | 1 (<0.1%) |
| Revista Latino-Americana de Enfermagem | 0 (0%) | 1 (0.1%) | 1 (<0.1%) |
| Revista Latinoamericana de Psicologia | 1 (0.3%) | 0 (0%) | 1 (<0.1%) |
| Revista Mexicana de Trastornos Alimentarios | 1 (0.3%) | 0 (0%) | 1 (<0.1%) |
| Revista Pesquisa em Fisioterapia | 1 (0.3%) | 0 (0%) | 1 (<0.1%) |
| Revista Uruguaya de Enfermería | 1 (0.3%) | 0 (0%) | 1 (<0.1%) |
| Salud Pública de México | 1 (0.3%) | 0 (0%) | 1 (<0.1%) |
| Saudi Dental Journal | 0 (0%) | 1 (0.1%) | 1 (<0.1%) |
| Scandinavian Journal of Caring Sciences | 1 (0.3%) | 0 (0%) | 1 (<0.1%) |
| Scandinavian Journal of Primary Health Care | 0 (0%) | 1 (0.1%) | 1 (<0.1%) |
| Scandinavian Journal of Public Health | 1 (0.3%) | 0 (0%) | 1 (<0.1%) |
| Schizophrenia Bulletin | 0 (0%) | 1 (0.1%) | 1 (<0.1%) |
| Schizophrenia Research | 1 (0.3%) | 0 (0%) | 1 (<0.1%) |
| Scientific World Journal | 1 (0.3%) | 0 (0%) | 1 (<0.1%) |
| Social Psychiatry and Psychiatric Epidemiology | 1 (0.3%) | 0 (0%) | 1 (<0.1%) |
| Stroke | 0 (0%) | 1 (0.1%) | 1 (<0.1%) |
| Supportive Care in Cancer | 0 (0%) | 1 (0.1%) | 1 (<0.1%) |
| Systematic Reviews | 0 (0%) | 1 (0.1%) | 1 (<0.1%) |
| The American Journal of Hospice and Palliative Care | 1 (0.3%) | 0 (0%) | 1 (<0.1%) |
| The Brazilian journal of infectious diseases | 1 (0.3%) | 0 (0%) | 1 (<0.1%) |
| The Journal of Allergy and Clinical Immunology in Practice | 0 (0%) | 1 (0.1%) | 1 (<0.1%) |
| The Journal of Frailty & Aging | 1 (0.3%) | 0 (0%) | 1 (<0.1%) |
| The Journal of Gastroenterology and Hepatology | 1 (0.3%) | 0 (0%) | 1 (<0.1%) |
| The Journal of Rheumatology | 0 (0%) | 1 (0.1%) | 1 (<0.1%) |
| The Journal of Tehran Heart Center | 0 (0%) | 1 (0.1%) | 1 (<0.1%) |
| The Lancet Diabetes & Endocrinology | 0 (0%) | 1 (0.1%) | 1 (<0.1%) |
| The Lancet Neurology | 0 (0%) | 1 (0.1%) | 1 (<0.1%) |
| The Lancet Oncology | 0 (0%) | 1 (0.1%) | 1 (<0.1%) |
| The Brazilian Journal of Otorhinolaryngology | 1 (0.3%) | 0 (0%) | 1 (<0.1%) |
| Therapeutic Apheresis and Dialysis | 0 (0%) | 1 (0.1%) | 1 (<0.1%) |
| Thesis | 0 (0%) | 1 (0.1%) | 1 (<0.1%) |
| Thrombosis and Haemostasis | 0 (0%) | 1 (0.1%) | 1 (<0.1%) |
| Tijdschrift voor Psychiatrie | 1 (0.3%) | 0 (0%) | 1 (<0.1%) |

| **Journal** | **Systematic reviews without meta-analysis n=387*** | **Systematic reviews with meta-analysis n=785*** | **Total**, n=1172* |
|---|---|---|---|
| Traffic Injury Prevention | 0 (0%) | 1 (0.1%) | 1 (<0.1%) |
| Transgender Health | 0 (0%) | 1 (0.1%) | 1 (<0.1%) |
| Translational Behavioral Medicine | 1 (0.3%) | 0 (0%) | 1 (<0.1%) |
| Turk Psikiyatri Dergisi | 0 (0%) | 1 (0.1%) | 1 (<0.1%) |
| Turkish Journal of Gastroenterology | 1 (0.3%) | 0 (0%) | 1 (<0.1%) |
| Urologia Internationalis | 0 (0%) | 1 (0.1%) | 1 (<0.1%) |
| Urology Journal | 0 (0%) | 1 (0.1%) | 1 (<0.1%) |
| Vascular Health and Risk Management | 0 (0%) | 1 (0.1%) | 1 (<0.1%) |
| Vector Borne and Zoonotic Diseases | 0 (0%) | 1 (0.1%) | 1 (<0.1%) |
| Virology Journal | 0 (0%) | 1 (0.1%) | 1 (<0.1%) |
| Viruses | 0 (0%) | 1 (0.1%) | 1 (<0.1%) |
| Wiener Klinische Wochenschrift | 1 (0.3%) | 0 (0%) | 1 (<0.1%) |
| Women and Birth | 1 (0.3%) | 0 (0%) | 1 (<0.1%) |

*Abbreviations:* SR, systematic review
* n (%)

**Figure S1.** Geographic heat map of the first author reported affiliation in systematic reviews of prevalence in adults 2010-2020

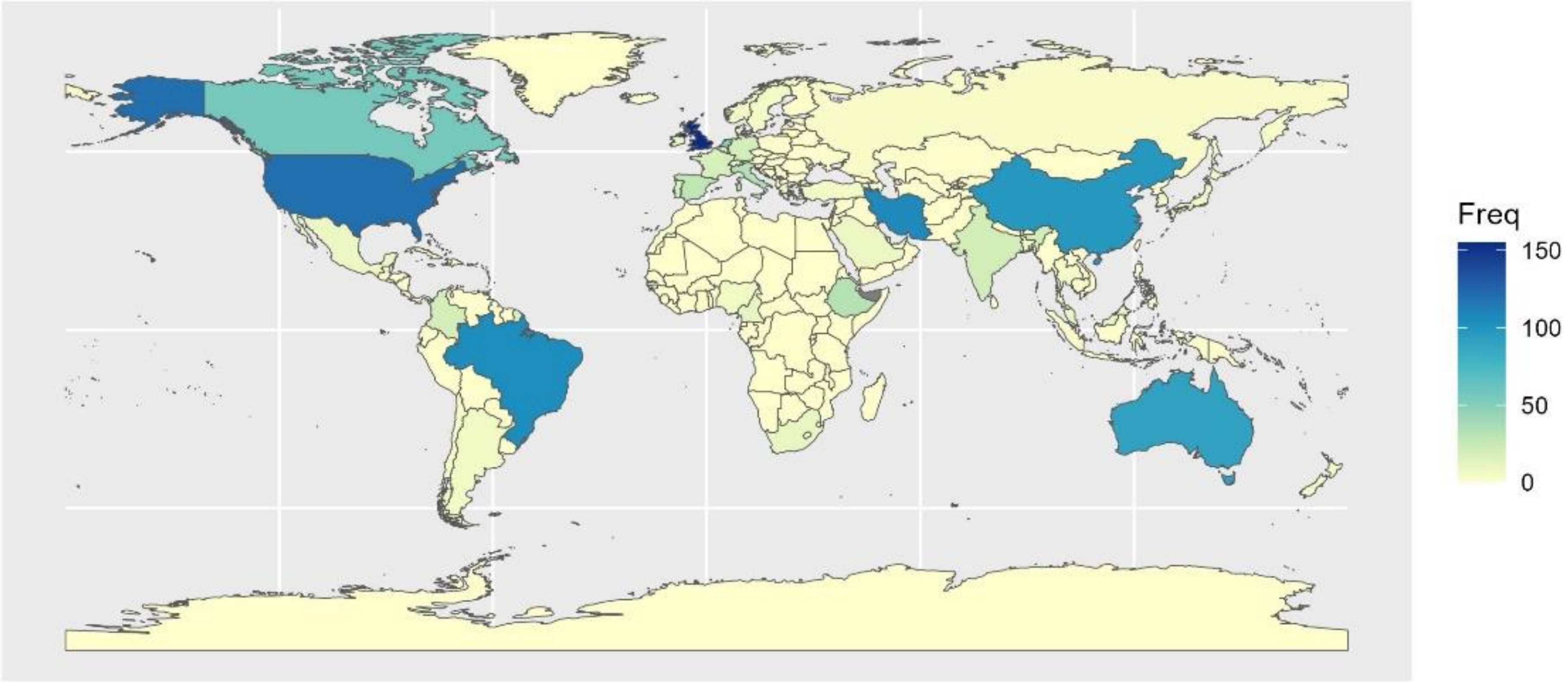

**Table S2.** Country of affiliation reported by first author of systematic reviews of prevalence in adult populations published 2010-2020, by total frequency

| **Country** | **SR without meta-analysis** n=387* | **SR with meta-analysis** n=785* | **Total** n=1172* |
|---|---|---|---|
| United Kingdom | 55 (14.2%) | 100 (12.7%) | 155 (13.2%) |
| United States | 38 (9.8%) | 82 (10.4%) | 120 (10.2%) |
| Iran | 30 (7.8%) | 77 (9.8%) | 107 (9.1%) |
| Brazil | 47 (12%) | 58 (7.4%) | 105 (9.0%) |
| China | 26 (6.7%) | 72 (9.2%) | 98 (8.4%) |
| Australia | 37 (9.6%) | 54 (6.9%) | 91 (7.8%) |
| Canada | 11 (2.8%) | 45 (5.7%) | 56 (4.8%) |
| Netherlands | 16 (4.1%) | 34 (4.3%) | 50 (4.3%) |
| Ethiopia | 4 (1.0%) | 28 (3.6%) | 32 (2.7%) |
| Italy | 9 (2.3%) | 23 (2.9%) | 32 (2.7%) |
| Spain | 11 (2.8%) | 17 (2.2%) | 28 (2.4%) |
| India | 7 (1.8%) | 13 (1.7%) | 20 (1.7%) |
| Colombia | 7 (1.8%) | 12 (1.5%) | 19 (1.6%) |
| Germany | 13 (3.4%) | 6 (0.8%) | 19 (1.6%) |
| France | 6 (1.6%) | 12 (1.5%) | 18 (1.5%) |
| Portugal | 10 (2.6%) | 7 (0.9%) | 17 (1.5%) |
| Singapore | 4 (1.0%) | 10 (1.3%) | 14 (1.2%) |
| Switzerland | 1 (0.3%) | 13 (1.7%) | 14 (1.2%) |
| Belgium | 2 (0.5%) | 8 (1.0%) | 10 (0.9%) |
| Cameroon | 2 (0.5%) | 8 (1.0%) | 10 (0.9%) |
| Ireland | 2 (0.5%) | 8 (1.0%) | 10 (0.9%) |
| Malaysia | 5 (1.3%) | 5 (0.6%) | 10 (0.9%) |
| South Africa | 4 (1.0%) | 6 (0.8%) | 10 (0.9%) |
| Denmark | 1 (0.3%) | 7 (0.9%) | 8 (0.7%) |
| Greece | 1 (0.3%) | 7 (0.9%) | 8 (0.7%) |
| Mexico | 3 (0.8%) | 5 (0.6%) | 8 (0.7%) |
| Saudi Arabia | 5 (1.3%) | 3 (0.4%) | 8 (0.7%) |
| South Korea | 1 (0.3%) | 6 (0.8%) | 7 (0.6%) |
| Argentina | 2 (0.5%) | 4 (0.5%) | 6 (0.5%) |
| Japan | 1 (0.3%) | 5 (0.6%) | 6 (0.5%) |

| Country | SR without meta-analysis n=387* | SR with meta-analysis n=785* | Total n=1172* |
|---|---|---|---|
| Nigeria | 2 (0.5%) | 4 (0.5%) | 6 (0.5%) |
| New Zealand | 0 (0%) | 5 (0.6%) | 5 (0.4%) |
| Norway | 2 (0.5%) | 3 (0.4%) | 5 (0.4%) |
| Sweden | 2 (0.5%) | 3 (0.4%) | 5 (0.4%) |
| Turkey | 2 (0.5%) | 3 (0.4%) | 5 (0.4%) |
| Qatar | 3 (0.8%) | 1 (0.1%) | 4 (0.3%) |
| Thailand | 1 (0.3%) | 3 (0.4%) | 4 (0.3%) |
| Austria | 1 (0.3%) | 2 (0.3%) | 3 (0.3%) |
| Bangladesh | 1 (0.3%) | 2 (0.3%) | 3 (0.3%) |
| Chile | 1 (0.3%) | 2 (0.3%) | 3 (0.3%) |
| Ghana | 0 (0%) | 3 (0.4%) | 3 (0.3%) |
| Russia | 2 (0.5%) | 1 (0.1%) | 3 (0.3%) |
| Israel | 0 (0%) | 2 (0.3%) | 2 (0.2%) |
| Kyrgyzstan | 0 (0%) | 2 (0.3%) | 2 (0.2%) |
| Lebanon | 0 (0%) | 2 (0.3%) | 2 (0.2%) |
| Taiwan | 0 (0%) | 2 (0.3%) | 2 (0.2%) |
| Benin | 0 (0%) | 1 (0.1%) | 1 (<0.1%) |
| Burkina Faso | 1 (0.3%) | 0 (0%) | 1 (<0.1%) |
| Democratic Republic of the Congo | 1 (0.3%) | 0 (0%) | 1 (<0.1%) |
| Costa Rica | 0 (0%) | 1 (0.1%) | 1 (<0.1%) |
| Croatia | 1 (0.3%) | 0 (0%) | 1 (<0.1%) |
| Czech Republic | 0 (0%) | 1 (0.1%) | 1 (<0.1%) |
| Egypt | 1 (0.3%) | 0 (0%) | 1 (<0.1%) |
| Guatemala | 1 (0.3%) | 0 (0%) | 1 (<0.1%) |
| Hong Kong | 0 (0%) | 1 (0.1%) | 1 (<0.1%) |
| Hungary | 1 (0.3%) | 0 (0%) | 1 (<0.1%) |
| Indonesia | 1 (0.3%) | 0 (0%) | 1 (<0.1%) |
| Kazakhstan | 0 (0%) | 1 (0.1%) | 1 (<0.1%) |
| Malawi | 0 (0%) | 1 (0.1%) | 1 (<0.1%) |
| Morocco | 0 (0%) | 1 (0.1%) | 1 (<0.1%) |
| Pakistan | 0 (0%) | 1 (0.1%) | 1 (<0.1%) |

| Country | SR without meta-analysis n=387* | SR with meta-analysis n=785* | Total n=1172* |
|---|---|---|---|
| Peru | 0 (0%) | 1 (0.1%) | 1 (<0.1%) |
| Serbia | 0 (0%) | 1 (0.1%) | 1 (<0.1%) |
| Sri Lanka | 1 (0.3%) | 0 (0%) | 1 (<0.1%) |
| United Arab Emirates | 1 (0.3%) | 0 (0%) | 1 (<0.1%) |

*Abbreviations:* SR, systematic review
* n (%)

**Figure S2.** Medical specialty or subject of systematic reviews of prevalence in adult populations, published 2010-2020

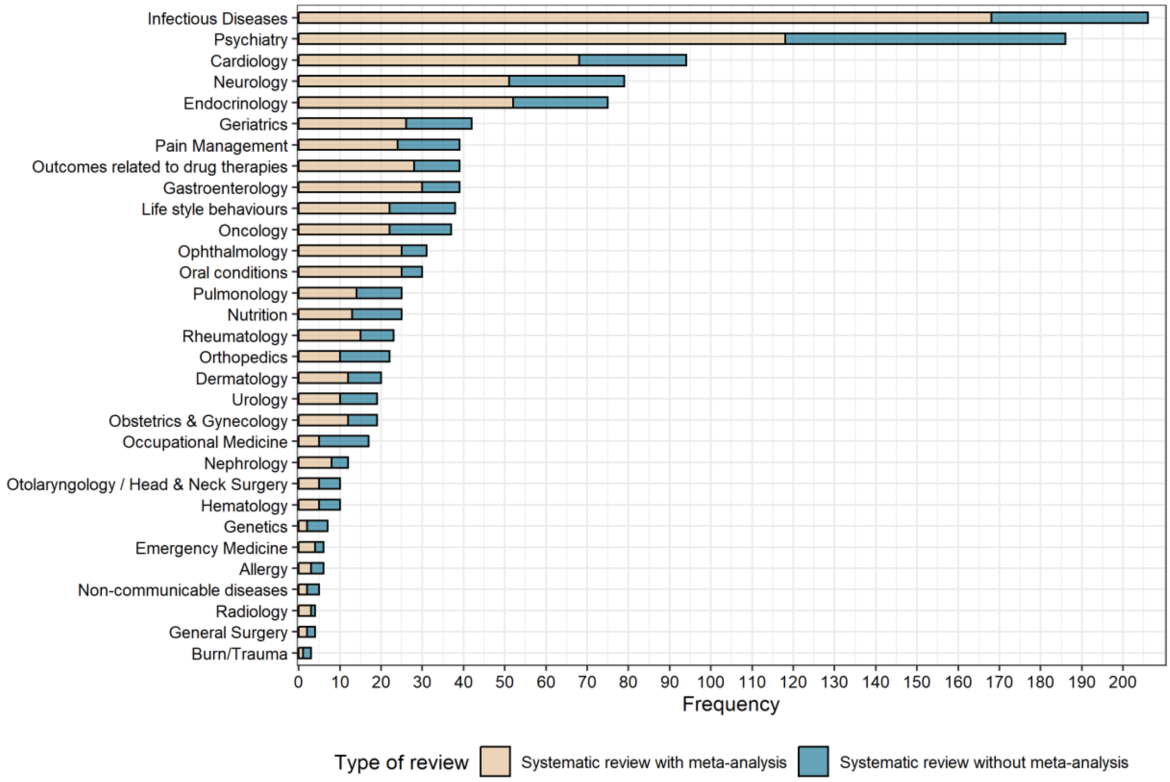

**Table S3.** Medical specialty or subject of systematic reviews of prevalence in adult populations, published 2010-2020

| **Medical specialty** | **SR without meta-analysis** n=387* | **SR with meta-analysis** n=785* | **Total** n=1172* |
|---|---|---|---|
| Infectious diseases | 38 (9.8%) | 168 (21.4%) | 206 (17.6%) |
| Psychiatry | 68 (18%) | 118 (15.0%) | 186 (15.9%) |
| Cardiology | 26 (6.7%) | 68 (8.7%) | 94 (8.0%) |
| Neurology | 28 (7.2%) | 51 (6.5%) | 79 (6.7%) |
| Endocrinology | 23 (5.9%) | 52 (6.6%) | 75 (6.4%) |
| Geriatrics | 16 (4.1%) | 26 (3.3%) | 42 (3.6%) |
| Gastroenterology | 9 (2.3%) | 30 (3.8%) | 39 (3.3%) |
| Outcomes related to drug therapies | 11 (2.8%) | 28 (3.6%) | 39 (3.3%) |
| Pain management | 15 (3.9%) | 24 (3.1%) | 39 (3.3%) |
| Lifestyle behaviours | 16 (4.1%) | 22 (2.8%) | 38 (3.2%) |
| Oncology | 15 (3.9%) | 22 (2.8%) | 37 (3.2%) |
| Ophthalmology | 6 (1.6%) | 25 (3.2%) | 31 (2.6%) |
| Oral conditions | 5 (1.3%) | 25 (3.2%) | 30 (2.6%) |
| Nutrition | 12 (3.1%) | 13 (1.7%) | 25 (2.1%) |
| Pulmonology | 11 (2.8%) | 14 (1.8%) | 25 (2.1%) |
| Rheumatology | 8 (2.1%) | 15 (1.9%) | 23 (2.0%) |
| Orthopaedics | 12 (3.1%) | 10 (1.3%) | 22 (1.9%) |
| Dermatology | 8 (2.1%) | 12 (1.5%) | 20 (1.7%) |
| Obstetrics & gynaecology | 7 (1.8%) | 12 (1.5%) | 19 (1.6%) |
| Urology | 9 (2.3%) | 10 (1.3%) | 19 (1.6%) |
| Occupational Medicine | 12 (3.1%) | 5 (0.6%) | 17 (1.5%) |
| Nephrology | 4 (1.0%) | 8 (1.0%) | 12 (1.0%) |
| Haematology | 5 (1.3%) | 5 (0.6%) | 10 (0.9%) |
| Otolaryngology / head & neck surgery | 5 (1.3%) | 5 (0.6%) | 10 (0.9%) |
| Genetics | 5 (1.3%) | 2 (0.3%) | 7 (0.6%) |
| Allergy | 3 (0.8%) | 3 (0.4%) | 6 (0.5%) |
| Emergency medicine | 2 (0.5%) | 4 (0.5%) | 6 (0.5%) |
| Non-communicable diseases | 3 (0.8%) | 2 (0.3%) | 5 (0.4%) |
| General surgery | 2 (0.5%) | 2 (0.3%) | 4 (0.3%) |
| Radiology | 1 (0.3%) | 3 (0.4%) | 4 (0.3%) |
| Burns/trauma | 2 (0.5%) | 1 (0.1%) | 3 (0.3%) |

*Abbreviations:* IQR, interquartile range; SR, systematic review
* n (%)

**Table S4.** Populations included in systematic reviews of prevalence in adult populations 2010-2020

| **Population*** | **SR without meta-analysis** n=387† | **SR with meta-analysis** n=785† | **Total** n=1172† |
|---|---|---|---|
| Adults in general‡ | 146 (37.7%) | 290 (36.9%) | 436 (37.2%) |
| Adults with a specific condition or characteristic | 101 (26.1%) | 281 (35.8%) | 382 (32.6%) |
| Older population | 47 (12.1%) | 53 (6.8%) | 100 (8.5%) |
| Women and pregnant women | 29 (7.5%) | 60 (7.6%) | 89 (7.6%) |
| Workers | 16 (4.1%) | 18 (2.3%) | 34 (2.9%) |
| Indigenous populations/ethnic minorities | 6 (1.6%) | 14 (1.8%) | 20 (1.7%) |
| Students | 7 (1.8%) | 11 (1.4%) | 18 (1.5%) |
| Hospitalised patients | 3 (0.8%) | 14 (1.8%) | 17 (1.5%) |
| Prison populations | 8 (2.1%) | 8 (1.0%) | 16 (1.4%) |
| Men | 6 (1.6%) | 9 (1.1%) | 15 (1.3%) |
| Dancers/actors/singers | 8 (2.1%) | 6 (0.8%) | 14 (1.2%) |
| Post-conflict adults (including veterans) | 4 (1.0%) | 4 (0.5%) | 8 (0.7%) |
| Sex workers | 0 (0%) | 8 (1.0%) | 8 (0.7%) |
| Migrants | 2 (0.5%) | 4 (0.5%) | 6 (0.5%) |
| Homeless people | 1 (0.3%) | 3 (0.4%) | 4 (0.3%) |
| Gender minorities | 2 (0.5%) | 1 (0.1%) | 3 (0.3%) |
| Autopsies | 1 (0.3%) | 1 (0.1%) | 2 (0.2%) |

*Abbreviations:* IQR, interquartile range; SR, systematic review

* We grouped this variable according to the populations defined by the study authors.

† n (%)

‡ Authors only reported included studies in adults without any specifications.

**Table S5.**Epidemiological study designs included in systematic reviews of prevalence in adult populations published 2010-2020, by total frequency

| Study designs included | SR without meta-analysis<br>n=387* | SR with meta-analysis<br>n=785* | Total<br>n=117* |
|---|---|---|---|
| Studies estimating the target prevalence for the review | 250 (65%) | 373 (48%) | 623 (53%) |
| Observational | 79 (20%) | 261 (33%) | 340 (29%) |
| Cross-sectional | 23 (5.9%) | 60 (7.6%) | 83 (7.1%) |
| Population based studies | 21 (5.4%) | 48 (6.1%) | 69 (5.9%) |
| Cohort Studies | 2 (0.5%) | 12 (1.5%) | 14 (1.2%) |
| Surveys | 5 (1.3%) | 6 (0.8%) | 11 (0.9%) |
| Prospective/ retrospective | 3 (0.8%) | 7 (0.9%) | 10 (0.9%) |
| Community-based studies | 3 (0.8%) | 3 (0.4%) | 6 (0.5%) |
| Case-control studies | 0 (0%) | 5 (0.6%) | 5 (0.4%) |
| Controlled studies | 0 (0%) | 5 (0.6%) | 5 (0.4%) |
| Clinical trials | 1 (0.3%) | 2 (0.3%) | 3 (0.3%) |
| Quantitative studies | 0 (0%) | 3 (0.4%) | 3 (0.3%) |

*Abbreviations:* IQR, interquartile range; SR, systematic review

[a] n (%)

**Table S6.** Summary measures and methods to conduct meta-analysis of prevalence systematic reviews of prevalence in adult populations published 2010-2020

| **Characteristic** | | **n (%)** |
|---|---|---|
| **Systematic reviews without meta-analysis (n=387)** | | |
| Summary measures reported | Prevalence | 380 (98.2) |
| | Mean prevalence | 4 (1.1) |
| | Median prevalence | 2 (0.7) |
| | Prevalence Odds Ratio | 1 (0.3) |
| **Systematic reviews with meta-analysis (n=785)** | | |
| Summary measures reported | Pooled prevalence | 713 (90.8) |
| | Odds ratio | 51 (6.5) |
| | Risk ratio | 12 (1.5) |
| | Mean prevalence | 6 (0.8) |
| | Median prevalence | 2 (0.3) |
| Model | Random-effects model | 703 (89.6) |
| | Unclear or not reported | 47 (6.6) |
| | Both fixed and random effects | 19 (2.4) |
| | Fixed-effects model | 10 (1.2) |
| | Mixed effects model | 1 (0.12) |
| Transformation functions | Freeman-Tukey double arcsine | 131 (17) |
| | Logit | 26 (3.3) |
| | Log | 6 (0.8) |
| Methods used to assess heterogeneity | $I^2$ | 720 (91.7) |
| | $I^2$ and $\tau^2$ | 16 (2) |
| | $\tau^2$ | 5 (0.6) |
| | Meta-regression | 3 (0.4) |
| | Not reported | 41 (5.2) |

**Table S7.** Median PRISMA score in systematic reviews of prevalence in adult populations published 2010-2020, by year of publication

| Year | SR without meta-analysis n=387 | | SR with meta-analysis n=785 | |
|---|---|---|---|---|
| | **Median (IQR)** | **Min, max** | **Median (IQR)** | **Min, max** |
| 2010 | 16.0 (14.2, 18.2) | 12.5, 20.0 | 20.5 (19.6, 22.0) | 13.5, 23.5 |
| 2011 | 15.5 (13.5, 17.6) | 11.0, 22.0 | 21.5 (20.0, 22.1) | 14.0, 25.0 |
| 2012 | 17.2 (16.3, 19.1) | 12.5, 22.5 | 20.5 (19.5, 22.0) | 14.5, 24.5 |
| 2013 | 16.5(15.0, 18.0) | 11.5, 20.0 | 21.5 (19.5, 22.2) | 15.5, 24.0 |
| 2014 | 17.5 (14.0, 19.0) | 10.5, 21.0 | 21.5 (19.5, 23.0) | 10.0, 25.0 |
| 2015 | 17.2 (14.5, 19.0) | 10.5, 22.0 | 21.5 (19., 22.5) | 17.0, 25.0 |
| 2016 | 18.5 (14.7, 19.5) | 9.5, 21.0 | 22.0 (20.1, 23.5) | 17.5, 25.0 |
| 2017 | 17.7 (15.7, 20.1) | 10.0, 21.5 | 22.0 (21.0, 23.5) | 14.0, 25.0 |
| 2018 | 18.5 (15.5, 19.5) | 10.5, 23.0 | 22.5 (21.0, 23.5) | 14.0, 25.0 |
| 2019 | 17.5 (16.5, 19.5) | 11.5, 23.0 | 22.5 (20.5, 23.5) | 14.0, 25.0 |
| 2020 | 17.5 (14.5, 18.2) | 8.0, 21.0 | 23.0 (21.0, 24.0) | 15.5, 25.0 |
| Overall | 17.5 IQR (15.0,19.0) | 8.0, 23.0 | 22.0 IQR (20.5,23.5) | 10.0, 25.0 |

*Abbreviations:* IQR, interquartile range; SR, systematic review

**Table S8.** Adequate reporting of PRISMA 2009 items, by year, systematic reviews of prevalence in adult populations published 2010-2020

| Year | 2010 | | 2011 | | 2012 | | 2013 | | 2014 | | 2015 | | 2016 | | 2017 | | 2018 | | 2019 | | 2020 | |
|---|---|---|---|---|---|---|---|---|---|---|---|---|---|---|---|---|---|---|---|---|---|---|
| **PRISMA 2009 item, number and section** | **SR, no MA** n=15[a] | **SR, with MA** n=10[a] | **SR, no MA** n=16[a] | **SR, with MA** n=16[a] | **SR, no MA** n=24[a] | **SR, with MA** n=27[a] | **SR, no MA** n=35[a] | **SR, with MA** n=31[a] | **SR, no MA** n=39[a] | **SR, with MA** n=25[a] | **SR, no MA** n=46[a] | **SR, with MA** n=53[a] | **SR, no MA** n=43[a] | **SR, with MA** n=70[a] | **SR, no MA** n=40[a] | **SR, with MA** n=73[a] | **SR, no MA** n=37[a] | **SR, with MA** n=100[a] | **SR, no MA** n=45[a] | **SR, with MA** n=154[a] | **SR, no MA** n=47[a] | **SR, with MA** n=226[a] |
| **1. Title** | 14 (93) | 10 (100) | 15 (94) | 16 (100) | 22 (92) | 27 (100) | 34 (97) | 28 (90) | 37 (95) | 25 (100) | 43 (93) | 52 (98) | 39 (91) | 68 (97) | 39 (97) | 72 (99) | 35 (95) | 99 (99) | 42 (93) | 153 (99) | 43 (91) | 225(100) |
| **2. Structured summary** | 14 (93) | 10 (100) | 16(100) | 16 (100) | 23 (96) | 27 (100) | 35(100) | 30 (97) | 37 (95) | 24 (96) | 45 (98) | 53 (100) | 42 (98) | 69 (99) | 40(100) | 71 (97) | 36 (97) | 99 (99) | 43 (96) | 153 (99) | 47 (100) | 224 (99) |
| **Introduction** | | | | | | | | | | | | | | | | | | | | | | |
| **3. Rationale** | 13 (87) | 10 (100) | 14 (88) | 16 (100) | 24(100) | 26 (96) | 35(100) | 29 (94) | 34(87) | 22 (88) | 44 (96) | 53 (100) | 41 (95) | 67 (96) | 39 (98) | 72 (99) | 37 (100) | 98 (98) | 45(100) | 151 (98) | 46 (98) | 225(100) |
| **4. Objectives** | 15(100) | 10 (100) | 15 (94) | 15 (94) | 23 (96) | 27 (100) | 34 (97) | 30 (97) | 35(90) | 25 (100) | 44 (96) | 52 (98) | 42 (98) | 70 (100) | 38 (95) | 71 (97) | 37 (100) | 96 (96) | 45(100) | 153 (99) | 44 (94) | 222 (98) |
| **Methods** | | | | | | | | | | | | | | | | | | | | | | |
| **5. Protocol** | 1 (7) | 0 (0) | 0 (0) | 1 (6) | 3 (13) | 0 (0) | 6 (17) | 1 (3) | 3 (8) | 3 (12) | 6 (13) | 8 (15) | 10 (23) | 18 (26) | 7 (18) | 24 (33) | 8 (22) | 38 (38) | 11 (24) | 48 (31) | 9 (19) | 91 (40) |
| **6. Eligibility criteria** | 15(100) | 9 (90) | 16(100) | 16 (100) | 24(100) | 26 (96) | 32 (91) | 31 (100) | 39(100) | 25 (100) | 44 (96) | 51 (96) | 40 (93) | 70 (100) | 39 (98) | 73 (100) | 36 (97) | 99 (99) | 41 (91) | 152 (99) | 45 (96) | 223 (99) |
| **7. Information sources** | 14 (93) | 10 (100) | 15 (94) | 16 (100) | 20 (83) | 27 (100) | 34 (97) | 30 (97) | 38 (97) | 25 (100) | 43 (93) | 52 (98) | 38 (88) | 66 (94) | 39 (98) | 71 (97) | 37(100) | 94 (94) | 39 (87) | 147 (95) | 45 (96) | 223 (99) |
| **8. Search** | 1 (7) | 2 (20) | 6 (38) | 7 (44) | 10 (42) | 9 (33) | 15 (43) | 15 (48) | 15 (38) | 15 (60) | 20 (43) | 30 (57) | 23 (53) | 36 (51) | 19 (48) | 42 (58) | 12 (32) | 55 (55) | 15 (33) | 90 (58) | 22 (47) | 148 (65) |
| **9. Study selection** | 9 (60) | 9 (90) | 12 (75) | 14 (88) | 22 (92) | 22 (81) | 30 (86) | 29 (94) | 28 (72) | 23 (92) | 30 (65) | 47 (89) | 36 (84) | 65 (93) | 33 (83) | 69 (95) | 27 (73) | 92 (92) | 36 (80) | 145 (94) | 37 (79) | 214 (95) |
| **10. Data collection process** | 10 (67) | 9 (90) | 12 (75) | 13 (81) | 22 (92) | 26 (96) | 24 (69) | 30 (97) | 31 (79) | 24 (96) | 35 (76) | 45 (85) | 35 (81) | 65 (93) | 30 (75) | 66 (90) | 31 (84) | 96 (96) | 37 (82) | 141 (92) | 29 (62) | 195 (86) |
| **11. Data items** | 12 (80) | 10 (100) | 13 (81) | 15 (94) | 24(100) | 27 (100) | 32 (91) | 30 (97) | 33 (85) | 22 (88) | 38 (83) | 51 (96) | 42 (98) | 68 (97) | 36 (90) | 71 (97) | 33 (89) | 94 (94) | 36 (80) | 147 (95) | 38 (81) | 209 (92) |
| **12. Risk of bias in individual studies** | 7 (47) | 3 (30) | 4 (25) | 10 (63) | 9 (38) | 16 (59) | 10 (29) | 19 (61) | 19 (49) | 15 (60) | 23 (50) | 32 (60) | 23 (53) | 53 (76) | 24 (60) | 55 (75) | 27 (73) | 84 (84) | 30 (67) | 124 (81) | 25 (53) | 186 (82) |
| **13. Summary measures** | 11 (73) | 9 (90) | 10 (63) | 15 (94) | 21 (88) | 27 (100) | 22 (63) | 29 (94) | 28 (72) | 23 (92) | 32 (70) | 49 (92) | 28 (65) | 67 (96) | 29 (73) | 69 (95) | 31 (84) | 98 (98) | 32 (71) | 144 (94) | 26 (55) | 191 (85) |
| **14. Synthesis of results**[b] | NA | 9 (90) | NA | 12 (75) | NA | 18 (67) | NA | 19 (61) | NA | 16 (64) | NA | 40 (75) | NA | 44 (63) | NA | 52 (71) | NA | 74 (74) | NA | 128 (83) | NA | 184 (81) |
| **15. Risk of bias across studies** | Not assessed in any systematic reviews of prevalence | | | | | | | | | | | | | | | | | | | | | |
| **16. Additional analyses** | 0 (0) | 8 (80) | 1 (6) | 11 (69) | 4 (17) | 12 (44) | 2 (6) | 19 (61) | 3 (8) | 17 (68) | 2 (4) | 31 (58) | 1 (2) | 46 (66) | 5 (13) | 53 (73) | 5 (14) | 71 (71) | 6 (13) | 97 (63) | 2 (4) | 161 (71) |
| **Results** | | | | | | | | | | | | | | | | | | | | | | |
| **17. Study selection** | 13 (87) | 10 (100) | 13 (81) | 15 (94) | 23 (96) | 27 (100) | 32 (91) | 30 (97) | 35 (90) | 24 (96) | 44 (96) | 53 (100) | 39 (91) | 70 (100) | 34 (85) | 72 (99) | 37 (100) | 98 (98) | 44 (98) | 154 (100) | 43 (91) | 223 (99) |
| **18. Study characteristics** | 14 (93) | 10 (100) | 16(100) | 15 (94) | 22 (92) | 24 (89) | 32 (91) | 28 (90) | 33 (85) | 22 (88) | 41 (89) | 51 (96) | 43(100) | 67 (96) | 38 (95) | 69 (95) | 33 (89) | 93 (93) | 42 (93) | 146 (95) | 41 (87) | 222 (98) |

| Year | 2010 | | 2011 | | 2012 | | 2013 | | 2014 | | 2015 | | 2016 | | 2017 | | 2018 | | 2019 | | 2020 | |
|---|---|---|---|---|---|---|---|---|---|---|---|---|---|---|---|---|---|---|---|---|---|---|
| **19. Risk of bias within studies** | 7 (47) | 2 (20) | 4 (25) | 6 (38) | 7 (29) | 14 (52) | 10 (29) | 16 (52) | 13 (33) | 11 (44) | 21 (46) | 29 (55) | 21 (49) | 46 (66) | 19 (48) | 45 (62) | 20 (54) | 69 (69) | 27 (60) | 179 (71) | 18 (38) | 161 (71) |
| **20. Results of individual studies** | 15(100) | 10 (100) | 15 (94) | 15 (94) | 21 (88) | 25 (93) | 34 (97) | 29 (94) | 37 (95) | 21 (84) | 41(89) | 51 (96) | 43 (100) | 67 (96) | 37(93) | 71 (97) | 36 (97) | 89 (89) | 41(91) | 146 (95) | 41 (87 ) | 222 (98) |
| **21. Synthesis of results[b]** | NA | 9 (90) | NA | 16 (100) | NA | 26 (96) | NA | 29 (94) | NA | 21 (84) | NA | 50 (94) | NA | 66 (94) | NA | 67 (92) | NA | 93 (93) | NA | 151 (98) | NA | 208 (92) |
| **22. Risk of bias across studies** | Not assessed in any systematic reviews of prevalence | | | | | | | | | | | | | | | | | | | | | |
| **23. Additional analysis** | 0 (0) | 8 (80) | 2 (13) | 11 (69) | 6 (25) | 14 (52) | 2 (6) | 20 (65) | 4 (10) | 17 (68) | 2 (4) | 29 (55) | 2 (5) | 45 (64) | 5 (13) | 50 (68) | 3 (8) | 71 (71) | 8 (18) | 102 (66) | 2 (4) | 173 (77) |
| **Discussion** | | | | | | | | | | | | | | | | | | | | | | |
| **24. Summary of evidence** | 15(100) | 10 (100) | 15 (94) | 16 (100) | 24(100) | 27 (100) | 35(100) | 31 (100) | 39(100) | 25 (100) | 44 (96) | 52 (98) | 41 (95) | 70 (100) | 39 (98) | 73 (100) | 37(100) | 99 (99) | 44 (98) | 154 (100) | 47 (100) | 226(100) |
| **25. Limitations** | 12 (80) | 7 (70) | 10 (63) | 14 (88) | 20 (83) | 22 (81) | 24 (69) | 24 (77) | 26 (67) | 22 (88) | 35 (76) | 44 (83) | 32 (74) | 62 (89) | 33 (83) | 67 (92) | 29 (78) | 81 (81) | 34 (76) | 140 (91) | 38 (81) | 203 (90) |
| **26. Conclusions** | 15(100) | 10 (100) | 15 (94) | 16 (100) | 22 (92) | 27 (100) | 35(100) | 31 (100) | 35 (90) | 25 (100) | 43 (93) | 52 (98) | 42 (98) | 68 (97) | 39 (98) | 73 (100) | 36 (97) | 100 (100) | 44 (98) | 151 (98) | 45 (96) | 225(100) |
| **27. Funding** | 8 (53) | 4 (40) | 8 (50) | 12 (75) | 16 (67) | 16 (59) | 13 (37) | 17 (55) | 26 (67) | 17 (68) | 28 (61) | 36 (68) | 25 (58) | 47 (67) | 28 (70) | 49 (67) | 17 (46) | 74 (74) | 28 (62) | 93 (60) | 27 (57) | 175 (77) |

*Abbreviations:* IQR, interquartile range; NA, not applicable; SR, systematic review

[a] n (%)

[b] Items 14 and 21 were assessed for quantitative syntheses of results. These items are not applicable to systematic reviews without meta-analysis and are highlighted in grey.

**Table S9.** Reporting of risk of bias assessment and reported tools used, systematic reviews of prevalence in adult populations published 2010-2020

| Tools used to conduct risk of bias assessment | SR without meta-analysis N=387* | SR with meta-analysis N=785* | Total N=1172* |
|---|---|---|---|
| The Newcastle-Ottawa Scale for assessing the quality of non-randomised studies [1] | 30 (7.8%) | 123 (15.7%) | 153 (13.1%) |
| Authors' adaptation of an existing tool | 29 (7.5%) | 68 (8.7%) | 97 (8.3%) |
| JBI tool for prevalence studies [2]† | 13 (3.4%) | 65 (8.3%) | 78 (6.7%) |
| Strengthening the Reporting of Observational studies in Epidemiology (STROBE) [3] | 16 (4.1%) | 62 (7.9%) | 78 (6.7%) |
| Tool designed by authors | 25 (6.5%) | 51 (6.5%) | 76 (6.5%) |
| Hoy et al. [4] † | 11 (2.8%) | 49 (6.2%) | 60 (5.1%) |
| Loney et al [5] † | 10 (2.6%) | 22 (2.8%) | 32 (2.7%) |
| Agency for Healthcare Research & Quality (AHRQ) [6] | 4 (1.0%) | 17 (2.2%) | 21 (1.8%) |
| Munn et al. [7]† | 4 (1.0%) | 14 (1.8%) | 18 (1.5%) |
| Boyle et al [8] † | 6 (1.6%) | 8 (1.0%) | 14 (1.2%) |
| National Heart and Lung Institute [9] | 6 (1.6%) | 9 (1.1%) | 15 (1.3%) |
| Critical appraisal tool to assess the quality of cross-sectional studies [10] | 1 (0.3%) | 13 (1.7%) | 14 (1.2%) |
| Cochrane Collaboration risk of bias tool [11] | 3 (0.8%) | 9 (1.1%) | 12 (1.0%) |
| Critical Appraisal Skills Program (CASP). Tool for cohort studies or case-control studies [12] | 4 (1.0%) | 7 (0.9%) | 11 (0.9%) |
| Leboeuf-Yde et al. [13]† | 3 (0.8%) | 2 (0.3%) | 5 (0.4%) |
| Methodological Evaluation of Observational Research [14]† | 1 (0.3%) | 4 (0.5%) | 5 (0.4%) |
| JBI tools for cross-sectional or cohort studies [15] | 2 (0.5%) | 3 (0.4%) | 6 (0.5%) |
| Luppa et al. [16] | 2 (0.5%) | 2 (0.3%) | 4 (0.3%) |
| Methodological index for non-randomised studies (MINORS) [17] | 0 (0%) | 4 (0.5%) | 4 (0.3%) |
| Reporting Guidelines for Meta-analyses of Observational Studies (MOOSE) [18] | 1 (0.3%) | 3 (0.4%) | 4 (0.3%) |
| QUADAS-2 [19] | 0 (0%) | 4 (0.5%) | 4 (0.3%) |
| Quality assessment tool for systematic reviews of observational studies (QATSO) [20] | 2 (0.5%) | 2 (0.3%) | 4 (0.3%) |
| Jadad Scale [21] | 1 (0.3%) | 2 (0.3%) | 3 (0.3%) |
| JBI tool for randomised controlled trials [15] | 0 (0%) | 3 (0.4%) | 2 (0.2%) |
| Grading of Recommendations, Assessment, Development, and Evaluations (GRADE) [22] | 0 (0%) | 2 (0.3%) | 2 (0.2%) |
| Preferred Reporting Items for Systematic Reviews and Meta-Analyses -PRISMA [23] | 0 (0%) | 2 (0.3%) | 2 (0.2%) |

**Table S9.** Reporting of risk of bias assessment and reported tools used, systematic reviews of prevalence in adult populations published 2010-2020

| **Tools used to conduct risk of bias assessment** | **SR without meta-analysis** N=387* | **SR with meta-analysis** N=785* | **Total** N=1172* |
|---|---|---|---|
| ROBINS-I [24] | 1 (0.3%) | 1 (0.1%) | 2 (0.2%) |
| Oxford Levels of Evidence [25] | 0 (0%) | 2 (0.3%) | 2 (0.2%) |
| Giannakopoulos et al. [26]† | 0 (0%) | 1 (0.1%) | 1 (0.1%) |
| Other tools | 30 (7.8%) | 55 (7.0%) | 85 (7.3%) |
| Not described/unclear | 2 (0.5%) | 10 (1.3%) | 12 (1.0%) |
| Did not report any risk of bias assessment | 180 (46.5%) | 166 (21.1%) | 346 (29.5%) |

*Abbreviations:* SR, systematic review

* n (%)

† Tool designed for prevalence studies
